# ISARIC COVID-19 Clinical Data Report: Final report January 2020 – January 2023

**DOI:** 10.1101/2020.07.17.20155218

**Authors:** ISARIC Clinical Characterisation Group, J. Kenneth Baillie, Joaquin Baruch, Abigail Beane, Lucille Blumberg, Fernando Augusto Bozza, Tessa Broadley, Aidan Burrell, Gail Carson, Barbara Wanjiru Citarella, Jake Dunning, Loubna Elotmani, Noelia Garcia Barrio, Jean-Christophe Goffard, Matthew Hall, Madiha Hashmi, Peter Horby, Waasila Jassat, Christiana Kartsonaki, Bharath Kumar Tirupakuzhi Vijayaraghavan, Pavan Kumar Vecham, Cedric Laouenan, Samantha Lissauer, Ignacio Martin-Loeches, France Mentre, Ben Morton, Daniel Munblit, Nikita A. Nekliudov, Alistair Nichol, David S.Y. Ong, Prasan Kumar Panda, Miguel Pedrera Jimenez, Michelle Petrovic, Nagarajan Ramakrishnan, Grazielle Viana Ramos, Claire Roger, Amanda Rojek, Oana Sandulescu, Malcolm G. Semple, Pratima Sharma, Sally Shrapnel, Louise Sigfrid, Benedict Sim Lim Heng, Budha Charan Singh, Emily Somers, Anca Streinu-Cercel, Fabio S. Taccone, Jia Wei, Evert-Jan Wils, Xin Ci Wong, Kyle Young, Piero L. Olliaro, Laura Merson

## Abstract

ISARIC (International Severe Acute Respiratory and emerging Infections Consortium) partnerships and outbreak preparedness initiatives enabled the rapid launch of standardised clinical data collection on COVID-19 in Jan 2020. Extensive global participation has resulted in a large, standardised collection of comprehensive clinical data from hundreds of sites across dozens of countries. Data are analysed regularly and reported publicly to inform patient care and public health response. This report, our 18th and final report, is a part of a series published over 3 years. Data have been entered for 945,317 individuals from 1807 partner institutions and networks across 76 countries.

The comprehensive analyses detailed in this report includes hospitalised individuals of all ages for whom data collection occurred between 30 January 2020 and up to and including 10 January 2023, AND who have laboratory-confirmed SARS-COV-2 infection or clinically diagnosed COVID-19.

For the 845,291 cases who meet eligibility criteria for this report, selected findings include:

- Median age of 57 years, with an approximately equal (50/50) male:female sex distribution
- 29% of the cohort are at least 70 years of age, whereas 6% are 0-19 years of age
- The most common symptom combination in this hospitalised cohort is shortness of breath, cough, and history of fever, which has remained constant over time
- The five most common symptoms at admission were shortness of breath, cough, history of fever, fatigue/malaise, and altered consciousness/confusion, which is unchanged from the previous reports
- Age-associated differences in symptoms are evident, including the frequency of altered consciousness increasing with age, and fever, respiratory and constitutional symptoms being present mostly in those 40 years and above
- 15% of patients with relevant data available (845,291) were admitted at some point during their illness into an intensive care unit (ICU), which has decreased from 19% during the 3 years of ISARIC reporting
- Antibiotic agents were used in 37% of patients for whom relevant data are available (802,241), a significant reduction from our previous reports (80%) which reflects a shifting proportion of data contributed by different institutions; in ICU/HDU admitted patients with data available (64,669), 90% received antibiotics
- Use of corticosteroids was reported in 25% of all patients for whom data were available (809,043); in ICU/HDU admitted patients with data available (64,713), 71% received corticosteroids
- Outcomes are known for 762,728 patients and the overall estimated case fatality ratio (CFR) is 22% (95%CI 21.9-22), rising to 36% (95%CI 35.6-36.1) for patients who were admitted to ICU/HDU, demonstrating worse outcomes in those with the most severe disease

To access previous versions of **ISARIC COVID-19 Clinical Data Report** please use the link below: https://isaric.org/research/covid-19-clinical-research-resources/evidence-reports/

## Background

This report describes the clinical presentation, treatment and outcome of a cohort of over 945,000 patients with COVID-19 hospitalised between January 2020 and January 2023 in 76 countries. This has been possible thanks to 1807 institutions and their clinical teams contributing to ISARIC (International Severe Acute Respiratory and Emerging Infection Consortium), a grassroots collaboration https://isaric.org/.

After over 750 million cases and over 6 million deaths, COVID-19 has shown us how pandemics disrupt populations by stretching healthcare systems, and shaking economies. Although clinicians and well-equipped surveillance systems can adapt to health emergencies by implementing new treatments and information systems, clinical research lacks this agility.

### The advantages of an international standardised case record form

Novel diseases are, by definition, new; the scientific method is not. By developing consensus on a road map for clinical research methods before a pandemic, we are able to minimise further time defining study types, data collection tools, and potential outputs for evidence summarisation once the pathogen emerges. Using slightly adjusted standardised methods for novel diseases, evidence can be generated quickly.

While small observational studies are easy to conduct, they often record different data or may record variables in different ways. This may not allow data to be collated or findings to be compared across studies, or combined to produce a more powerful and accurate representation. In contrast, large international observational studies with a standardised form allow for international data comparisons, improved analytic control of confounders, and therefore enable the acquisition of high levels of evidence.

### How we got here with ISARIC

In 2013, ISARIC and the World Health Organisation (WHO) implemented the standardised Clinical Characterisation Protocol to evolve clinical data into research evidence during health emergencies.

In January 2020, we launched our case report form when only 846 COVID-19 cases had been reported globally. The first patient was enrolled on 30 January 2020. Today, our partners have collected standardised, in-hospital clinical data from over 945,000 patients, in 1807 sites, across 76 countries. More than half of these patients live in low- and middle-income countries. This partner-fuelled initiative is to our knowledge the largest COVID-19 in-hospital international database in the world.

In this report, we present the current state of our database, which all our collaborators can access by submitting a statistical analysis plan to our clinical team. We encourage our collaborators to test their research hypothesis within the framework of ISARIC’s collaborative Partner Analysis scheme [2]. The majority of this database is additionally available to external researchers via application to our Data Access Committee at https://www.iddo.org/covid19/data-sharing/accessing-data

These analyses are available on the ISARIC COVID-19 Dashboard at https://livedataoxford.shinyapps.io/CovidClinicalDataDashboard/

To contribute, please contact

## Methods

The results in this report were produced using data from the ISARIC COVID-19 database of international, prospective observational data on clinical features of patients admitted to hospital with COVID-19. Data collection was structured on the ISARIC/WHO Clinical Characterisation Protocol for Severe Emerging Infections, a standardized protocol for investigation of severe acute infections by pathogens of public health interest [3] and the ISARIC case report forms [4] designed for standardized data collection from the start of an outbreak and rapid dissemination of clinical information [5–9]. Data were collected on Research electronic Data Capture (REDCap, version 8.11.11, Vanderbilt University, Nashville, Tenn.), hosted by the University of Oxford. Additional data, collected using a wide variety of data systems, were submitted by international investigators who were not using the University of Oxford REDCap instance. Data were curated by the Infectious Diseases Data Observatory [10] to the CDISC Study Data Tabulation Model (SDTM) for harmonised analysis [11].

SDTM formatted data were processed in R 4.1.0 [12]. Initial data cleaning included custom scripts designed to identify results of laboratory SARS-CoV-2 testing, and to standardise nomenclature for comorbidities, symptoms at admission, treatments, and outcomes.

Patients were excluded if they did not have laboratory or clinically confirmed SARS-CoV-2 infection. Patients were considered to be lost to follow up if either a) they were transferred to another facility, or b) they had an unknown outcome and the last date upon which any data was recorded for them was 45 days or before the date of data extraction. Patients with unknown outcome where the last recorded data was less than 45 days old were instead categorised as receiving ongoing care.

Comorbidities, symptoms at admission, and treatments (in both the full population and the intensive care unit (ICU)/high dependency unit (HDU) population) were included in the report only if at least 10% of patients had their presence or absence recorded. Laboratory and vital sign measurements that fell within the top 2.5% and bottom 2.5% of values, within both the <10 year age group and the ≥ 10 year age group, were taken to be likely mis-entered and changed to missing for this report. Similarly, variables for time durations were converted to missing if their values were either negative or in the top 2.5% of values. Dates were converted to missing where they were before the 1st January 2020 (except for dates of birth and hospital admission) or after the date of data extraction.

The following COVID-19 symptom definitions were used:

- WHO:
- A combination of acute fever and cough, Or
- A combination of three or more of: fever, cough, general weakness and fatigue, headache, myalgia (we used muscle aches, joint pain), sore throat, coryza (we used runny nose), dyspnoea (we used shortness of breath), anorexia, nausea and vomiting, diarrhoea, altered mental status (we used altered consciousness or confusion)
- Centers for Disease Control (CDC), United States:

1. At least two of: fever, chills (not available), rigors (not available), myalgia, headache, sore throat, new olfactory and taste disorder, Or
2. At least one of: cough, shortness of breath, difficulty breathing (not available)

- UK Health Security Agency:

New cough (we used cough), or temperature ≥ 37.8°C (we used history of fever), or a loss or change in sense of smell or taste

- European Centre for Disease Prevention and Control:

At least one of: cough, fever, shortness of breath, sudden onset anosmia, ageusia or dysgeusia Data analysis was performed using the tidyverse collection of packages [13], as well as ff [14] and dtplyr [15]. Data visualisation used ggplot [16] and its extension ggupset [17]. The report was generated using knitr [18]. This report included individuals for whom data collection commenced on or before 10 January 2023.

## Results

The flowchart (Figure 1) gives an overview of the cohort and outcomes as of 10 January 2023.

**Figure 1:**
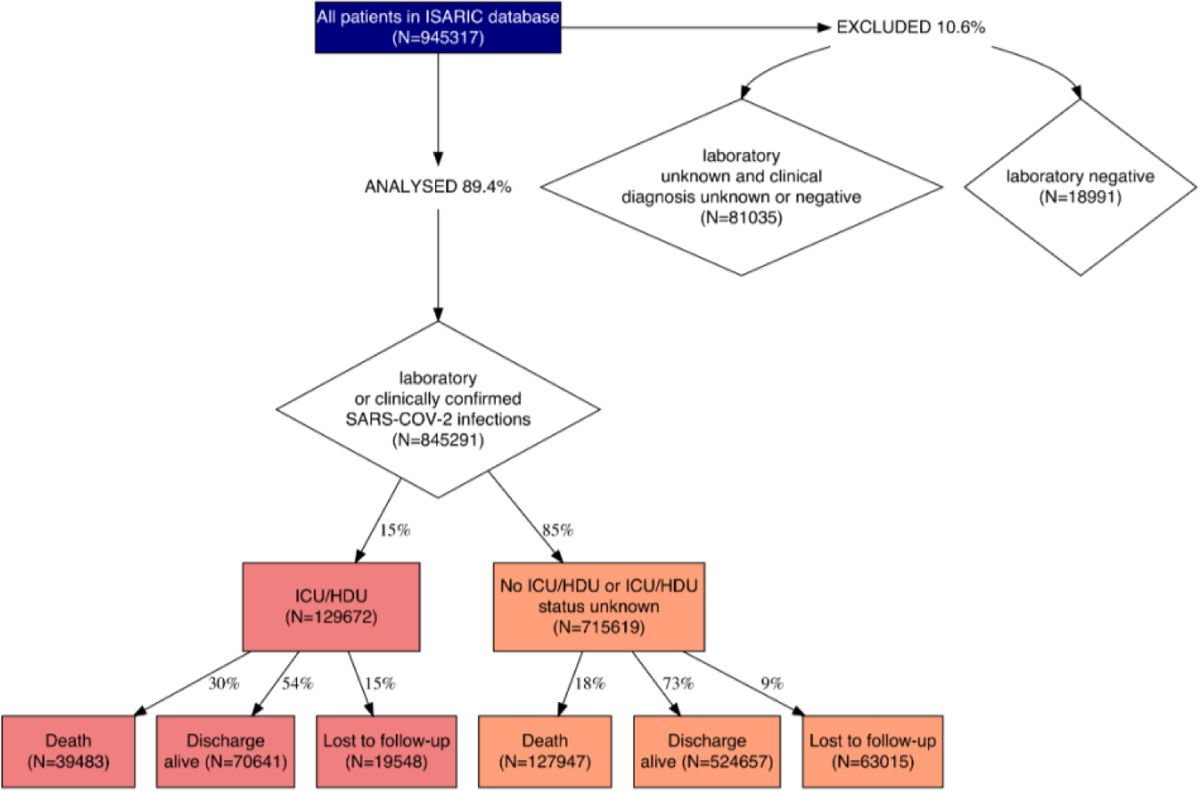
Overview of cohort and outcomes as of 10 January 2023.

Data were contributed by 1807 sites in 76 countries (see Figure 2). 555635 (58.8%) of the patients were from low- and middle-income and 389603 (41.2%) from high-income countries. The two largest contributors to the database were South Africa (56.5%) and the UK (31.5%). Plots grouped by regions and calendar time are shown in Figure 3 and 4. The cohort satisfying the inclusion criteria had **845291** cases.

**Figure 2:**
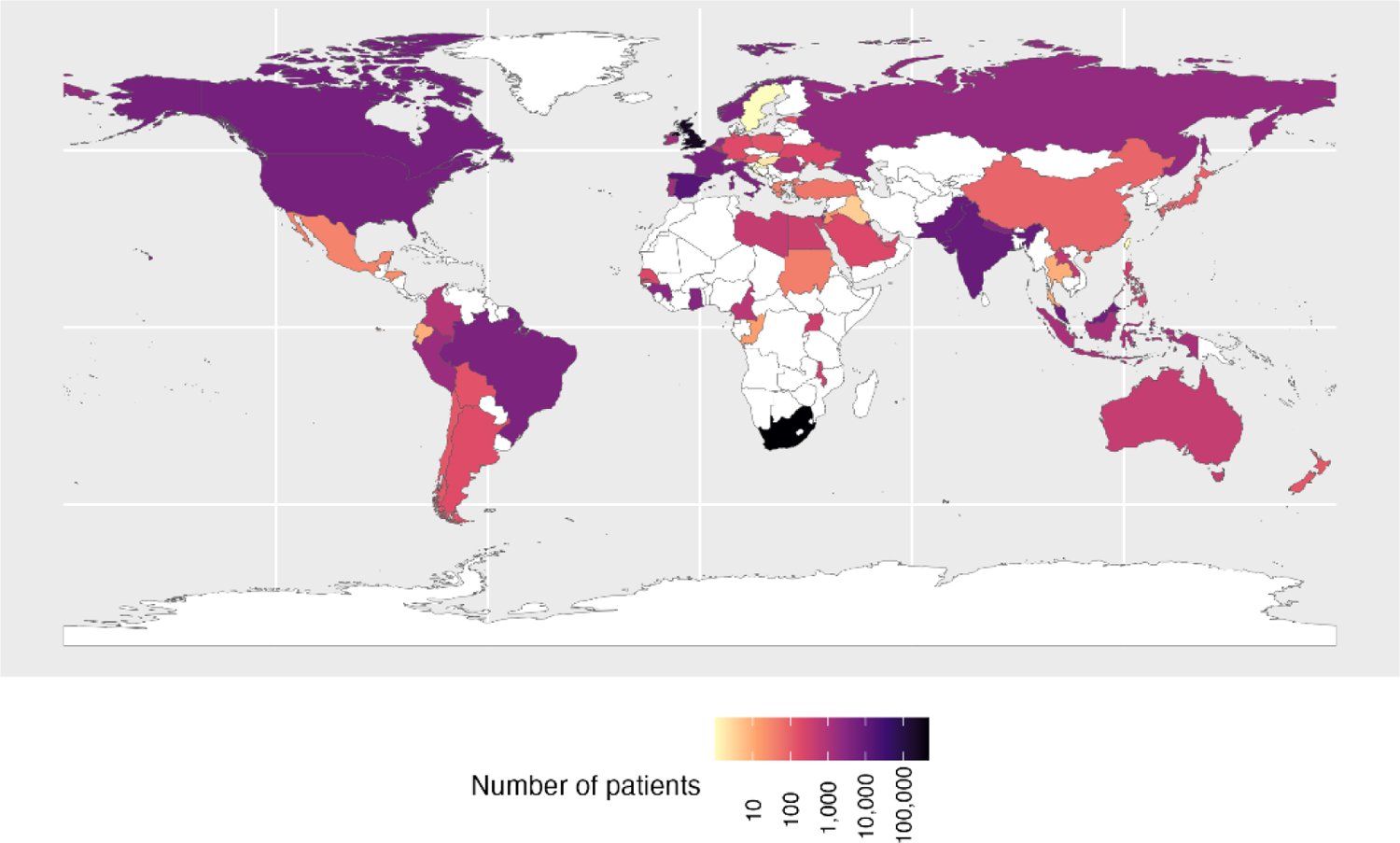
Distribution of patients by country

**Figure 3:**
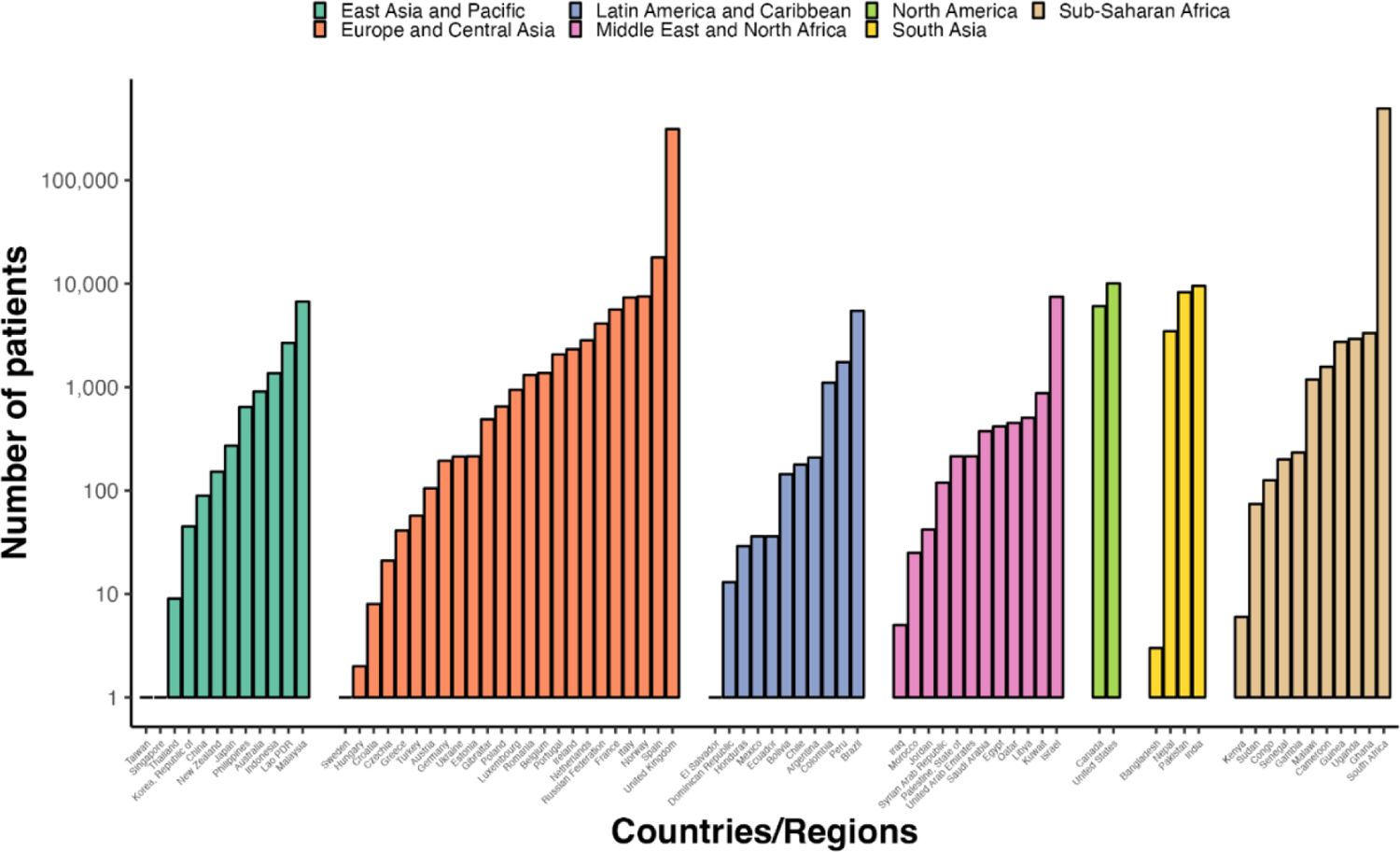
Distribution of patients by world bank region and country

**Figure 4:**
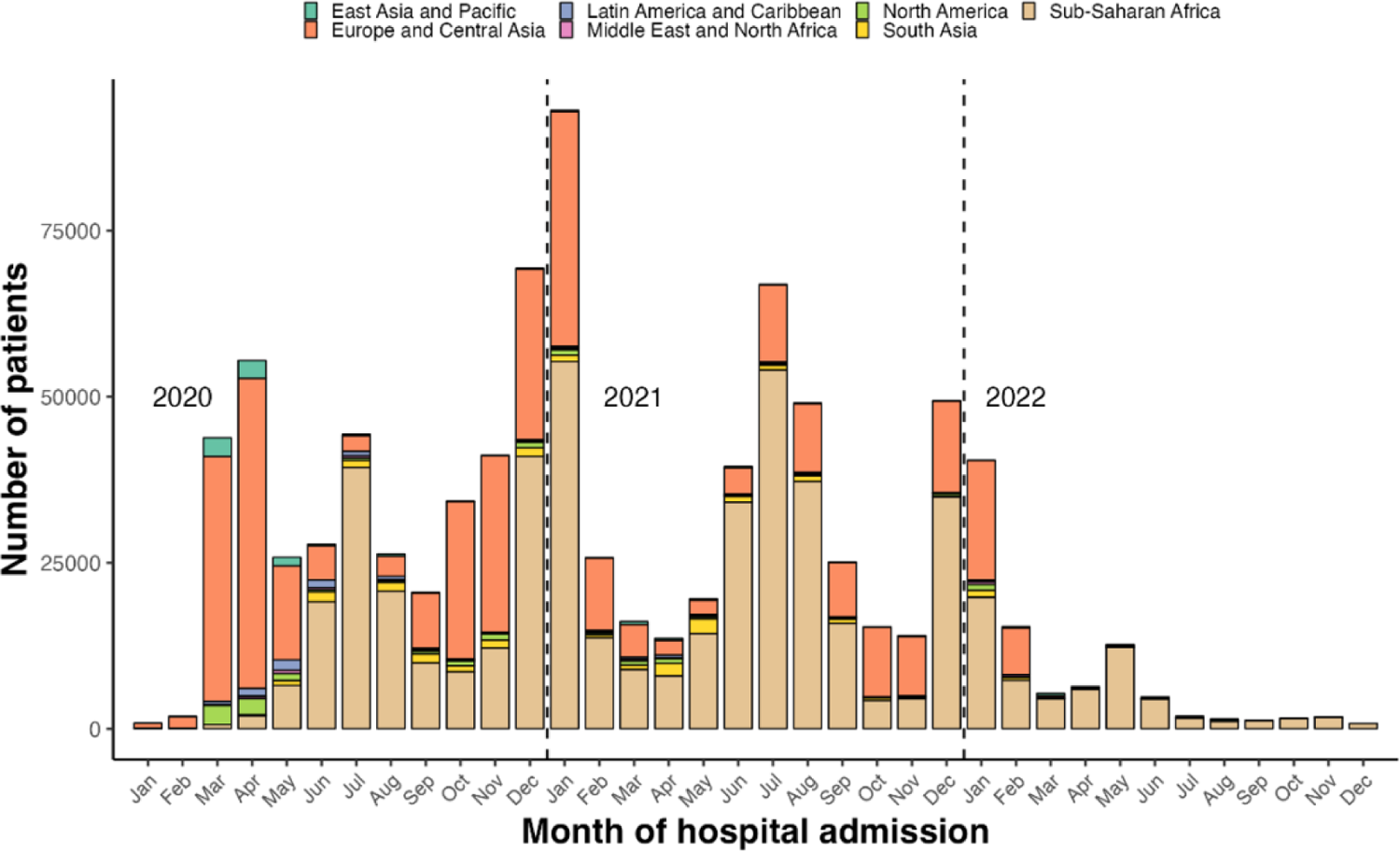
Distribution of patients by world bank region and time

### Demographics

Of these 845291 cases, 416555 (49.3%) were males and 427476 (50.6%) were females. Sex was unreported for 1260 cases. The median age was 57 years, ranging from 0 to 120 years. One-third of the patients were 70 years old and above. Demographics, including age, sex, and outcome, are presented in Table 1 and Figure 5.

**Figure 5:**
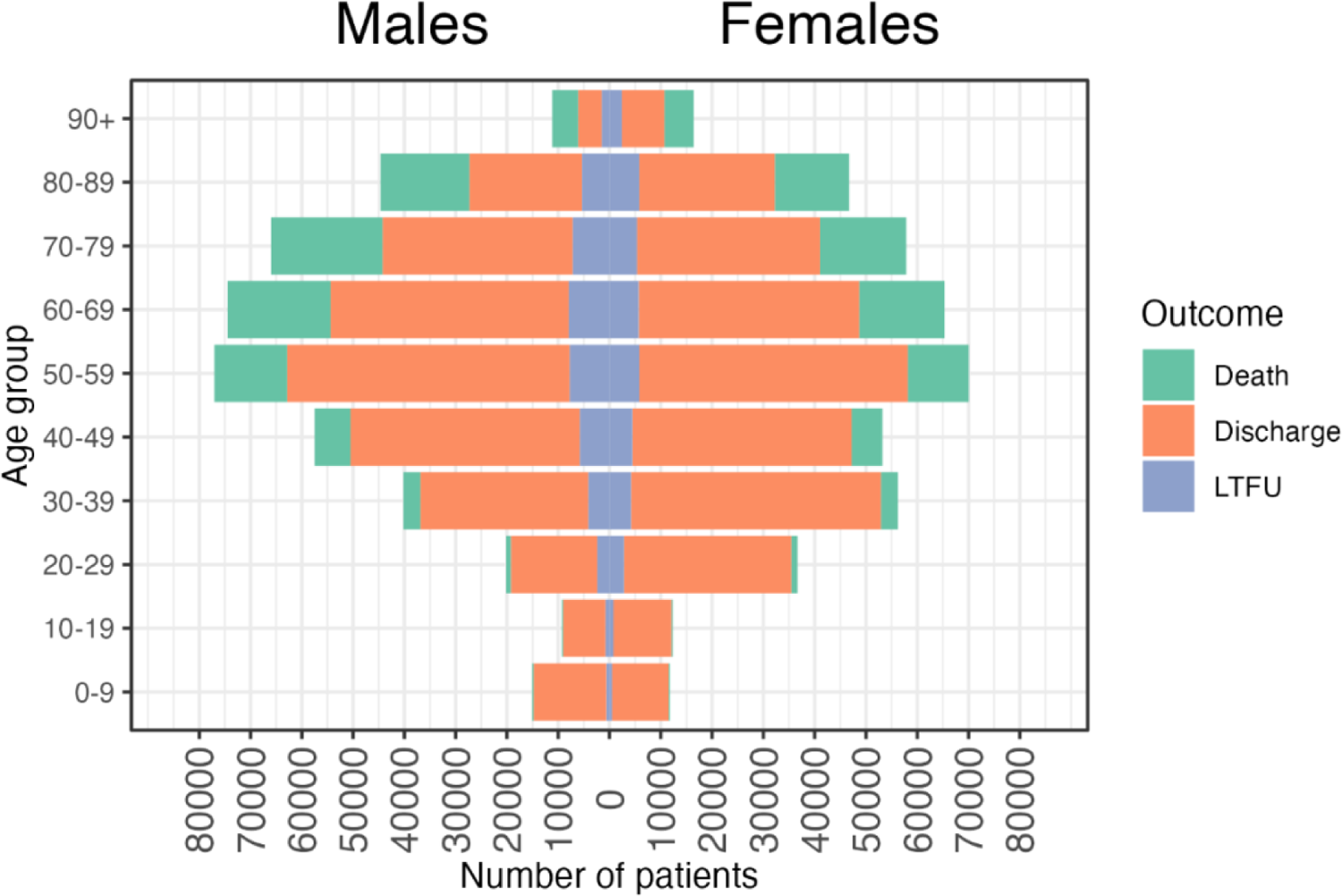
Age and sex distribution of patients. Bar fills are outcome (death/discharge/lost to follow-up (LTFU)) at the time of report.

**Table 1:** Patient Characteristics. Proportions are presented in parentheses, and have been rounded to two decimal places.

| Variable | Count (proportion) |
| --- | --- |
| Size of cohort | 845291 |
| By sex |  |
| Female | 427476 (0.51) |
| Male | 416555 (0.49) |
| Unknown | 1260 (0) |
| By outcome status |  |
| Death | 167430 (0.20) |
| Discharge | 595298 (0.70) |
| LTFU | 82563 (0.10) |
| By age group |  |
| 0-9 | 26898 (0.03) |
| 10-19 | 21659 (0.03) |
| 20-29 | 56907 (0.07) |
| 30-39 | 96540 (0.11) |
| 40-49 | 110828 (0.13) |
| 50-59 | 147213 (0.17) |
| 60-69 | 139938 (0.17) |
| 70+ | 243126 (0.29) |
| Unknown | 2182 (0) |

**Table 2:** Outcome by age and sex. Proportions are calculated using the row total as the denominator. Case fatality ratio: patients who died/patients not lost to follow up.

| Variable | Death | Discharge | LTFU | Case fatality ratio |
| --- | --- | --- | --- | --- |
| Age |  |  |  |  |
| 0-9 | 564 (0.02) | 25242 (0.94) | 1092 (0.04) | 0.02 |
| 10-19 | 529 (0.02) | 19580 (0.9) | 1550 (0.07) | 0.03 |
| 20-29 | 2235 (0.04) | 49480 (0.87) | 5192 (0.09) | 0.04 |
| 30-39 | 6659 (0.07) | 81562 (0.84) | 8319 (0.09) | 0.08 |
| 40-49 | 13013 (0.12) | 87501 (0.79) | 10314 (0.09) | 0.13 |
| 50-59 | 26145 (0.18) | 107529 (0.73) | 13539 (0.09) | 0.2 |
| 60-69 | 36780 (0.26) | 89487 (0.64) | 13671 (0.1) | 0.29 |
| 70+ | 81377 (0.33) | 134098 (0.55) | 27651 (0.11) | 0.38 |
| Sex |  |  |  |  |
| Female | 76755 (0.18) | 312498 (0.73) | 38223 (0.09) | 0.2 |
| Male | 90497 (0.22) | 282193 (0.68) | 43865 (0.11) | 0.24 |

### Presenting symptoms

The observed mean number of days from (first) symptom onset to hospital admission was 5.4, with a standard deviation (SD) of 4.8 days and a median of 5 days.

A detailed analysis of symptoms from 60109 hospitalized symptomatic patients between 30 January and 3 August 2020 has been reported in a previous publication [19]. The prevalence of symptoms on admission to the hospital was summarised in Figure 6a. (Note that South Africa did not collect symptoms data. The overall frequency of missing data outside South Africa is 11%). The five most common symptoms at admission were shortness of breath, cough, history of fever, fatigue/malaise, and altered consciousness/confusion, accounting for 59%, 57.3%, 55.2%, 37.7%, 17.5% of patients with at least one symptom recorded.

**Figure 6:**
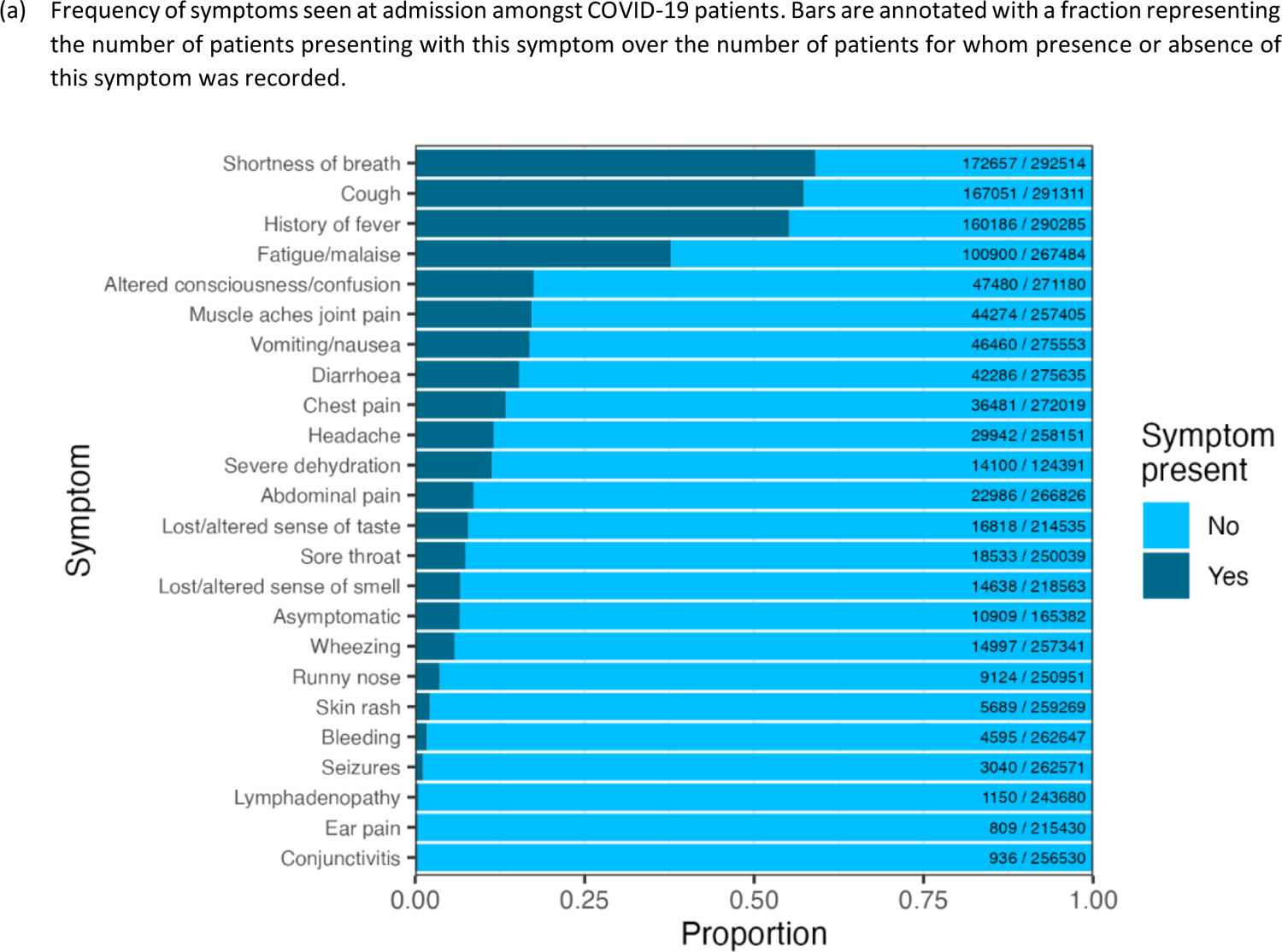
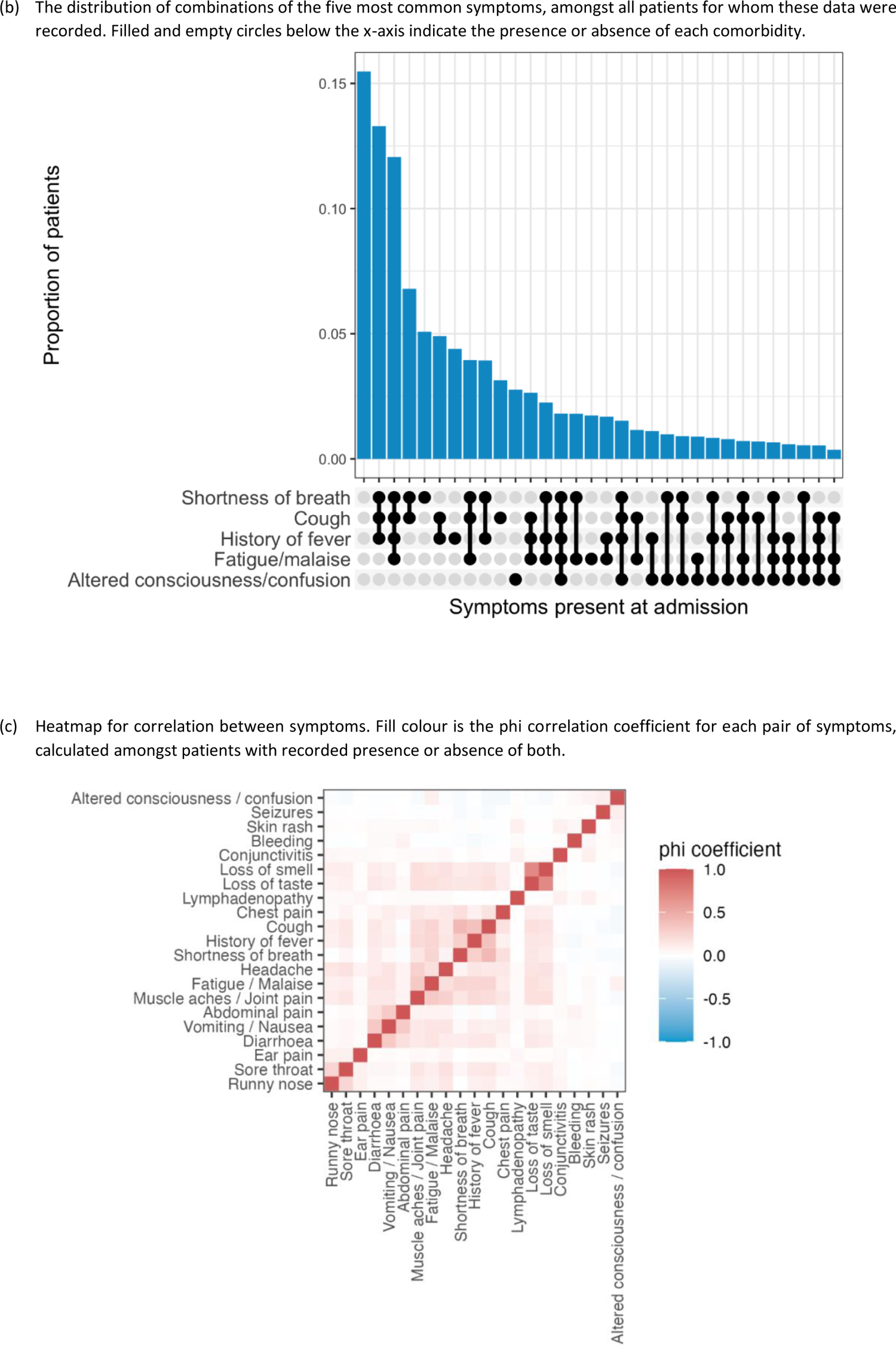
Clinical symptoms of patients at admission

Clusters of symptoms are presented in Figure 6b and Figure 6c. The most common symptom combination was shortness of breath, cough, and history of fever.

The observed symptoms on admission partly represented case definitions and policies for hospital admission, which may change as time passes. The overall percentages of patients meeting the four most common case definitions were 77%, 84%, 77%, 61% for CDC, ECDC, PHE, and WHO, respectively, and varies by age (Figure 7).

**Figure 7:**
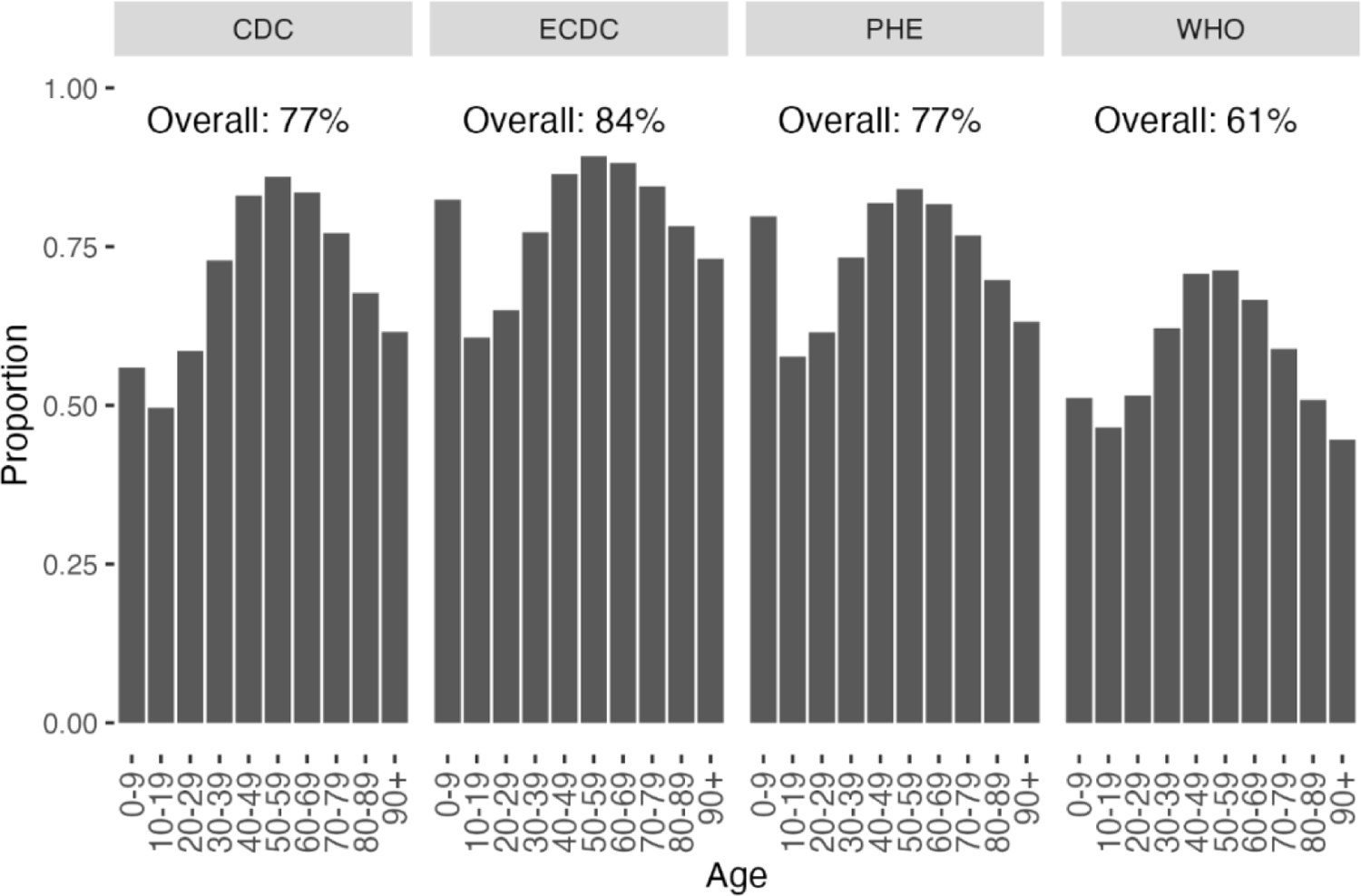
Proportion of patients that meet the 4 most common COVID-19 symptom case definitions by age

The frequency of presenting symptoms varied with age (Figure 8). Altered consciousness increased with age. The proportions of fever, respiratory and constitutional symptoms were higher in patients aged 40-60. Cough or fever were also common in children aged 0-9. The proportion of abdominal pain, diarrhoea, and vomiting were less age-dependent and similar across age groups.

**Figure 8:**
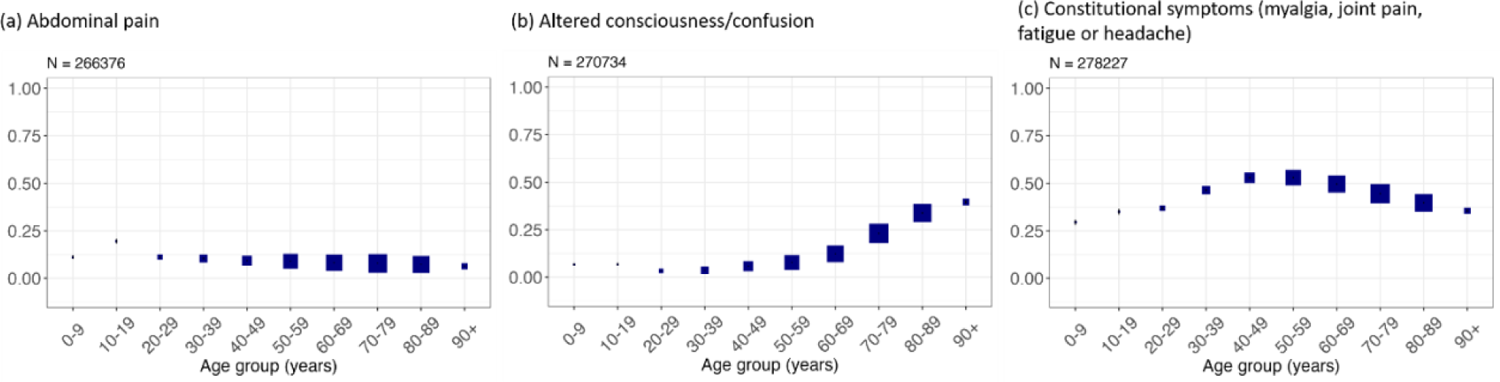
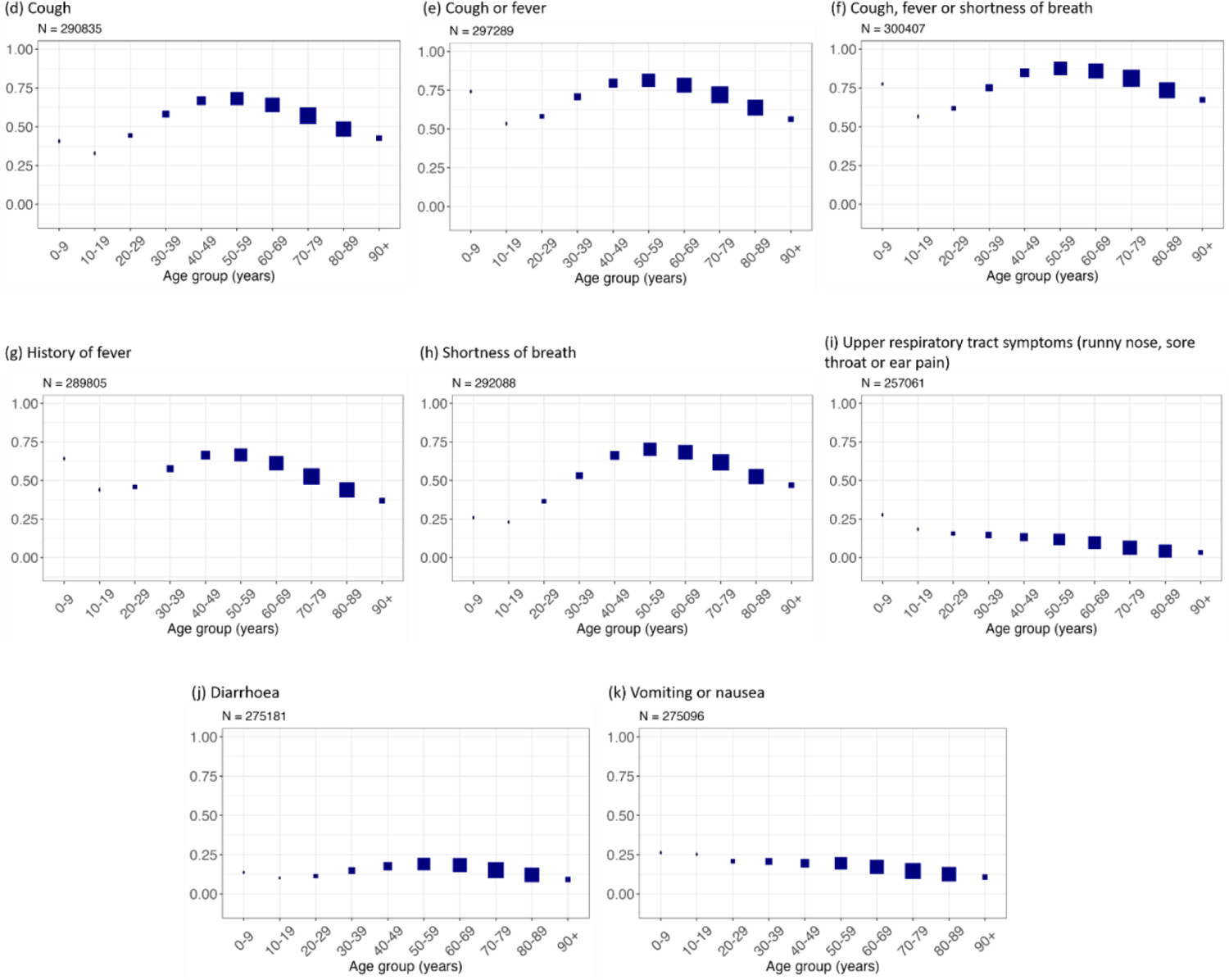
Symptoms recorded at hospital presentation stratified by age group. Boxes show the proportion of individuals with each symptom. The size of each box is proportional to the number of individuals represented. N is the number of individuals included in the plot (this varies between plots due to data completeness. The numbers for panel c, e, f, i were patients who had information on at least one symptom).

In Figure 9, we present the frequencies of the most common symptoms varied by regions. Each panel corresponds to a different region, and within region, symptoms that were present more often correspond to higher y-axis coordinates. To improve visualization, transparency decreases with increasing frequency. Across all geographical regions, shortness of breath, fever, and cough were the three symptoms with the highest prevalence.

**Figure 9:**
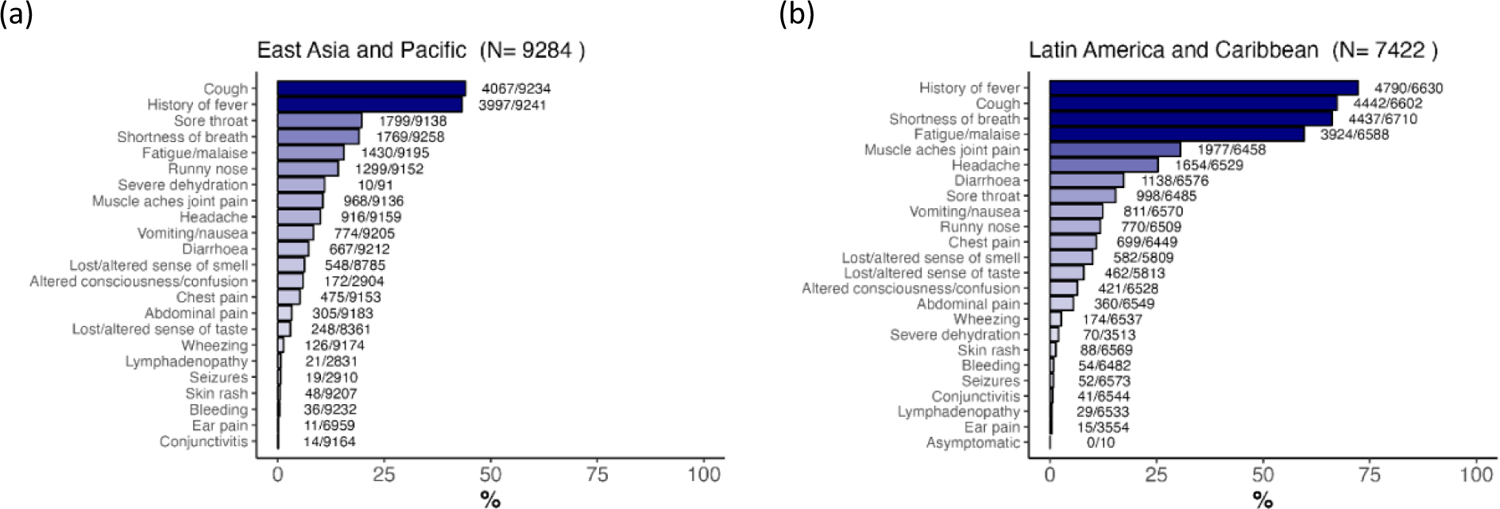
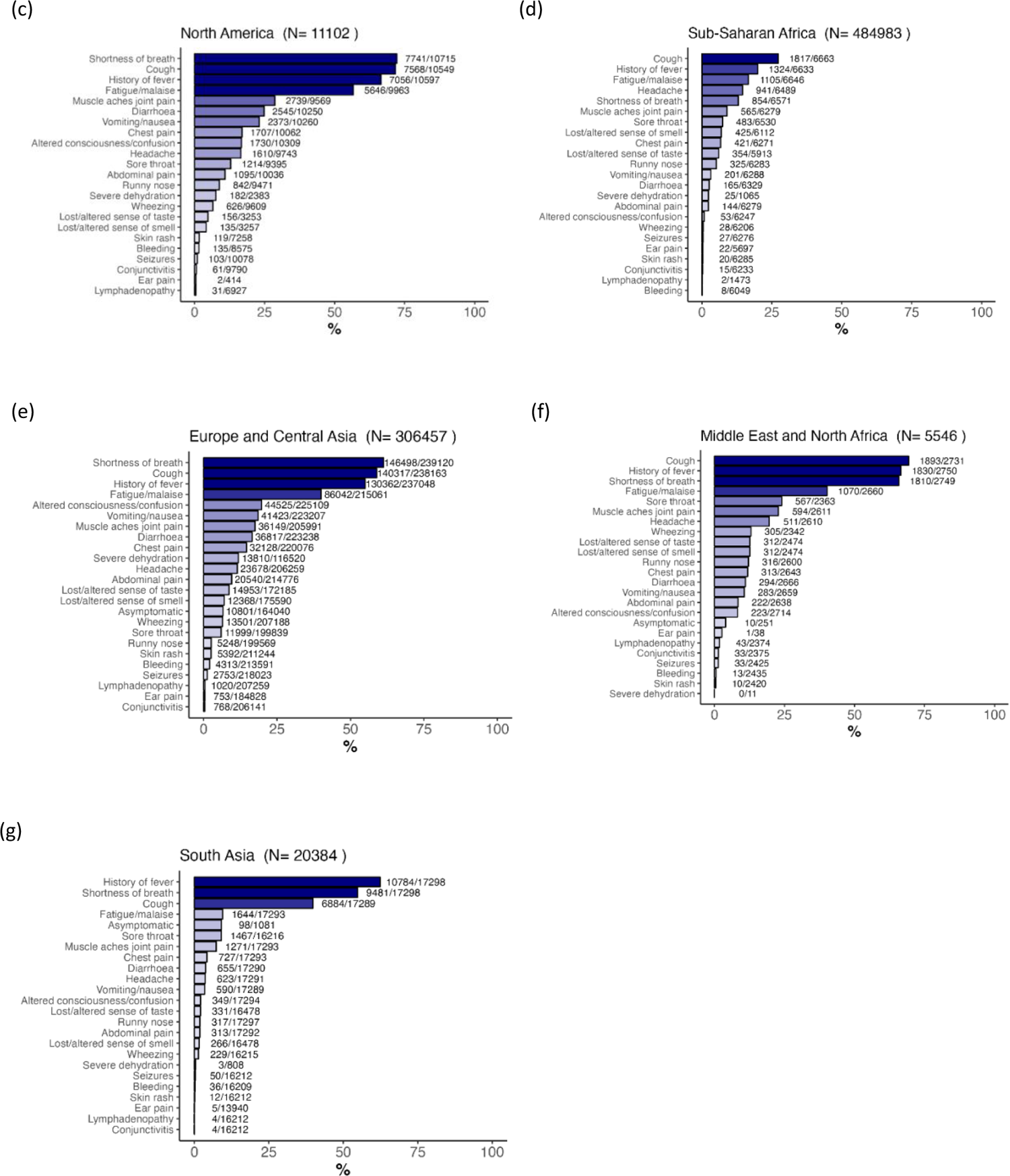
Symptoms recorded at hospital presentation stratified by region. The numbers next to each bar represent patients presented with this symptom/patients recorded. The N in the title represents the total number of patients in each region, including those with or without symptoms recorded.

### Comorbidities and other concomitant conditions at admission

Information was available on at least 75% of cases for diabetes, chronic pulmonary disease, chronic kidney disease, asthma, malignant neoplasm, cardiac chronic disease, HIV/AIDS, and hypertension (Figure 10).

**Figure 10:**
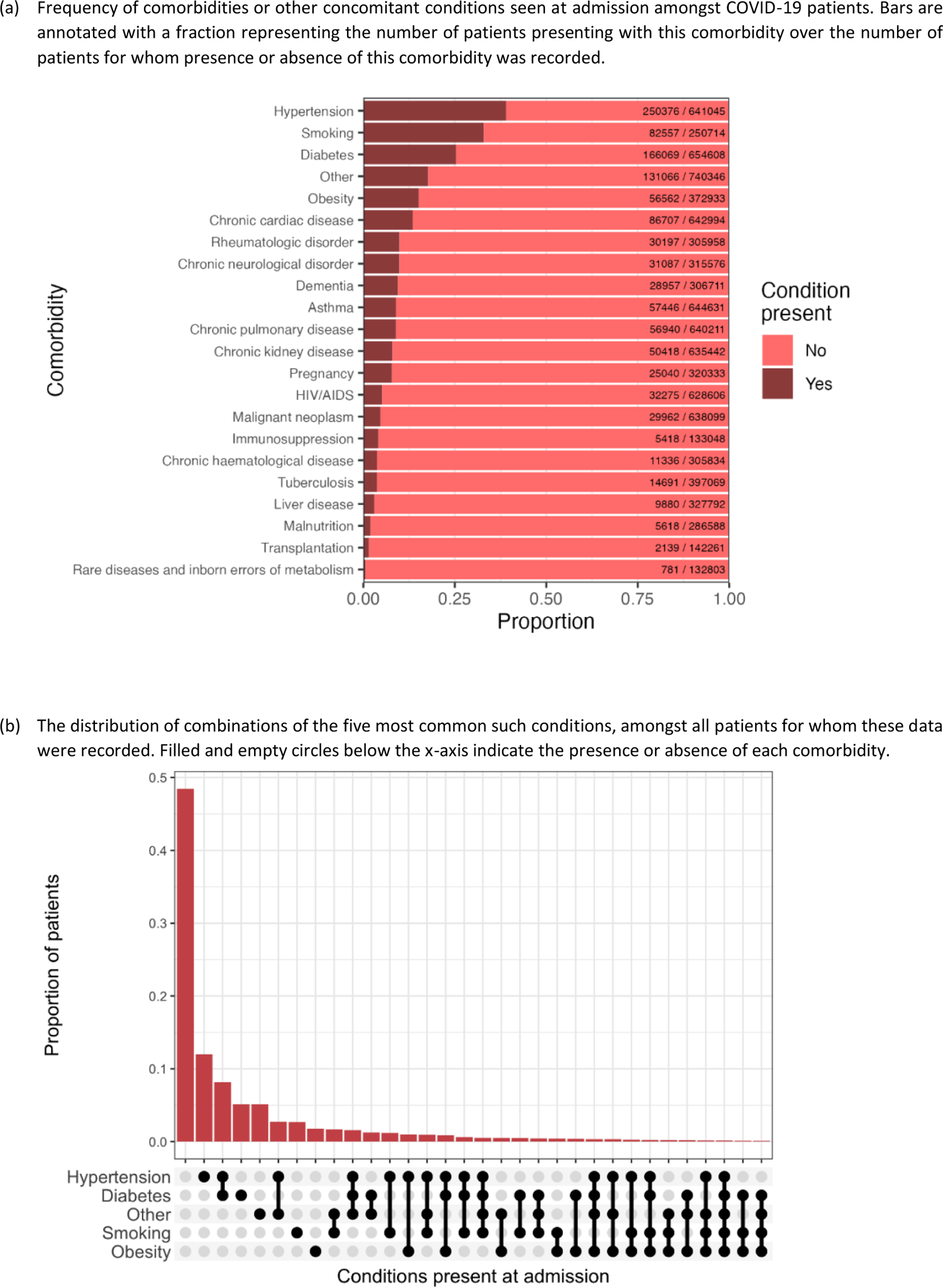
Comorbidities and other concomitant conditions

Conditions presented in at least 10% of cases were hypertension (40% of those reported), smoking (33%), diabetes (25%), obesity (15%), chronic cardiac disease (13%) (Figure 10a). Combinations of the five most prevalent conditions are presented in Figure 10b.

The age distribution of the most frequent concomitant conditions is presented in Figure 11. Dementia and hypertension increased with age.

**Figure 11:**
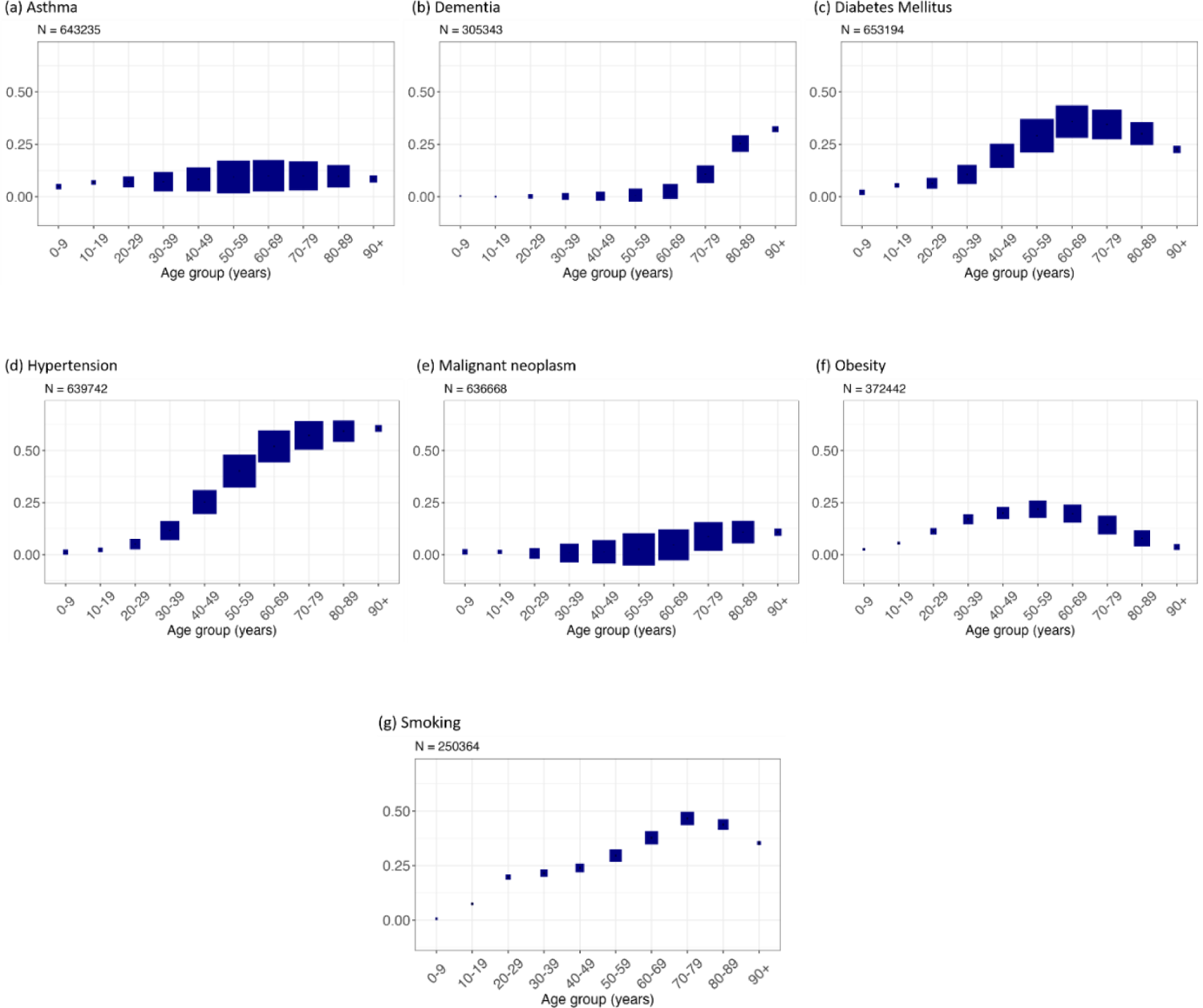
Comorbidities recorded at hospital presentation stratified by age group. Boxes show the proportion of individuals with each symptom. The size of each box is proportional to the number of individuals represented. N is the number of individuals included in the plot (this varies between plots due to data completeness).

In Figure 12, the frequencies of most common comorbidities in patients by regions are presented. Each panel corresponds to a different region, and within region, more frequent comorbidities correspond to higher y-axis coordinates. To improve visualization, transparency decreases with increasing frequency. The most common comorbidities were hypertension, diabetes, obesity, chronic cardiac disease, smoking, which are similar across regions.

**Figure 12:**
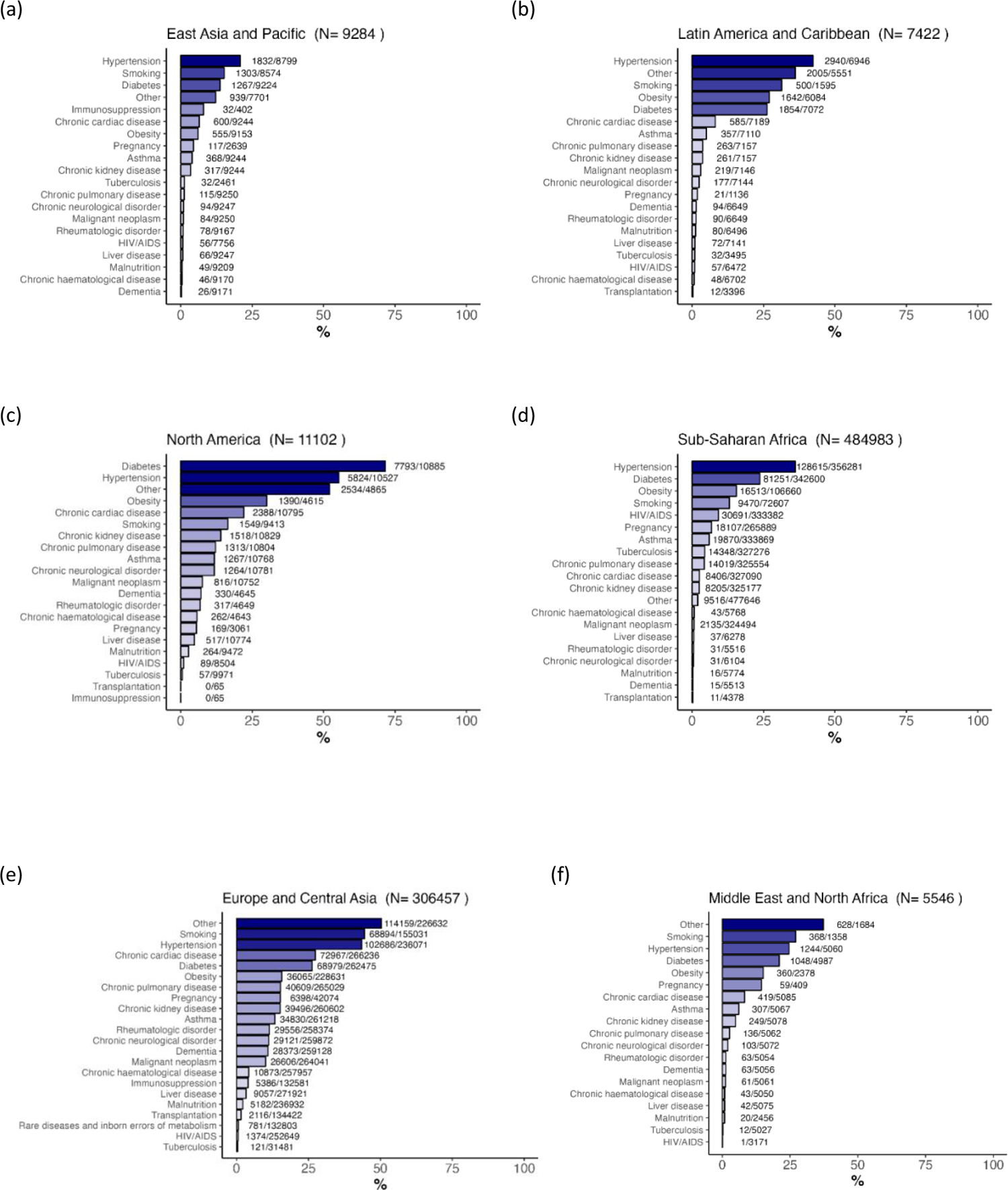
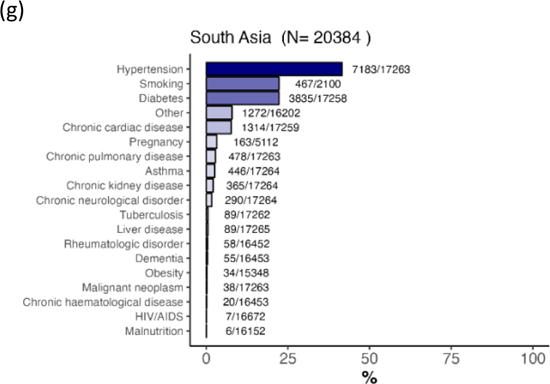
Comorbidities recorded at hospital presentation stratified by region. The numbers next to each bar represent patients presented with this symptom/patients recorded. The N in the title represents the total number of patients in each region, including those with or without symptoms recorded.

### Vital signs

Oxygen saturation was recorded for 286853 patients at admission. Of these, 32.7% were on oxygen therapy, and 67.3% were not. At admission, the median oxygen saturation of those who were on oxygen therapy was 95%, whereas the median was 96% for those who were not on oxygen therapy (Figure 13 provides a breakdown of oxygen saturation by age and oxygen therapy status). Among those who were on oxygen therapy at admission (N = 93843), 34.7% had oxygen saturation <94, and among those who were not on oxygen therapy at admission (N = 193010), 24.1% had oxygen saturation <94. All vital signs are presented in Figure 13 by age.

**Figure 13:**
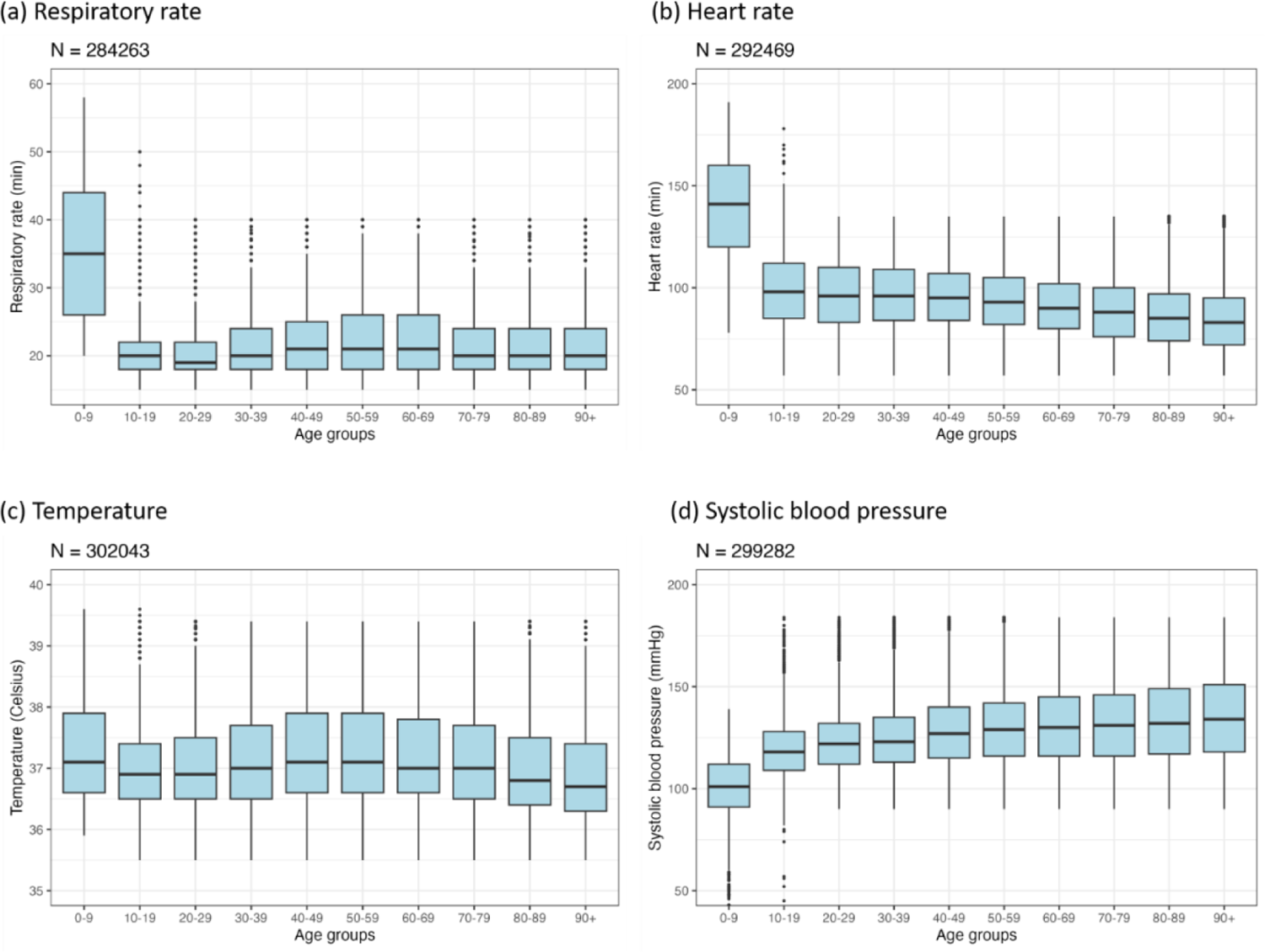
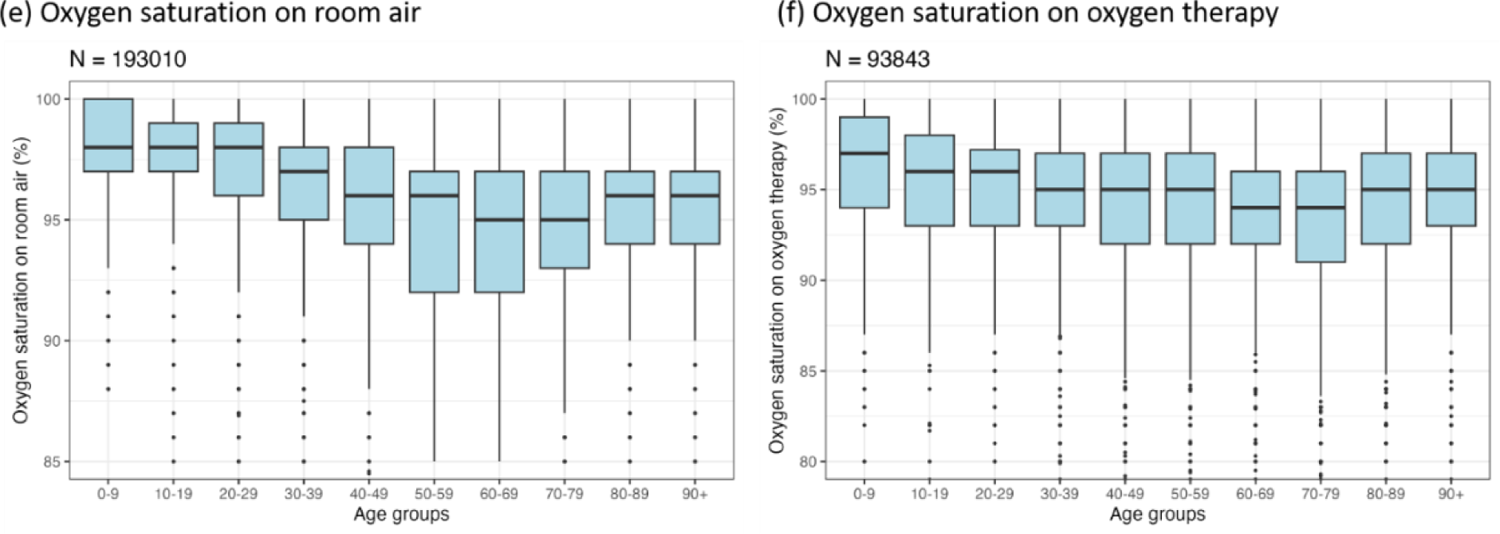
Box and whisker plots for observations at hospital presentation stratified by age group. N is the number of individuals included in the plot (this varies between plots due to data completeness).

### Laboratory values

Laboratory tests results were available for a minority of patients. They are summarised in Figure 14.

**Figure 14:**
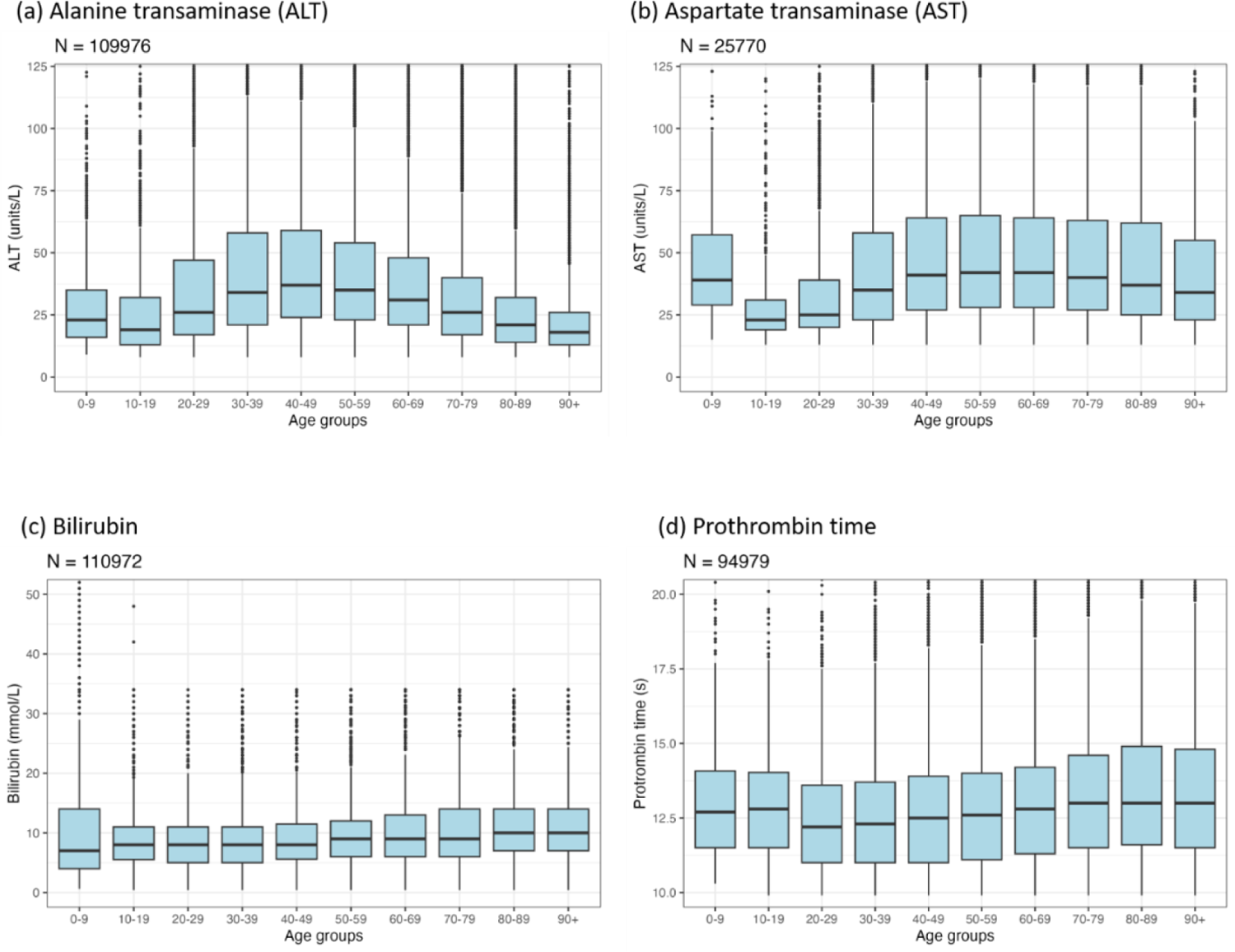
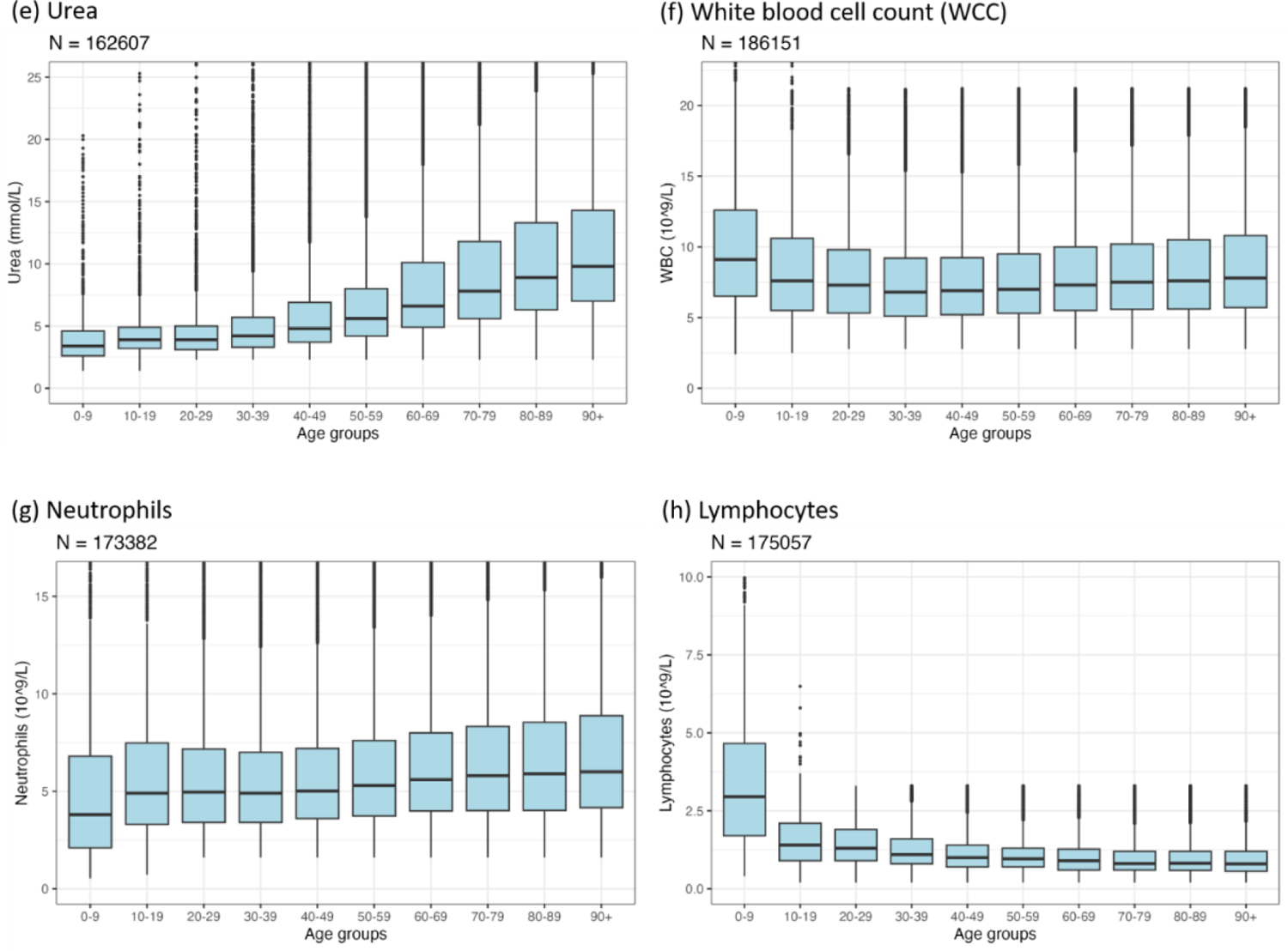
Box and whisker plots for laboratory results within 24 hours of hospital presentation stratified by age group. N is the number of individuals included in the plot (this varies between plots due to data completeness).

### Treatment

Antibiotics were given to 298349/802241 (37.2%) patients, and 75202/801537 (9.4%) received antivirals. These treatment categories were not mutually exclusive since some patients received multiple treatments - the denominators differ due to data completeness. Of 717419 patients with data available on oxygen supplementation within 24 hours from admission, 206247 (28.7%) received supplemental oxygen and 511172 (71.3%) did not. Throughout their stay in the hospital, 421580/824444 (51.1%) patients received some degree of oxygen supplementation; of these, 56437/553792 (10.2%) received non-invasive ventilation (NIV) and 74592/818428 (9.1%) IMV.

Corticosteroids were administered to 205853/809043 (25.4%) patients. This includes 34982/72276 (48.4%) of those who received IMV, 41849/54107 (77.3%) of those who had oxygen therapy but not IMV, and 31019/391729 (7.9%) of those who had no oxygen therapy. On 16 June 2020, results for dexamethasone were released from the RECOVERY randomized controlled trial [20,21]. This trial found that dexamethasone reduced deaths for patients receiving IMV and oxygen therapy, but not among patients not receiving respiratory support. Of patients admitted since 16 June 2020, corticosteroids were given to 27535/56216 (49%) of those who received IMV, and 28014/350016 (8%) of those with no oxygen therapy.

**Figure 15:**
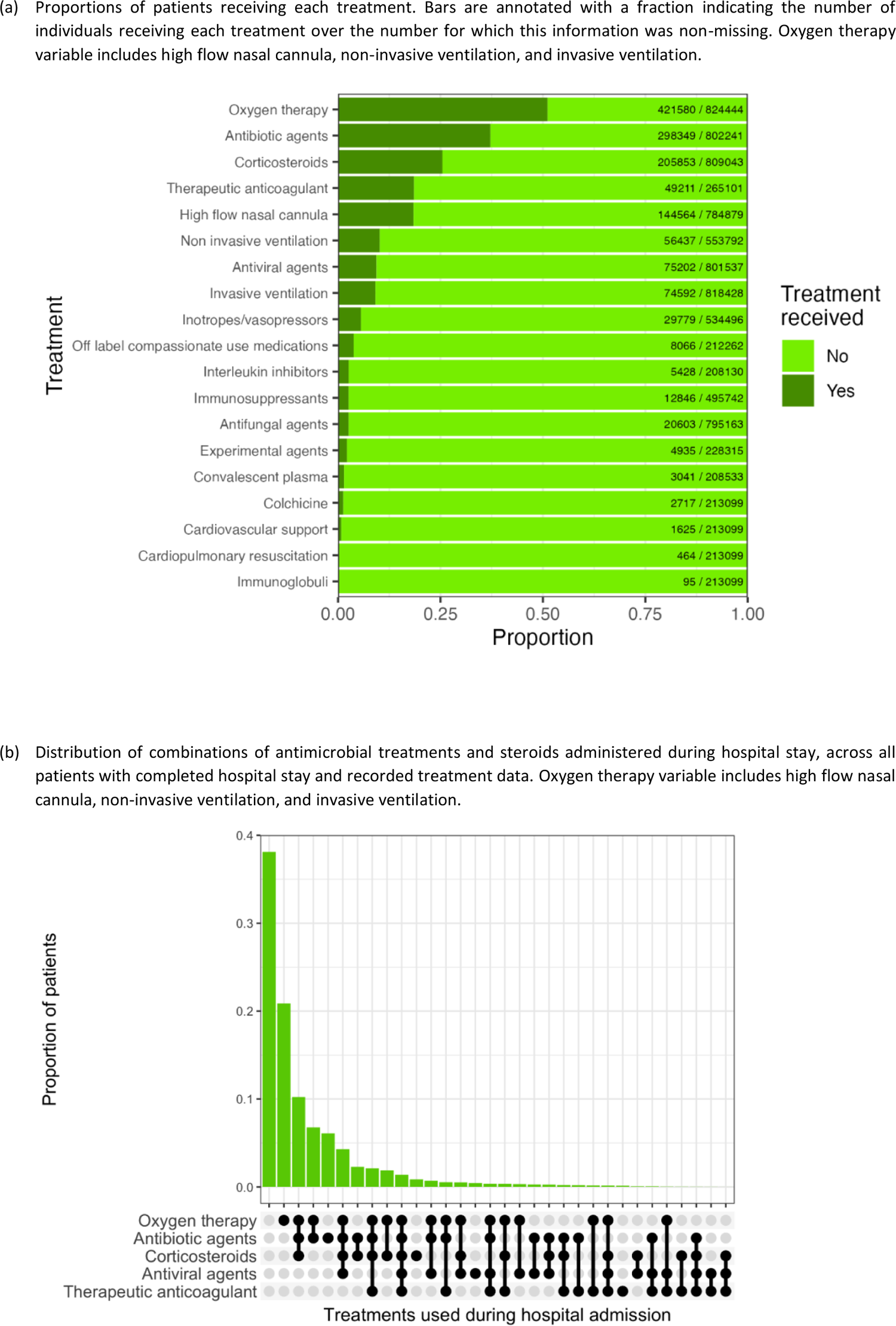
Treatments in the entire population

### Intensive Care and High Dependency Unit Treatments

Of the 845291 patients with data on ward admissions available, a total of 129672 (15.3%) were admitted at some point of their illness into an intensive care unit (ICU) or high dependency unit (HDU), among which, 39040 (53.6%) on the day of hospital admission (56879 patients without ICU admission date).

The range of treatments received whilst in ICU/HDU is presented in Figure 16a and Figure 16b. Of the patients admitted into ICU/HDU, 58375/64669 (90.3%) received antibiotics and 27314/64389 (42.4%) antivirals. 66618/69296 (96.1%) received some degree of oxygen supplementation, of which, 32753/68196 (48%) received NIV and 41672/67910 (61.4%) IMV.

**Figure 16:**
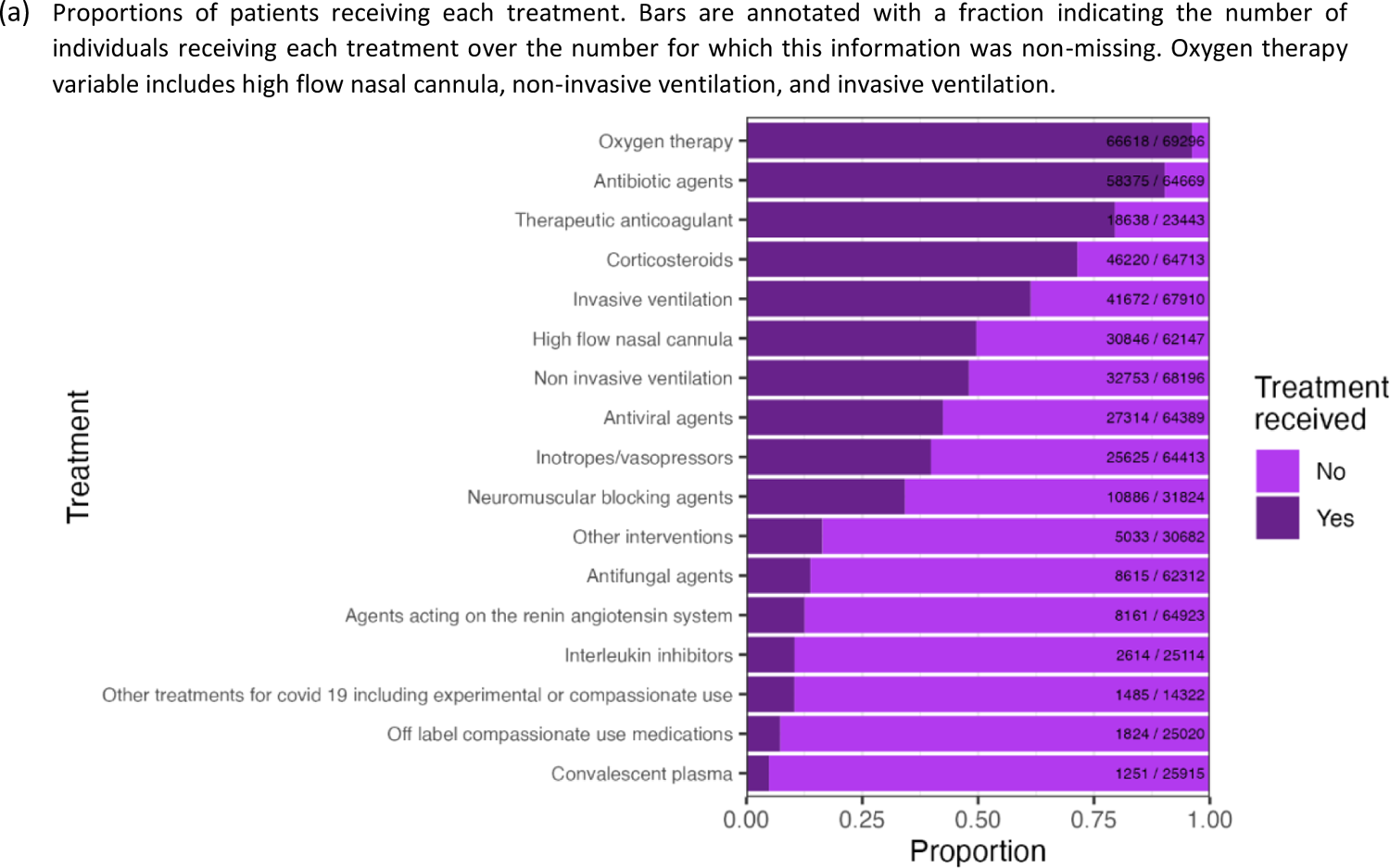
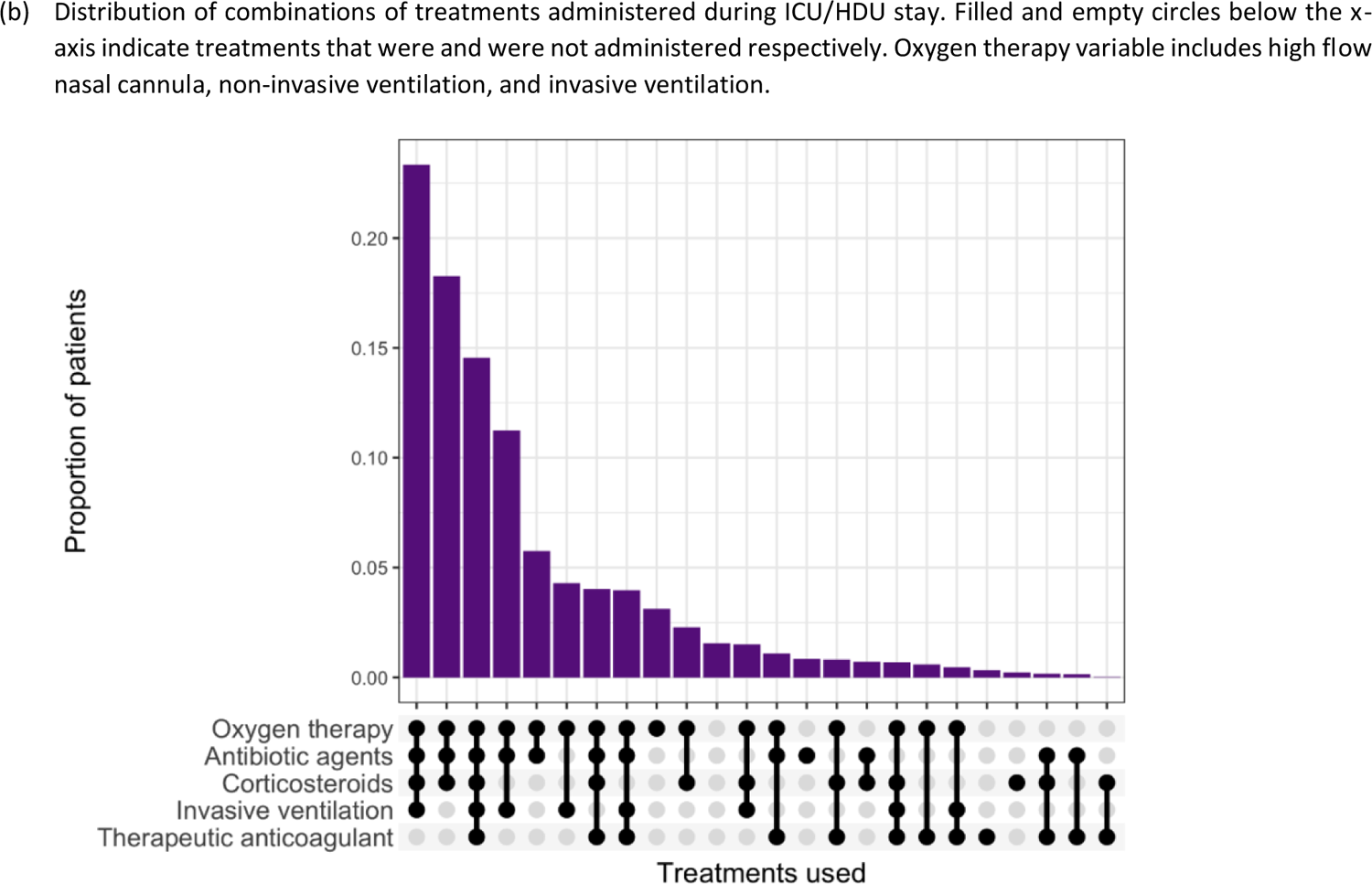
Treatments in the ICU/HDU population

A total of 56437 patients received non-invasive mechanical ventilation (NIV). The mean and median number of days from admission to receiving NIV were 2.2 days and 1 days respectively (SD: 3.6 days) – estimated from records on cases with complete records on dates of hospital admission and NIV onset (N = 36307). The mean and median duration for NIV were 3.9 days and 3 days respectively (SD: 3.5 days) – estimated based on only those cases which have completed NIV duration records (N = 11661).

A total of 74592 patients received invasive mechanical ventilation (IMV). The mean and median duration from admission to receiving IMV were 3.4 days and 1 days respectively (SD: 4.7 days) – estimated from records on cases with complete records on dates of hospital admission and IMV onset (N = 27003). The mean, median and SD for the duration of IMV – estimated based on all 41046 cases with complete records on IMV stays – were 10.1 days, 7 days and 9.8 days respectively.

### Key time variables

The key time variables are presented in Table 3. Patients tend to come to the hospital within the first week of their illness with a mean of 5.4 days and a median of 5 days from symptoms onset. 53.4% of ICU/HDU admissions occurred within the first day at the hospital. The mean and median duration of hospital stay were 9.6 and 7 days, respectively. The duration of stay in ICU/HDU had a mean of 10.2 days and a median of 7 (SD: 10.3 days) – estimated on only those cases with complete records for ICU/HDU duration or ICU/HDU start/end dates (N = 71188).

**Table 3:**
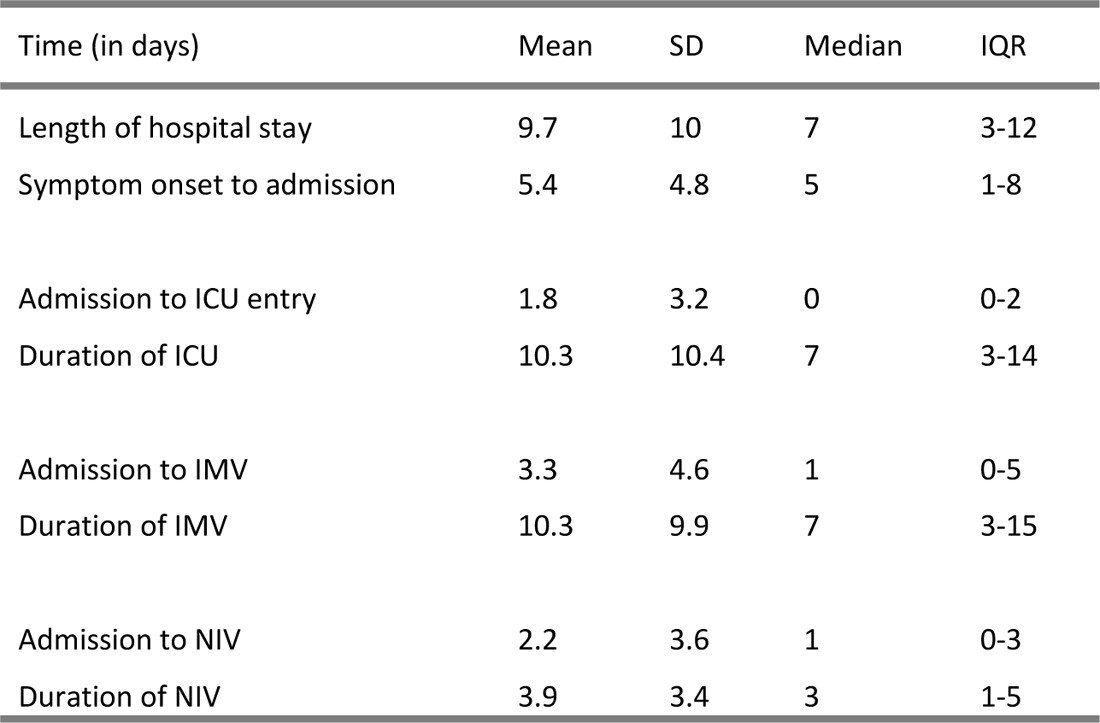
Key time variables. SD: Standard deviation; IQR: Interquartile range. Outliers (values greater than 120) were excluded prior to the computation of estimates.

| Time (in days) | Mean | SD | Median | IQR |
| --- | --- | --- | --- | --- |
| Length of hospital stay | 9.7 | 10 | 7 | 3-12 |
| Symptom onset to admission | 5.4 | 4.8 | 5 | 1-8 |
| Admission to ICU entry | 1.8 | 3.2 | 0 | 0-2 |
| Duration of ICU | 10.3 | 10.4 | 7 | 3-14 |
| Admission to IMV | 3.3 | 4.6 | 1 | 0-5 |
| Duration of IMV | 10.3 | 9.9 | 7 | 3-15 |
| Admission to NIV | 2.2 | 3.6 | 1 | 0-3 |
| Duration of NIV | 3.9 | 3.4 | 3 | 1-5 |

**Figure 17:**
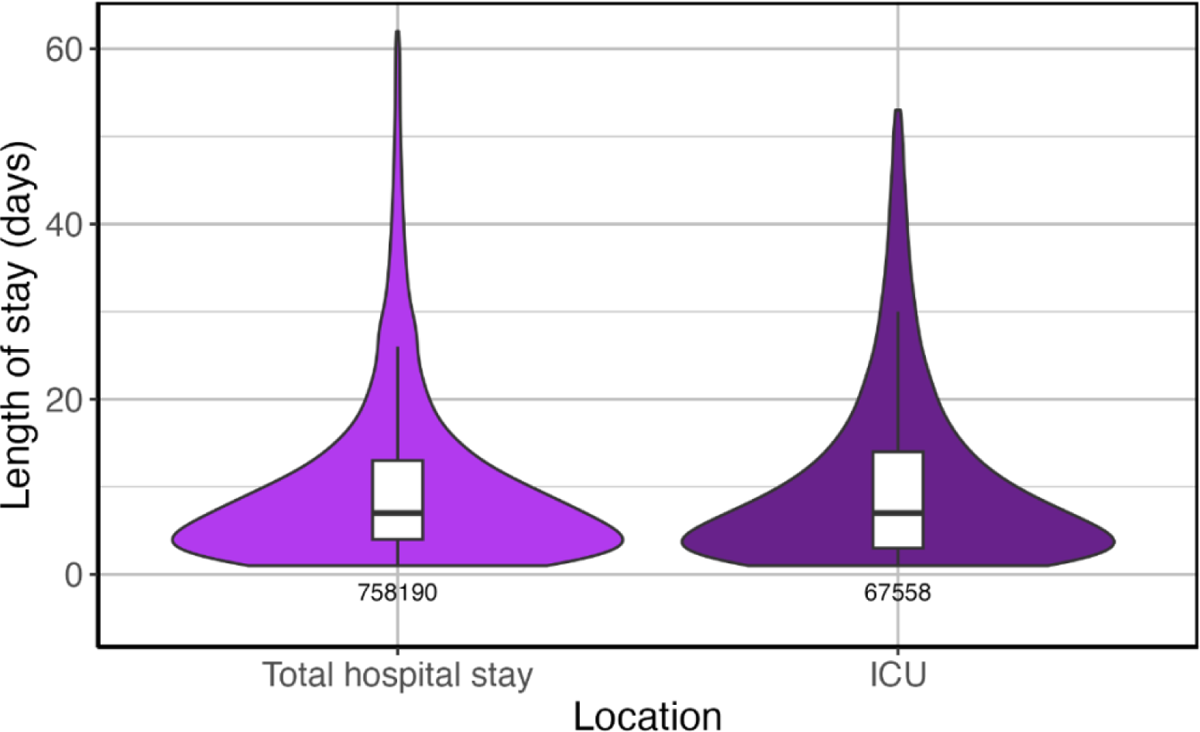
Distribution of lengths of stay for patients who were admitted to ICU/HDU: total length of stay for this group and length of stay within intensive care. This only includes cases with reported completed stays. The coloured areas indicate the kernel probability density of the observed data and the box plots show the median and interquartile range of the variable of interest.

The distribution of the number of days from admission to ICU admission is shown in Figure 18.

**Figure 18:**
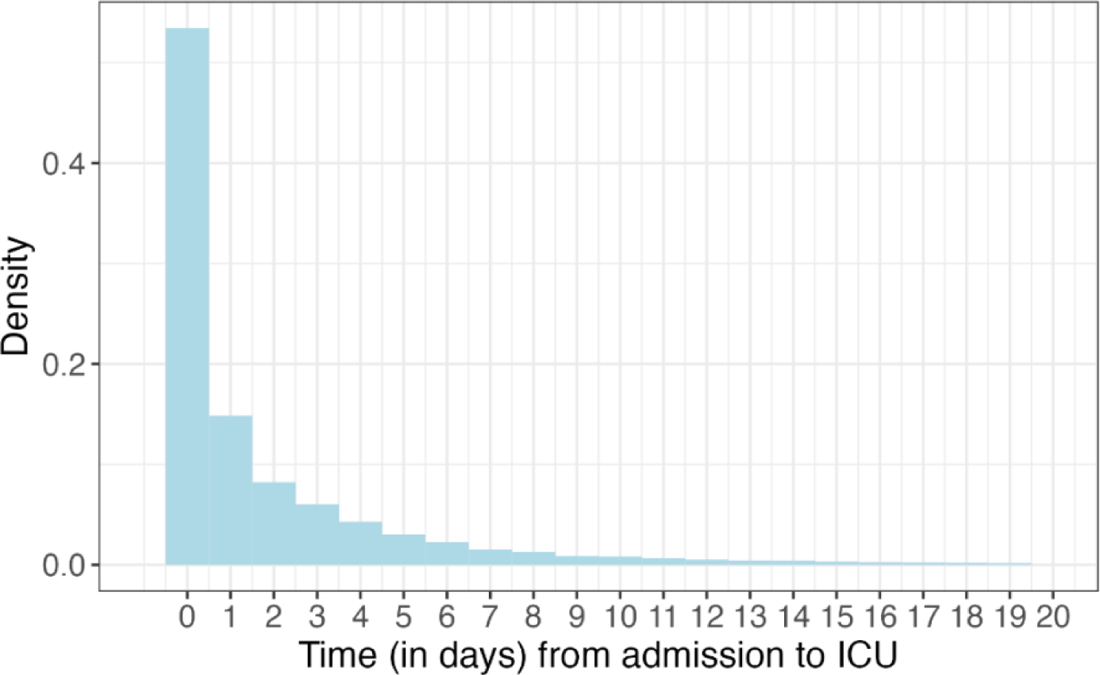
Distribution of time (in days) from hospital admission to ICU admission. The figure displays data on only those cases with a reported ICU start date.

The observed mean duration for the number of days from hospital admission to outcome (death or discharge) was 9.4, with SD 9.5 days and a median of 7 days. These estimates were based on all cases which have completed records on length of hospital stay (N = 746709). The distributions of length of hospital stay are presented in Figure 19a according to sex and Figure 19b by patient age group.

**Figure 19:**
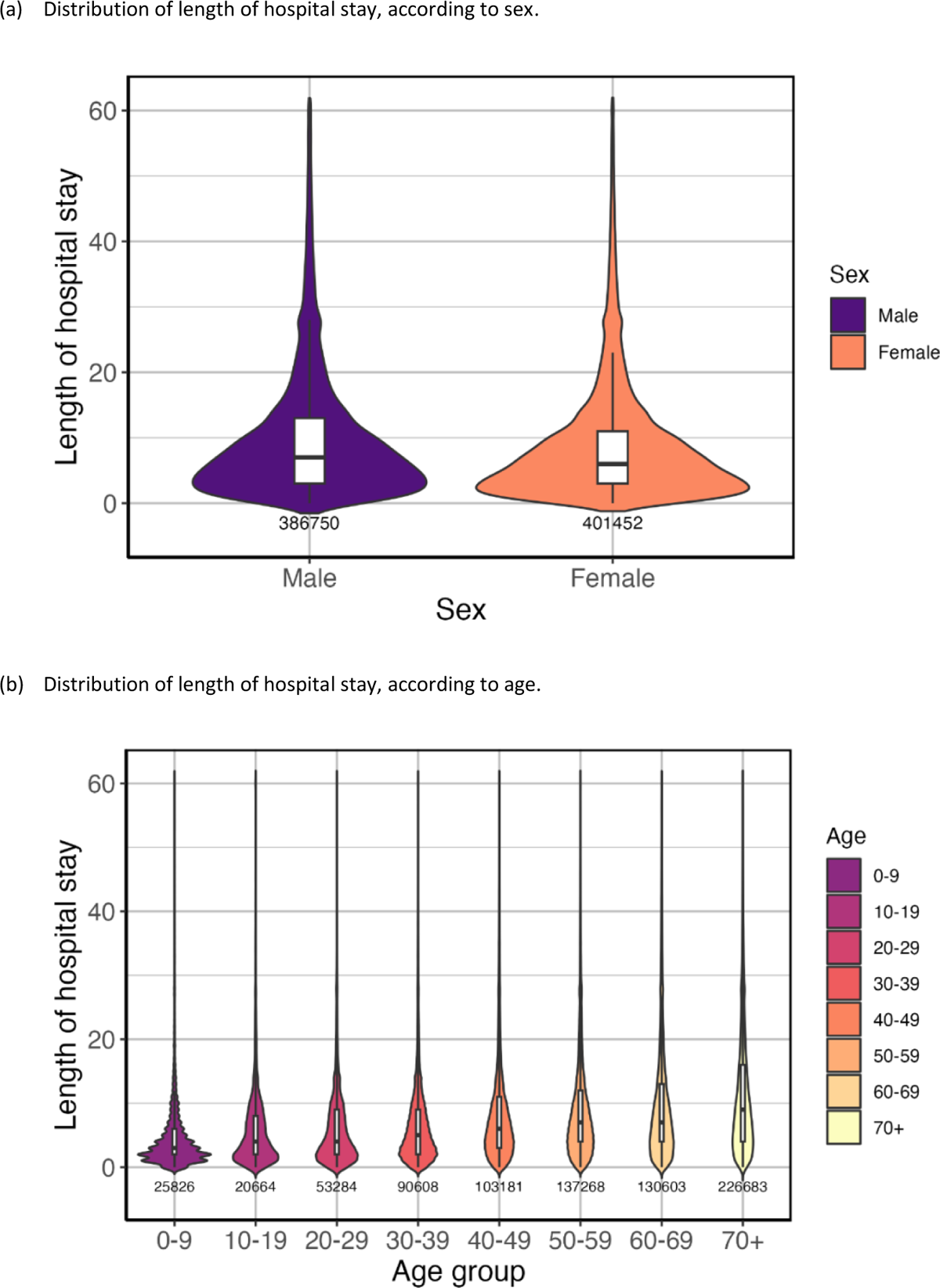
Distribution of length of hospital stay. This only includes cases with reported outcomes. The coloured areas indicate the kernel probability density of the observed data and the box plots show the median and interquartile range of the variable of interest.

### Outcomes

Outcomes were recorded for 762728 patients (90.2%), consisting of 595298 recoveries and 167430 deaths. Outcome records were unavailable for 82563 patients.

**Figure 20:**
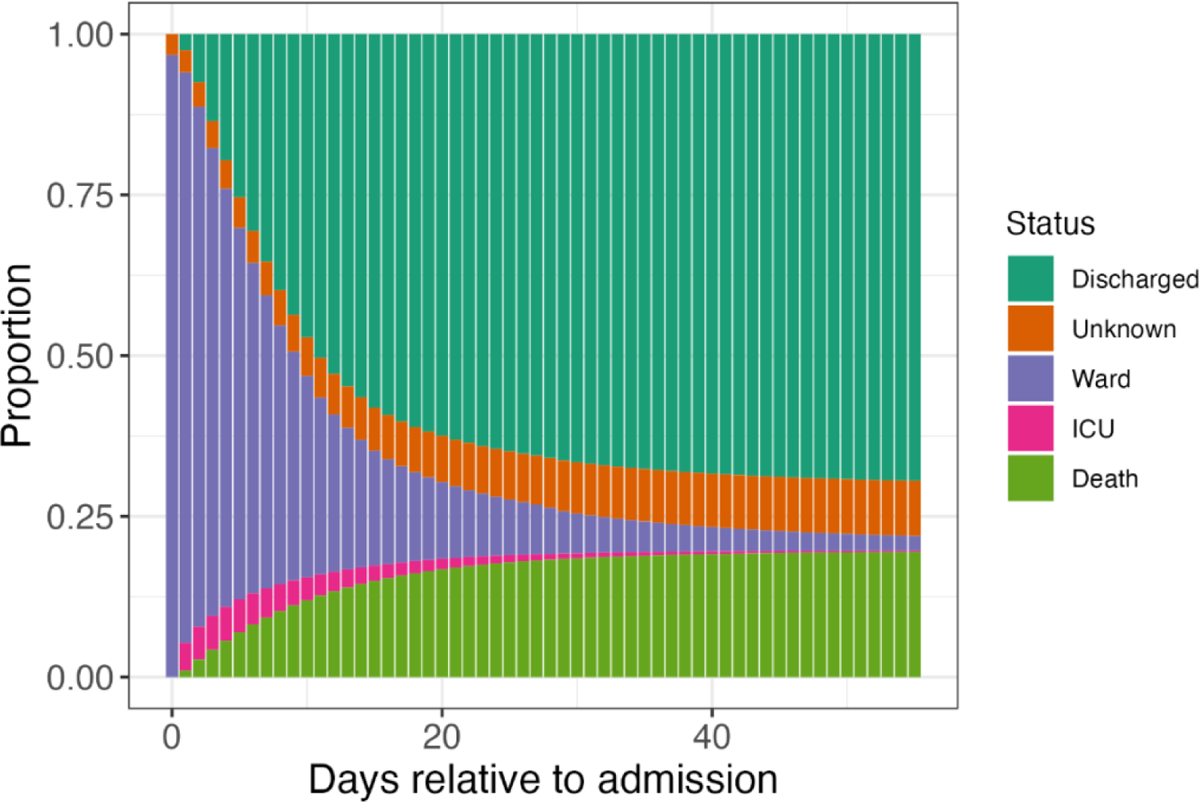
The distribution of patient status by number of days after admission. Patients with ‘unknown’ status have left the site at the time of report but have unknown outcomes due to missing data.

**Figure 21:**
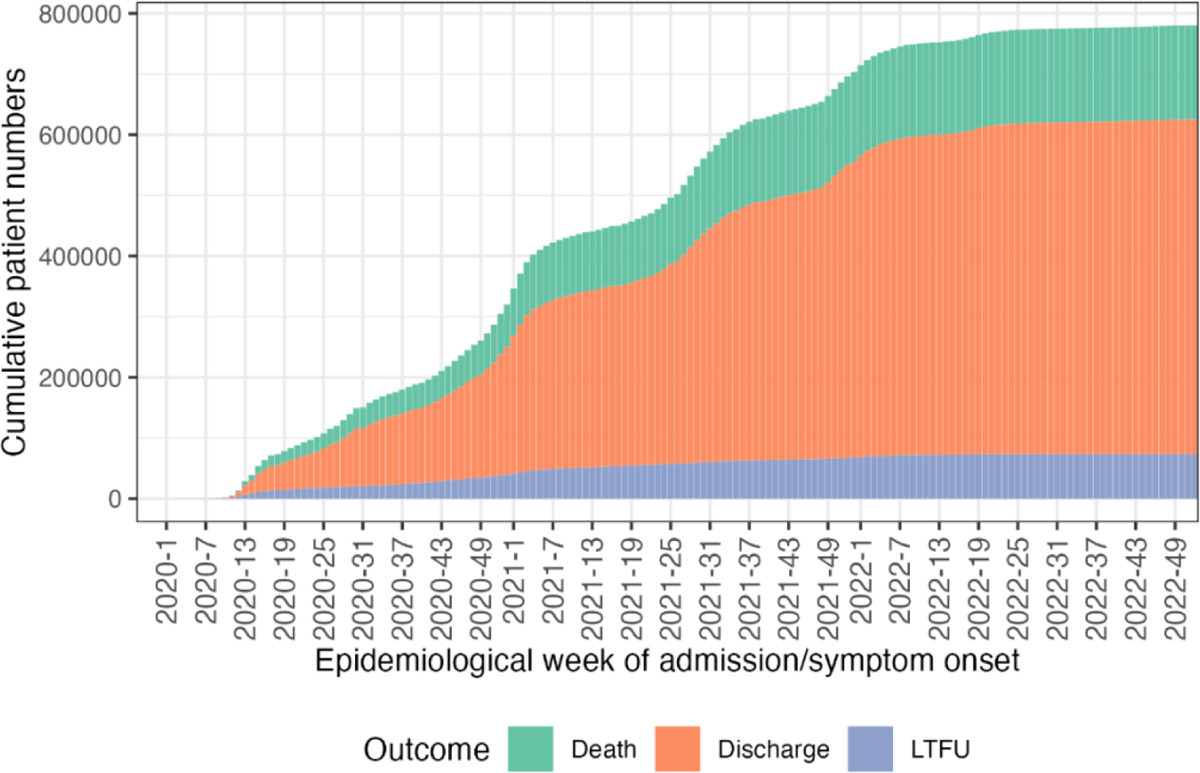
Cumulative patient numbers and outcomes by epidemiological week of admission (or, for patients infected in hospital, of symptom onset).

The overall estimated case fatality ratio (CFR) was 22% (95% CI 21.9-22) and was 35.9% (95% CI 35.6-36.1) for patients admitted to ICU/HDU - calculated out of those with data on outcome.

## Discussion and conclusions

COVID-19 has tested the effectiveness of ISARIC’s operating model: a global, open-source, collaborative approach set up 10 years ago to prevent illness and deaths from infectious disease outbreaks. In January 2020, ISARIC launched its COVID-19 Clinical Characterisation Protocol (CCP) and Case Report Form (CRF) as well as a free data management platform for researchers to upload their clinical data, globally. The ISARIC COVID-19 CCP database was open and publicly accessible from late January 2020, when less than a thousand COVID-19 cases had been reported globally; by mid-February 2020, the first patient record was successfully uploaded on to the platform; by mid-March 2020, the platform had ten thousand records.

Now, 3 years into the pandemic, with more than 945,000 records, the ISARIC clinical data platform has grown to become the largest international individual patient dataset of COVID-19 hospitalised cases. ISARIC is a model of global peer-to-peer collaboration, with contributions by 1807 sites in 76 countries, with similar representations of patients from high-income and low- and middle-income countries.

This report describes the features of COVID-19 patients in the entire cohort, made from cases accrued over 36 months with different contributions from various countries. Everyone can trace the evolution over time of the database through 18 previous versions of this report. In addition, specific questions regarding clinical presentations, risks factors for outcomes, and trends are addressed through ad-hoc analyses.

Partners can submit ad-hoc request to perform analyses, to access the 945,000 case-based dataset, by using our Statistical Analysis Plan form (SAP). Once a SAP has been submitted, our statistical team assesses the plan for quality of research and feasibility of data analysis. Then, our clinical team provides feedback on clinical relevance and overlap with other analyses. Once this process has been completed, the SAP is sent to all ISARIC collaborators for the opportunity to review. This initial peer review process ensures that research is of the utmost quality and it helps encourage a truly participatory approach.

All ISARIC contributors are invited to participate in analyses through review and input on the SAP and the resulting publication. The outputs of our joint work are disseminated as widely as possible to inform patient care and public health policy. ISARIC aims to include the names of all those who contribute data in the cited authorship of publications, subject to the submission of contact details and confirmation of acceptance of the final manuscript within the required timelines, per ICMJE policies and the ISARIC publication policy.

### What have our partners achieved

Using the ISARIC and ISARIC4C databases, our partners have produced a broad range of evidence. More than 100 reports and manuscripts have been published to date, and many more are in progress.

True international collaboration during a pandemic is key to putting data into action. ISARIC partners have achieved this objective, and the results are shown in this final report of the COVID-19 Clinical Database.

## Data Availability

We welcome applications for access to data on the COVID-19 Data Sharing Platform via the Data Access Committee (www.iddo.org/covid-19).

## Acknowledgement

We are grateful for the expertise and efforts of these individuals, listed at the end of this report. For further details and access to the resources listed above, go to www.isaric.org

## Contributing individuals

Moharam S. A, Sabriya Abdalasalam, Alaa Abdalfattah Abdalhadi, Naana Reyam Abdalla, Walaa Abdalla, Almthani Hamza Abdalrheem, Ashraf Abdalsalam, Saedah Abdeewi, Esraa Hassan Abdelgaum, Mohamed Abdelhalim, Mohammed Abdelkabir, Israa Abdelrahman, Sheryl Ann Abdukahil, Lamees Adil Abdulbaqi, Salaheddin Abdulhamid, Widyan Abdulhamid, Nurul Najmee Abdulkadir, Eman Abdulwahed, Rawad Abdunabi, Ryuzo Abe, Laurent Abel, Ahmed Mohammed Abodina, Khaled Abouelmagd, Amal Abrous, Lara Absil, Kamal Abu Jabal, Nashat Abu Salah, Salsabeel M. A. Abukhalaf, Abdurraouf Abusalama, Tareg Abdallah Abuzaid, Subhash Acharya, Andrew Acker, Shingo Adachi, Elisabeth Adam, Safia Adem, Manuella Ademnou, Francisca Adewhajah, Enrico Adriano, Diana Adrião, Samuel Yaw Adu, Anthony Afum-Adjei Awuah, Melvin Agbogbatey, Saleh Al Ageel, Aya Mustafa Ahmed, Musaab Mohammed Ahmed, Shakeel Ahmed, Zainab Ahmed Alaraji, Abdulrahman Ahmed Elhefnawy Enan, Reham Abdelhamid Ahmed Khalil, Ali Mostafa Ahmed Mohamed Abdelaziz, Marina Aiello, Kate Ainscough, Eka Airlangga, Tharwat Aisa, Bugila Aisha, Ali Aisha, Ali Ait Hssain, Ali Ait Hssain, Younes Ait Tamlihat, Takako Akimoto, Ernita Akmal, Chika Akwani, Eman Al Qasim, Mohammed Al-Aquily, Abdulrahman Al-Fares, Yousef Al-Saba’a, Tala Al-dabbous, Yasmina Alaa, Ahmed Alajeeli, Ahmed Alali, Razi Alalqam, Aliya Mohammed Alameen, Zinah A. Alaraji, Khalid Albakry, Safa Albatni, Angela Alberti, Osama Aldabbourosama, Amer Aldhalia, Abdulkarim Aldoukali, Senthilkumar Alegesan, Cynthia Alegre, Marta Alessi, Beatrice Alex, Kévin Alexandre, Asil Alflite, Huda Alfoudri, Huda Alfoudri, Khadeejeh M. A. Alfroukh, Qamrah Alhadad, Hoda Salem Alhaddad, Maali Khalid Mohamed Abdalla Alhasan, Ahmad Nabil Alhouri, Hasan Alhouri, Maha TagElser Mohammed Ali, Imran Ali, Adam Ali, Syed Ali Abbas, Yomna Ali Abdelghafar, Naseem Ali Shah, Naseem Ali Sheikh, Kazali Enagnon Alidjnou, Mahmoud Aljadi, Sarah Aljamal, Mohammed Alkahlout, Khalid Jehad Khalid Alkaraki, Akram Alkaseek, Qabas Alkhafajee, Clotilde Allavena, Nathalie Allou, Lana Almasri, Abdulrahman Almjersah, Raja Ahmed Alqandouz, Walaa Alrfaea, Moayad Alrifaee, Rawan Alsaadi, Entisar Alshareea, Eslam Alshenawy, Aneela Altaf, João Alves, Rita Alves, João Melo Alves, Joana Alves Cabrita, Maria Amaral, Amro Essam Amer, Nur Amira, Heidi Ammerlaan, Amos Amoako Adusei, John Amuasi, Roberto Andini, Margarita Andreeva, Claire Andrejak, Andrea Angheben, François Angoulvant, Sophia Ankrah, Séverine Ansart, Sivanesen Anthonidass, Massimo Antonelli, Carlos Alexandre Antunes de Brito, Kazi Rubayet Anwar, Ardiyan Apriyana, Yaseen Arabi, Irene Aragao, Francisco Arancibia, Carolline Araujo, Antonio Arcadipane, Patrick Archambault, Lukas Arenz, Jean-Benoît Arlet, Christel Arnold-Day, Ana Aroca, Rakesh Arora, Lovkesh Arora, Elise Artaud-Macari, Diptesh Aryal, Motohiro Asaki, Angel Asensio, Elizabeth A. Ashley, Muhammad Ashraf, Muhammad Sheharyar Ashraf, Abir Ben Ashur, Franklin Asiedu-Bekoe, Namra Asif, Mohammad Asim, Grace Assi, Jean Baptiste Assie, Amirul Asyraf, Fouda Atangana, Ahmed Atia, Minahel Atif, Asia Atif Abdelrhman Abdallahrs, Anika Atique, Moad Atlowly, AM Udara Lakshan Attanyake, Johann Auchabie, Hugues Aumaitre, Adrien Auvet, Sergey Avdeev, Abdelmalek Awad Ali Mohammed, Eyvind W. Axelsen, Ared Ayad, Ahmed Ayman Hassan Helmi, Laurène Azemar, Mohammed Azizeldin, Cecile Azoulay, Hakeem Babatunde, Benjamin Bach, Delphine Bachelet, Claudine Badr, Nadia Baig, John Kenneth Baillie, J Kevin Baird, Zinah Aqeel Abdulzahra Bairmani, Erica Bak, Erica Bak, Agamemnon Bakakos, Nazreen Abu Bakar, Hibah Bileid Bakeer, Ashraf Bakri, Andriy Bal, Mohanaprasanth Balakrishnan, Irene Bandoh, Firouzé Bani-Sadr, Renata Barbalho, Nicholas Yuri Barbosa, Wendy S. Barclay, Saef Umar Barnett, Michaela Barnikel, Helena Barrasa, Cleide Barrigoto, Marie Bartoli, Cheryl Bartone, Joaquín Baruch, Romain Basmaci, Muhammad Fadhli Hassin Basri, AbdAlkarim Batool, Denise Battaglini, Jules Bauer, Diego Fernando Bautista Rincon, Denisse Bazan Dow, Abigail Beane, Alexandra Bedossa, Ker Hong Bee, Husna Begum, Sylvie Behilill, Albertus Beishuizen, Aleksandr Beljantsev, David Bellemare, Anna Beltrame, Beatriz Amorim Beltrão, Marine Beluze, Nicolas Benech, Lionel Eric Benjiman, Suzanne Bennett, Luís Bento, Jan-Erik Berdal, Lamis Berdeweel, Delphine Bergeaud, Hazel Bergin, José Luis Bernal Sobrino, Kikaire Bernard, Giulia Bertoli, Lorenzo Bertolino, Simon Bessis, Adam Betz, Sybille Bevilcaqua, Karine Bezulier, Amar Bhatt, Krishna Bhavsar, Isabella Bianchi, Claudia Bianco, Sandra Bichoka, Farah Nadiah Bidin, Felwa Bin Humaid, Mohd Nazlin Bin Kamarudin, Muhannud Binnawara, Zeno Bisoffi, Patrick Biston, Laurent Bitker, Mustapha Bittaye, Jonathan Bitton, Pablo Blanco-Schweizer, Catherine Blier, Frank Bloos, Mathieu Blot, Lucille Blumberg, Polina Bobkova, Filomena Boccia, Laetitia Bodenes, Debby Bogaert, Anne-Hélène Boivin, Ariel Bolanga, Isabela Bolaños, Pierre-Adrien Bolze, Patrizia Bonelli, Aurelius Bonifasius, Joe Bonney, Diogo Borges, Raphaël Borie, Hans Martin Bosse, Elisabeth Botelho-Nevers, Lila Bouadma, Olivier Bouchaud, Sabelline Bouchez, Damien Bouhour, Kévin Bouiller, Laurence Bouillet, Camile Bouisse, Latsaniphone Bountthasavong, Anne-Sophie Boureau, John Bourke, Maude Bouscambert, Aurore Bousquet, Marielle Boyer-Besseyre, Maria Boylan, Fernando Augusto Bozza, Axelle Braconnier, Cynthia Braga, Timo Brandenburger, Luca Brazzi, Patrick Breen, Dorothy Breen, David Brewster, Kathy Brickell, Christopher Brighting, Tessa Broadley, Helen Brotherton, Alex Browne, Nicolas Brozzi, Sonja Hjellegjerde Brunvoll, Marjolein Brusse-Keizer, Petra Bryda, Filipa Brás Monteiro, Nina Buchtele, Polina Bugaeva, Marielle Buisson, Danilo Buonsenso, Erlina Burhan, Donald Buri, Aidan Burrell, Ingrid G. Bustos, Denis Butnaru, Roar Bævre-Jensen, André Cabie, Susana Cabral, Joana Cabrita, Eder Caceres, Cyril Cadoz, Rui Caetano Garcês, Mia Callahan, Kate Calligy, Jose Andres Calvache, Caterina Caminiti, Valentine Campana, Paul Campbell, Josie Campisi, João Camões, Cecilia Canepa, Mireia Cantero, Janice Caoili, Pauline Caraux-Paz, Nelson Cardoso, Filipe Cardoso, Sofia Cardoso, Filipa Cardoso, Simone Carelli, Francesca Carlacci, Nicolas Carlier, Thierry Carmoi, Gayle Carney, Inês Carqueja, Marie-Christine Carret, François Martin Carrier, Ida Carroll, Gail Carson, Leonor Carvalho, Maire-Laure Casanova, Mariana Cascão, Siobhan Casey, José Casimiro, Bailey Cassandra, Nidyanara Castanheira, Silvia Castañeda, Guylaine Castor-Alexandre, Ivo Castro, Ana Catarino, François-Xavier Catherine, Roberta Cavalin, Giulio Giovanni Cavalli, Alexandros Cavayas, Adrian Ceccato, Masaneh Ceesay, Shelby Cerkovnik, Minerva Cervantes-Gonzalez, Muge Cevik, Anissa Chair, Catherine Chakveatze, Adrienne Chan, Meera Chand, Jean-Marc Chapplain, Charlotte Charpentier, Julie Chas, Muhammad Mobin Chaudry, Anjellica Chen, Yih-Sharng Chen, Léo Chenard, Matthew Pellan Cheng, Antoine Cheret, Alfredo Antonio Chetta, Thibault Chiarabini, Julian Chica, Suresh Kumar Chidambaram, Leong Chin Tho, Catherine Chirouze, Davide Chiumello, Hwa Jin Cho, Sung-Min Cho, Bernard Cholley, Danoy Chommanam, Marie-Charlotte Chopin, Ting Soo Chow, Yock Ping Chow, Nathaniel Christy, Jonathan Chua, Hiu Jian Chua, Jonathan Samuel Chávez Iñiguez, Jose Pedro Cidade, José Miguel Cisneros Herreros, Barbara Wanjiru Citarella, Anna Ciullo, Jennifer Clarke, Rolando Claure-Del Granado, Sara Clohisey, Cassidy Codan, Caitriona Cody, Jennifer Coles, Megan Coles, Gwenhaël Colin, Michael Collins, Pamela Combs, Jennifer Connolly, Marie Connor, Anne Conrad, Sofía Contreras, Elaine Conway, Graham S. Cooke, Hugues Cordel, Amanda Corley, Sabine Cornelis, Alexander Daniel Cornet, Arianne Joy Corpuz, Andrea Cortegiani, Grégory Corvaisier, Camille Couffignal, Sandrine Couffin-Cadiergues, Roxane Courtois, Stéphanie Cousse, Juthaporn Cowan, Rachel Cregan, Charles Crepy D’Orleans, Cosimo Cristella, Gloria Crowl, Jonathan Crump, Claudina Cruz, Juan Luis Cruz Bermúdez, Jaime Cruz Rojo, Marc Csete, Alberto Cucino, Ailbhe Cullen, Matthew Cummings, Gerard Curley, Elodie Curlier, Colleen Curran, Paula Custodio, Sheila Cárcel, Umberto D’Alessandro, Federico D’Amico, Frédérick D’Aragon, Eric D’Ortenzio, Charlene Da Silveira, Al-Awwab Dabaliz, Andrew Dagens, John Arne Dahl, Darren Dahly, Peter Daley, Zaina Dalloul, Jo Dalton, Heidi Dalton, Seamus Daly, Juliana Damas, Joycelyn Dame, Cammandji Damien, Hegazy Dana, Nick Daneman, Jorge Dantas, Etienne De Montmollin, Rosanna De Rosa, Cristina De Rose, Bianca DeBenedictis, Nathalie DeCastro, Jillian Deacon, David Dean, Alexa Debard, Marie-Pierre Debray, William Dechert, Romain Decours, Eve Defous, Isabelle Delacroix, Alexandre Delamou, Eric Delaveuve, Karen Delavigne, Nathalie M. Delfos, Ionna Deligiannis, Andrea Dell’Amore, Christelle Delmas, Pierre Delobel, Corine Delsing, Elisa Demonchy, Emmanuelle Denis, Dominique Deplanque, Pieter Depuydt, Mehul Desai, Diane Descamps, Mathilde Desvallées, Santi Dewayanti, Pathik Dhangar, Souleymane Taran Diallo, Alpha Diallo, Sylvain Diamantis, André Dias, Andrea Dias, Fernanda Dias Da Silva, Juan Jose Diaz, Rodrigo Diaz, Priscila Diaz, Bakary K Dibba, Kévin Didier, Jean-Luc Diehl, Wim Dieperink, Jérôme Dimet, Vincent Dinot, Fara Diop, Alphonsine Diouf, Yael Dishon, Cedric Djadda, Félix Djossou, Annemarie B. Docherty, Helen Doherty, Arjen M Dondorp, Christl A. Donnelly, Sean Donohue, Yoann Donohue, Peter Doran, Céline Dorival, Yash Doshi, Phouvieng Douangdala, James Joshua Douglas, Renee Douma, Nathalie Dournon, Joanne Downey, Mark Downing, Thomas Drake, Aoife Driscoll, Amiel A. Dror, Murray Dryden, Ibrahim Kwaku Duah, Claudio Duarte Fonseca, Vincent Dubee, François Dubos, Audrey Dubot-Pérès, Alexandre Ducancelle, Toni Duculan, Susanne Dudman, Abhijit Duggal, Paul Dunand, Jake Dunning, Mathilde Duplaix, Emanuele Durante-Mangoni, Lucian Durham III, Bertrand Dussol, Juliette Duthoit, Xavier Duval, Anne Margarita Dyrhol-Riise, Hamida ELMagrahi, Sim Choon Ean, Ada Ebo, Marco Echeverria-Villalobos, Giorgio Economopoulos, Michael Edelstein, Siobhan Egan, Linn Margrete Eggesbø, Khadeja Ehzaz, Carla Eira, Mohammed El Sanharawi, Marwan El Sayed, Mohammed Elabid, Mohamed Bashir Elagili, Subbarao Elapavaluru, Mohammad Elbahnasawy, Sohail Elboshra, Brigitte Elharrar, Natalie Elkheir, Jacobien Ellerbroek, Merete Ellingjord-Dale, Rami Elmorsi, Mohammad Muatasm Elmubark, Loubna Elotmani, Lauren Eloundou, Philippine Eloy, Abelrahman Elsayed, Basma Elshaikhy, Tarek Elshazly, Wafa Elsokni, Aml Ahmed Eltayeb, Iqbal Elyazar, Zarief Kamel Emad, Hussein Embarek, Isabelle Enderle, Tomoyuki Endo, Gervais Eneli, Chan Chee Eng, Ilka Engelmann, Vincent Enouf, Olivier Epaulard, Haneen Esaadi, Mariano Esperatti, Hélène Esperou, Marina Esposito-Farese, Catarina Espírito Santo, Rachel Essaka, Lorinda Essuman, João Estevão, Hiba Et-taghy, Manuel Etienne, Rachael Evans, Anna Greti Everding, Mirjam Evers, Isabelle Fabre, Marc Fabre, Ismaila Fadera, Asgad Osman Abdalla Fadlalla, Amna Faheem, Amna Faheem, Mohamed Fahim Elalfy, Arabella Fahy, Hamza Faida, Cameron J. Fairfield, Zul Fakar, Komal Fareed, Pedro Faria, Ahmed Farooq, Hanan Fateena, Mohamed Fathi, Salem Fatima, Arie Zainul Fatoni, Karine Faure, Raphaël Favory, Mohamed Fayed, Niamh Feely, Jorge Fernandes, Susana Fernandes, Marília Andreia Fernandes, François-Xavier Ferrand, Eglantine Ferrand Devouge, Carlo Ferrari, Mário Ferraz, Isabel Ferreira, Sílvia Ferreira, Benigno Ferreira, Bernardo Ferreira, Ricard Ferrer-Roca, Nicolas Ferriere, Joana Ferrão, Céline Ficko, Claudia Figueiredo-Mello, William Finlayson, Thomas Flament, Tom Fletcher, Aline-Marie Florence, Letizia Lucia Florio, Brigid Flynn, Deirdre Flynn, Federica Fogliazza, Jean Foley, Victor Fomin, Tatiana Fonseca, Patricia Fontela, Karen Forrest, Simon Forsyth, Denise Foster, Giuseppe Foti, Berline Fotso, Mohamed Fouad Abdrabo, Erwan Fourn, Robert A. Fowler, Robert A. Fowler, Marianne Fraher, Diego Franch-Llasat, Christophe Fraser, John F. Fraser, Marcela Vieira Freire, Ana Freitas Ribeiro, Craig French, Caren Friedrich, Ricardo Fritz, Stéphanie Fry, Nora Fuentes, Masahiro Fukuda, Argin G, Valérie Gaborieau, Rostane Gaci, Massimo Gagliardi, Jean-Charles Gagnard, Amandine Gagneux-Brunon, Nathalie Gagné, Abdou Gai, Linda Gail Skeie, Sérgio Gaião, Adham Mohamed Galal Mohamed Ramadan, Phil Gallagher, Elena Gallego Curto, Carrol Gamble, Aysylu Gamirova, Yasmin Gani, Arthur Garan, Rebekha Garcia, Julia Garcia-Diaz, Esteban Garcia-Gallo, Noelia García Barrio, Navya Garimella, Federica Garofalo, Denis Garot, Valérie Garrait, Basanta Gauli, Anatoliy Gavrylov, Alexandre Gaymard, Johannes Gebauer, Eva Geraud, Louis Gerbaud Morlaes, Nuno Germano, Ahmed Noman Ghaleb, Abdulrahman Ghaleb, Malak Ghemmeid, Praveen Kumar Ghisulal, Jade Ghosn, Marco Giani, Carlo Giaquinto, Tristan Gigante, Elaine Gilroy, Guillermo Giordano, Michelle Girvan, Valérie Gissot, Jesse Gitaka, Gezy Giwangkancana, Daniel Glikman, Petr Glybochko, Eric Gnall, Geraldine Goco, François Goehringer, Siri Goepel, Jean-Christophe Goffard, Jin Yi Goh, Jonathan Golob, Brigitta Golács, Rui Gomes, Kyle Gomez, Kyle Gomez, Marie Gominet, Alicia Gonzalez, Patricia Gordon, Yanay Gorelik, Isabelle Gorenne, Laure Goubert, Cécile Goujard, Tiphaine Goulenok, Margarite Grable, Jeronimo Graf, Edward Wilson Grandin, Pascal Granier, Giacomo Grasselli, Lorenzo Grazioli, Christopher A. Green, Courtney Greene, William Greenhalf, Segolène Greffe, Domenico Luca Grieco, Matthew Griffee, Matthew Griffee, Fiona Griffiths, Ioana Grigoras, Albert Groenendijk, Fassou Mathias Grovogui, Heidi Gruner, Yusing Gu, Fabio Guarracino, Jérémie Guedj, Martin Guego, Anne-Marie Guerguerian, Daniela Guerreiro, Romain Guery, Anne Guillaumot, Laurent Guilleminault, Thomas Guimard, Maisa Guimarães de Castro, Joan Gómez-Junyent, Marieke Haalboom, Marieke Haalboom, Daniel Haber, Ali Hachemi, Amy Hackmann, Abdurrahman Haddud, Nadir Hadri, Wael Hafez, Fakhir Raza Haidri, Fatima Mhd Rida Hajij, Sheeba Hakak, Matthew Hall, Adam Hall, Sophie Halpin, Shaher Hamdan, Abdelhafeez Hamdi, Jawad Hameed, Ansley Hamer, Raph L. Hamers, Rebecca Hamidfar, Bato Hammarström, Naomi Hammond, Terese Hammond, Lim Yuen Han, Matly Hanan, Rashan Haniffa, Kok Wei Hao, Hayley Hardwick, Samuel Bernard Ekow Harrison, Janet Harrison, Ewen M. Harrison, Alan Hartman, Sulieman Hasan, Mohammad Ali Nabil Hasan, Mohd Shahnaz Hasan, Madiha Hashmi, Junaid Hashmi, Amoni Hassan, Ebtisam Hassanin, Claire Hastie, Muhammad Hayat, Ailbhe Hayes, Leanne Hays, Jan Heerman, Lars Heggelund, Ahmed Helmi, Ross Hendry, Martina Hennessy, Aquiles Rodrigo Henriquez-Trujillo, Maxime Hentzien, Diana Hernandez, Daniel Herr, Andrew Hershey, Liv Hesstvedt, Astarini Hidayah, Rupert Higgins, Eibhlin Higgins, Samuel Hinton, Hiroaki Hiraiwa, Haider Hirkani, Hikombo Hitoto, Yi Bin Ho, Antonia Ho, Alexandre Hoctin, Isabelle Hoffmann, Wei Han Hoh, Oscar Hoiting, Rebecca Holt, Jan Cato Holter, Peter Horby, Juan Pablo Horcajada, Kota Hoshino, Koji Hoshino, Ikram Houas, Mabrouka Houderi, Catherine L. Hough, Stuart Houltham, Jimmy Ming-Yang Hsu, Jean-Sébastien Hulot, Stella Huo, Abby Hurd, Iqbal Hussain, Aliae Mohamed Hussein, Mahmood Hussein, Fatima Ibrahim, Bashir Ibran, Samreen Ijaz, M. Arfan Ikram, Carlos Cañada Illana, Patrick Imbert, Muhammad Imran Ansari, Rana Imran Sikander, Carmen Infante Dominguez, Yun Sii Ing, Hugo Inácio, Elias Iosifidis, Mariachiara Ippolito, Vera Irawany, Sarah Isgett, Tiago Isidoro, Nadiah Ismail, Margaux Isnard, Mette Stausland Istre, Junji Itai, Asami Ito, Daniel Ivulich, Danielle Jaafar, Salma Jaafoura, Hamza Jaber, Julien Jabot, Clare Jackson, Abubacarr Jagne, Victoria Janes, Waasila Jassat, Stéphane Jaureguiberry, Jeffrey Javidfar, Jeffrey Javidfar, Denise Jaworsky, Florence Jego, Anilawati Mat Jelani, Synne Jenum, Edwin Jesudason, Ruth Jimbo-Sotomayor, Ong Yiaw Joe, Ruth Noemí Jorge García, Mark Joseph, Cédric Joseph, Swosti Joshi, Mercé Jourdain, Philippe Jouvet, Anna Jung, Hanna Jung, Dafsah Juzar, Silje Bakken Jørgensen, Ouifiya Kafif, Florentia Kaguelidou, Neerusha Kaisbain, Thavamany Kaleesvran, Sabina Kali, Alina Kalicinska, Karl Trygve Kalleberg, Smaragdi Kalomoiri, Muhammad Aisar Ayadi Kamaluddin, Armand Saloun Kamano, Zul Amali Che Kamaruddin, Nadiah Kamarudin, Kavita Kamineni, Darshana Hewa Kandamby, Kong Yeow Kang, Darakhshan Kanwal, Dyah Kanyawati, Valentina Kapustina, Mohamed Karghul, Pratap Karpayah, Todd Karsies, Christiana Kartsonaki, Daisuke Kasugai, Kevin Katz, Tatsuya Kawasaki, Christy Kay, Lamees Kayyali, Seán Keating, Pulak Kedia, Yvelynne Kelly, Aoife Kelly, Sadie Kelly, Andrea Kelly, Niamh Kelly, Claire Kelly, Maeve Kelsey, Ryan Kennedy, Kalynn Kennon, Sommay Keomany, Maeve Kernan, Younes Kerroumi, Sharma Keshav, Evelyne Kestelyn, Shams Khail, Dina Osman Khair, Dalia Osman Hamed Mohamed Khair, Sarah Khaled, Imrana Khalid, Antoine Khalil, Quratul Ain Khan, Irfan Khan, Sushil Khanal, Abid Khatak, Krish Kherajani, Michelle E. Kho, Saye Khoo, Ryan Khoo, Denisa Khoo, Muhammad Nasir Khoso, Amin Khuwaja, Khor How Kiat, Yuri Kida, Harrison Kihuga, Peter Kiiza, Beathe Kiland Granerud, Anders Benjamin Kildal, Anders Benjamin Kildal, Jae Burm Kim, Antoine Kimmoun, Detlef Kindgen-Milles, Nobuya Kitamura, Stany Kivuruga, Eyrun Floerecke Kjetland Kjetland, Paul Klenerman, Rob Klont, Gry Kloumann Bekken, Stephen R Knight, Robin Kobbe, Paa Kobina Forson, Chamira Kodippily, Malte Kohns Vasconcelos, Sabin Koirala, Mamoru Komatsu, Franklina Korkor Abebrese, Volkan Korten, Stephanie Kouba, Mohamed Lamine Kourouma, Karifa Kourouma, Karifa Kourouma, Arsène Kpangon, Karolina Krawczyk, Ali Kredan, Sudhir Krishnan, Vinothini Krishnan, Oksana Kruglova, Anneli Krund, Pei Xuan Kuan, Ganesh Kumar, Mukesh Kumar, Deepali Kumar, Ashok Kumar, Dinesh Kuriakose, Ethan Kurtzman, Neurinda Permata Kusumastuti, Demetrios Kutsogiannis, Galyna Kutsyna, Ama Kwakyewaa Bedu-Addo, Sylvie Kwedi, Konstantinos Kyriakoulis, Erwan L’Her, Marie Lachatre, Marie Lacoste, John G. Laffey, Nadhem Lafhej, Marie Lagrange, Fabrice Laine, Olivier Lairez, Sanjay Lakhey, Sulaiman Lakoh, Antonio Lalueza, Marc Lambert, François Lamontagne, François Lamontagne, Marie Langelot-Richard, Vincent Langlois, Eka Yudha Lantang, Marina Lanza, Cédric Laouénan, Samira Laribi, Delphine Lariviere, Stéphane Lasry, Sakshi Lath, Naveed Latif, Youssef Latifeh, Odile Launay, Didier Laureillard, Yoan Lavie-Badie, Andy Law, Cassie Lawrence, Teresa Lawrence, Minh Le, Clément Le Bihan, Cyril Le Bris, Georges Le Falher, Lucie Le Fevre, Quentin Le Hingrat, Marion Le Maréchal, Soizic Le Mestre, Gwenaël Le Moal, Vincent Le Moing, Hervé Le Nagard, Sylvie LeGac, Ema Leal, Marta Leal Santos, Yi Lin Lee, Biing Horng Lee, James Lee, Heng Gee Lee, Su Hwan Lee, Todd C. Lee, Jennifer Lee, Gary Leeming, Bénédicte Lefebvre, Laurent Lefebvre, Benjamin Lefèvre, Merili-Helen Lehiste, Jean-Daniel Lelievre, François Lellouche, Adrien Lemaignen, Véronique Lemee, Anthony Lemeur, Gretchen Lemmink, Ha Sha Lene, Jenny Lennon, Marc Leone, Tanel Lepik, Quentin Lepiller, François-Xavier Lescure, Olivier Lesens, Mathieu Lesouhaitier, Amy Lester-Grant, Andrew Letizia, Sophie Letrou, Yves Levy, Bruno Levy, Claire Levy-Marchal, Katarzyna Lewandowska, Rafael León, Gianluigi Li Bassi, Janet Liang, Ali Liaquat, Geoffrey Liegeon, Kah Chuan Lim, Wei Shen Lim, Chantre Lima, Lim Lina, Bruno Lina, Andreas Lind, Maja Katherine Lingad, Guillaume Lingas, Sylvie Lion-Daolio, Samantha Lissauer, Keibun Liu, Marine Livrozet, Patricia Lizotte, Antonio Loforte, Navy Lolong, Leong Chee Loon, Diogo Lopes, Dalia Lopez-Colon, Jose W. Lopez-Revilla, Anthony L. Loschner, Paul Loubet, Bouchra Loufti, Guillame Louis, Silvia Lourenco, Lara Lovelace-Macon, Lee Lee Low, David Lowe, Marije Lowik, Jia Shyi Loy, Jean Christophe Lucet, Carlos Lumbreras Bermejo, Carlos M. Luna, Olguta Lungu, Miles Lunn, Liem Luong, Nestor Luque, Dominique Luton, Olavi Maasikas, Sarah MacDonald, Sarah MacDonald, Moïse Machado, Sara Machado, Gabriel Macheda, Juan Macias Sanchez, Jai Madhok, Guillermo Maestro de la Calle, Jacob Magara, Giuseppe Maglietta, Rachel Maguru, Mustafa Magzoub, Moataz Maher Emara, Mohammed Maher Hadhoud, Rafael Mahieu, Sophie Mahy, Ana Raquel Maia, Lars S. Maier, Oumou Maiga Ascofare, Mylène Maillet, Thomas Maitre, Nimisha Abdul Majeed, Maria Majori, Maximilian Malfertheiner, Nadia Malik, Paddy Mallon, Fernando Maltez, Denis Malvy, Patrizia Mammi, Victoria Manda, Jose M. Mandei, Laurent Mandelbrot, Frank Manetta, Julie Mankikian, Edmund Manning, Aldric Manuel, Ceila Maria Sant’Ana Malaque, Flávio Marino, Samuel Markowicz, Ana Marques, Catherine Marquis, Laura Marsh, Brian Marsh, Megan Marshal, John Marshall, Celina Turchi Martelli, Dori-Ann Martin, Emily Martin, Guillaume Martin-Blondel, Ignacio Martin-Loeches, Alessandra Martinelli, F. Eduardo Martinez, Martin Martinot, João Martins, Nuno Martins, Ana Martins, Caroline Martins Rego, Gennaro Martucci, Olga Martynenko, Alejandro Martín-Quiros, Eva Miranda Marwali, Marsilla Marzukie, Veronika Maráczi, Juan Fernado Masa Jimenez, David Maslove, Phillip Mason, Sabina Mason, Sobia Masood, Fatma Masoud, Moise Massoma, Palmer Masumbe, Mohd Basri Mat Nor, Moshe Matan, Sébastien Matata, Henrique Mateus Fernandes, Meghena Mathew, Christina Mathew, Girish Matjeti, Mathieu Mattei, Laurence Maulin, Jennifer May, Juergen May, Marcel Mayala, Javier Maynar, Mayfong Mayxay, Mohd Zulfakar Mazlan, Thierry Mazzoni, Placide Mbala, Lisa Mc Sweeney, Colin McArthur, Naina McCann, Peter McCanny, Aine McCarthy, Anne McCarthy, Colin McCloskey, Colin McCloskey, Rachael McConnochie, Sherry McDermott, Sherry McDermott, Sarah E. McDonald, Aine McElroy, Samuel McElwee, Natalie McEvoy, Allison McGeer, Kenneth A. McLean, Paul McNally, Bairbre McNicholas, Edel Meaney, Cécile Mear-Passard, Maggie Mechlin, Nastia Medombou, Ahmed Megdi, Omar Mehkri, Omar Mehkri, Ferruccio Mele, Luis Melo, Kashif Ali Memon, João João Mendes, Ogechukwu Menkiti, Kusum Menon, France Mentré, Alexander J. Mentzer, Emmanuelle Mercier, Noémie Mercier, Antoine Merckx, Mayka Mergeay-Fabre, Blake Mergler, Laura Merson, Tiziana Meschi, António Mesquita, Roberta Meta, Osama Metwally, Agnès Meybeck, Dan Meyer, Alison M. Meynert, Vanina Meysonnier, Mehdi Mezidi, Céline Michelanglei, Isabelle Michelet, Efstathia Mihelis, Vladislav Mihnovit, Duha Milad Abdullah, Jennene Miller, Hugo Miranda-Maldonado, Nor Arisah Misnan, Nik Nur Eliza Mohamed, Nouralsabah Mohamed, Tahira Jamal Mohamed, Alaa Mohamed Ads, Ahmed Reda Mohamed Elsayed Abdelhalim, Libya Mohammed, Shrouk Fawze Mohammed Mostafa, Manahil Omer Abdelrahman Mohammedahmed, Omer Abdullah Mohammedelhassan, Asma Moin, Walaa Mokhtar, Elena Molinos, Brenda Molloy, Mary Mone, Agostinho Monteiro, Claudia Montes, Giorgia Montrucchio, Sarah Moore, Shona C. Moore, Lina Morales Cely, Marwa Morgom, Lucia Moro, Diego Rolando Morocho Tutillo, Ben Morton, Catherine Motherway, Ana Motos, Hugo Mouquet, Clara Mouton Perrot, Julien Moyet, Suleiman Haitham Mualla, Caroline Mudara, Caroline Mudara, Mohamed Muftah, Aisha Kalsoom Mufti, Ng Yong Muh, Mo’nes Muhaisen, Dzawani Muhamad, Jackson Muhindo, Daniel Mukadi, Marithé Mukoka, Jimmy Mullaert, Daniel Munblit, Muller Mundenga, Syed Muneeb Ali, Nadeem Munir, Laveena Munshi, Aisling Murphy, Patrick Murray, Marlène Murris, Srinivas Murthy, Srinivas Murthy, Himed Musaab, Alamin Mustafa, Dana Mustafa, Mus’ab Mustafa, Carlotta Mutti, Himasha Muvindi, Dimitra Melia Myrodia, Karl Erik Müller, Fredrik Müller, Farah Nadia Mohd-Hanafiah, Behzad Nadjm, Dave Nagpal, Alex Nagrebetsky, Blanka Nagybányai-Nagy, Herwin Nanda Boudoin, Mangala Narasimhan, Nageswaran Narayanan, Prashant Nasa, Rashid Nasim Khan, Ahmad Nasrallah, Adel Gerges Nassif Metri, Alasdair Nazerali-Maitland, Ebrahim Ndure, Nadège Neant, Holger Neb, Coca Necsoi, Nikita Nekliudov, Matthew Nelder, Erni Nelwan, Raul Neto, Emily Neumann, Bernardo Neves, Wing Yiu Ng, Pauline Yeung Ng, Anthony Nghi, Jane Ngure, Duc Nguyen, Orna Ni Choileain, Niamh Ni Leathlobhair, Nerissa Niba, Alistair D Nichol, Prompak Nitayavardhana, Stephanie Nonas, Nurul Amani Mohd Noordin, Nurul Faten Izzati Norharizam, Anita North, Alessandra Notari, Moneer Noureldean, Mahdad Noursadeghi, Adam Nowinski, Saad Nseir, Leonard Numfor, Nurnaningsih Nurnaningsih, Dwi Utomo Nusantara, Elsa Nyamankolly, Anders Benteson Nygaard, Fionnuala O Brien, Annmarie O Callaghan, Annmarie O’Callaghan, Max O’Donnell, Sophie O’Halloran, Katie O’Hearn, Conar O’Neil, Linda O’Shea, Miriam O’Sullivan, Derbrenn OConnor, Giovanna Occhipinti, Ebenezer Oduro-Mensah, Lawrence Ofori-Boadu, Tawnya Ogston, Takayuki Ogura, Tak-Hyuk Oh, Sally-Ann Ohene, Shinichiro Ohshimo, Agnieszka Oldakowska, Larissa Oliveira, João Oliveira, Joseph Oliver-Commey, Piero L. Olliaro, Cinderella Omar Rageh Elnaggar, Alsarrah Ali Mohammed Omer, Pierre Ondobo, Jee Yan Ong, David S. Y. Ong, Wilna Oosthuyzen, Anne Opavsky, Peter Openshaw, Saijad Orakzai, Claudia Milena Orozco-Chamorro, Andrés Orquera, Jamel Ortoleva, Mohamed Osama Elsayed Soliman, Javier Osatnik, Siti Zubaidah Othman, Eman Othman, Paul Otiku, Nadia Ouamara, Rachida Ouissa, Christian Owoo, Micheal Owusu, Ama Akyampomaa Owusu-Asare, Clark Owyang, Eric Oziol, Maïder Pagadoy, Justine Pages, Amanda Palacios, Massimo Palmarini, Carlo Palmieri, Giovanna Panarello, Prasan Kumar Panda, Hem Paneru, Lai Hui Pang, Mauro Panigada, Nathalie Pansu, Aurélie Papadopoulos, Paolo Parducci, Edwin Fernando Paredes Oña, Rachael Parke, Melissa Parker, Jérémie Pasquier, Bruno Pastene, Fabian Patauner, Drashti Patel, Mohan Dass Pathmanathan, Patricia Patricio, Patricia Patricio, Laura Patrizi, Luís Patrão, Lisa Patterson, Rajyabardhan Pattnaik, Christelle Paul, Mical Paul, Ellen Pauley, Jorge Paulos, William A. Paxton, Jean-François Payen, Sandra L Peake, Kalaiarasu Peariasamy, Miguel Pedrera Jiménez, Giles J. Peek, Florent Peelman, Nathan Peiffer-Smadja, Vincent Peigne, Mare Pejkovska, Jill Pell, Paolo Pelosi, Ithan D. Peltan, Rui Pereira, Daniel Perez, Thomas Perpoint, Antonio Pesenti, Lenina Pessey, Vincent Pestre, Lenka Petrou, Ventzislava Petrov-Sanchez, Michele Petrovic, Frank Olav Pettersen, Gilles Peytavin, Richard Odame Philips, Ooyanong Phonemixay, Soulichanya Phoutthavong, Michael Piagnerelli, Walter Picard, Olivier Picone, Maria de Piero, Djura Piersma, Carlos Pimentel, Raquel Pinto, Catarina Pires, Lionel Piroth, Roberta Pisi, Ayodhia Pitaloka, Chiara Piubelli, Riinu Pius, Simone Piva, Laurent Plantier, Hon Shen Png, Julien Poissy, Ryadh Pokeerbux, Maria Pokorska-Spiewak, Sergio Poli, Georgios Pollakis, Diane Ponscarme, Jolanta Popielska, Diego Bastos Porto, Andra-Maris Post, Douwe F. Postma, Pedro Povoa, Jeff Powis, Sofia Prapa, Viladeth Praphasiri, Sébastien Preau, Christian Prebensen, Jean-Charles Preiser, Anton Prinssen, Mark G. Pritchard, Gamage Dona Dilanthi Priyadarshani, Lucia Proença, Sravya Pudota, Bambang Pujo Semedi, Mathew Pulicken, Matteo Puntoni, Peter Puplampu, Gregory Purcell, Diana Póvoas, Luisa Quesada, Luisa Quesada, Vilmaris Quinones-Cardona, Víctor Quirós González, Else Quist-Paulsen, Mohammed Quraishi, Fadi Qutishat, Maia Rabaa, Christian Rabaud, Ebenezer Rabindrarajan, Aldo Rafael, Marie Rafiq, Abdelrahman Ragab, Gabrielle Ragazzo, Mutia Rahardjani, Arslan Rahat Ullah, Ahmad Kashfi Haji Ab Rahman, Rozanah Abd Rahman, Fernando Rainieri, Giri Shan Rajahram, Pratheema Ramachandran, Nagarajan Ramakrishnan, José Ramalho, Kollengode Ramanathan, Ahmad Afiq Ramli, Blandine Rammaert, Grazielle Viana Ramos, Anais Rampello, Muhammad Asim Rana, Asim Rana, Rajavardhan Rangappa, Ritika Ranjan, Elena Ranza, Christophe Rapp, Aasiyah Rashan, Thalha Rashan, Ghulam Rasheed, Menaldi Rasmin, Cornelius Rau, Francesco Rausa, Tharmini Ravi, Ali Raza, Andre Real, Stanislas Rebaudet, Sarah Redl, Brenda Reeve, Atta Ur Rehman, Attaur Rehman, Muhammad Osama Rehman Khalid, Dag Henrik Reikvam, Renato Reis, Jordi Rello, Jonathan Remppis, Martine Remy, Hongru Ren, Hanna Renk, Anne-Sophie Resseguier, Matthieu Revest, Oleksa Rewa, Luis Felipe Reyes, Luis Felipe Reyes, Maria Ines Ribeiro, Antonia Ricchiuto, Denise Richardson, David Richardson, Laurent Richier, Siti Nurul Atikah Ahmad Ridzuan, Jordi Riera, Ana L Rios, Asgar Rishu, Patrick Rispal, Karine Risso, Maria Angelica Rivera Nuñez, Doug Robb, Chiara Robba, André Roberto, Stephanie Roberts, Charles Roberts, David L. Robertson, Olivier Robineau, Anna Roca, Ferran Roche-Campo, Paola Rodari, Simão Rodeia, Julia Rodriguez Abreu, Bernhard Roessler, Claire Roger, Pierre-Marie Roger, Emmanuel Roilides, Amanda Rojek, Roberto Roncon-Albuquerque Jr, Mélanie Roriz, Manuel Rosa-Calatrava, Michael Rose, Dorothea Rosenberger, Andrea Rossanese, Matteo Rossetti, Sandra Rossi, Patrick Rossignol, Carine Roy, Benoît Roze, Desy Rusmawatiningtyas, Clark D. Russell, Maeve Ryan, Steffi Ryckaert, Aleksander Rygh Holten, Indrek Rätsep, Indrek Rätsep, Isabela Saba, Luca Sacchelli, Sairah Sadaf, Musharaf Sadat, Valla Sahraei, Abdurraouf Said, Moumen Said Ellawi, Nadia Saidani, Pranya Sakiyalak, Fodé Bangaly Sako, Moamen Salah, Ali Alaa Salah Eldin Mohamed Abbas, Nawal Salahuddin, Leonardo Salazar, Jodat Saleem, Mohammed Saleh Alyasiri, Talat Ahmed Abu Salem, Gabriele Sales, Charlotte Salmon Gandonniere, Hélène Salvator, Shaden Samardali, Dana Samardali, Yehia Samir Shaaban Aly Orabi, Olivier Sanchez, Emely Sanchez, Kizy Sanchez de Oliveira, Angel Sanchez-Miralles, Vanessa Sancho-Shimizu, Gyan Sandhu, Zulfiqar Sandhu, Pierre-François Sandrine, Marlene Santos, Shirley Sarfo-Mensah, Bruno Sarmento Banheiro, Iam Claire E. Sarmiento, Benjamine Sarton, Ankana Satya, Sree Satyapriya, Rumaisah Satyawati, Egle Saviciute, Parthena Savvidou, Yen Tsen Saw, Islam Sayed, Justin Schaffer, Tjard Schermer, Arnaud Scherpereel, Marion Schneider, János Schnur, Stephan Schroll, Michael Schwameis, Gary Schwartz, Janet T. Scott, James Scott-Brown, Nicholas Sedillot, Tamara Seitz, Jaganathan Selvanayagam, Mageswari Selvarajoo, Malcolm G. Semple, Malcolm G. Semple, Rasidah Bt Senian, Eric Senneville, Claudia Sepulveda, Filipa Sequeira, Tânia Sequeira, Ary Serpa Neto, Pablo Serrano Balazote, Ellen Shadowitz, Syamin Asyraf Shahidan, Hamza Shahla, Laila Shalabi, Haitam Shames, Anuraj Shankar, Shaikh Sharjeel, Pratima Sharma, Catherine A. Shaw, Victoria Shaw, Wejdan Ahmed Shawlan, Ahmed Shazly, John Robert Sheenan, Dr. Rajesh Mohan Shetty, Rohan Shetty, Nisreen Shiban, Mohiuddin Shiekh, Takuya Shiga, Anastasia Shikhaleva, Nobuaki Shime, Naoki Shimizu, Keiki Shimizu, Hiroaki Shimizu, Sally Shrapnel, Pramesh Sundar Shrestha, Shubha Kalyan Shrestha, Hoi Ping Shum, Nassima Si Mohammed, Ng Yong Siang, Moses Siaw-Frimpong, Jeanne Sibiude, Bountoy Sibounheuang, Nidhal Siddig, Maqsood Ahmed Siddiqui, Atif Siddiqui, Louise Sigfrid, Louise Sigfrid, Fatoumata Sillah, Piret Sillaots, Rogério Silva, Maria Joao Silva, Catarina Silva, Benedict Sim Lim Heng, Wai Ching Sin, Dario Sinatti, Girish Sindhwani, Punam Singh, Mahendra Singh, Pompini Agustina Sitompul, Karisha Sivam, Vegard Skogen, Sue Smith, Benjamin Smood, Coilin Smyth, Morgane Snacken, Dominic So, Tze Vee Soh, Lene Bergendal Solberg, Joshua Solomon, Tom Solomon, Emily Somers, Agnès Sommet, Myung Jin Song, Rima Song, Tae Song, Jack Song Chia, Albert Sotto, Edouard Soum, Marta Sousa, Ana Chora Sousa, Maria Sousa Uva, Vicente Souza-Dantas, Mamadou Saliou Sow, Alexandra Sperry, Elisabetta Spinuzza, Ekaterina Spiridonova, B. P. Sanka Ruwan Sri Darshana, Shiranee Sriskandan, Sarah Stabler, Thomas Staudinger, Stephanie-Susanne Stecher, Stephanie-Susanne Stecher, Trude Steinsvik, Ymkje Stienstra, Birgitte Stiksrud, Eva Stolz, Amy Stone, Zachary Stotz, Anca Streinu-Cercel, Anca Streinu-Cercel, Adrian Streinu-Cercel, Geoff Strong, Ami Stuart, David Stuart, Richa Su, Decy Subekti, Jacky Y. Suen, Gabriel Suen, Prasanth Sukumar, Asfia Sultana, Charlotte Summers, Dubravka Supic, Deepashankari Suppiah, Magdalena Surovcová, Atie Suwarti, Andrey Svistunov, Sarah Syahrin, Augustina Sylverken, Konstantinos Syrigos, Jaques Sztajnbok, Konstanty Szuldrzynski, Xavier Sánchez Choez, Arne Søraas, Oana Săndulescu, Shirin Tabrizi, Fabio S. Taccone, Lysa Tagherset, Shahdattul Mawarni Taib, Ewa Talarek, Sara Taleb, Cheikh Talla, Jelmer Talsma, Renaud Tamisier, Maria Lawrensia Tampubolon, Le Van Tan, Kim Keat Tan, Yan Chyi Tan, Clarice Tanaka, Taku Tanaka, Hiroyuki Tanaka, Hayato Taniguchi, Huda Taqdees, Arshad Taqi, Coralie Tardivon, Yousef Tarek Kamal Mostafa, Ali Tarhabat, Pierre Tattevin, M Azhari Taufik, Hassan Tawfik, Tze Yuan Tee, João Teixeira, Sofia Tejada, Marie-Capucine Tellier, Sze Kye Teoh, Vanessa Teotonio, Olivier Terrier, Nicolas Terzi, Hubert Tessier-Grenier, Adrian Tey, Alif Adlan Mohd Thabit, Anand Thakur, Zhang Duan Tham, Suvintheran Thangavelu, Samar Tharwat, Elmi Theron, Vincent Thibault, Simon-Djamel Thiberville, Benoît Thill, Jananee Thirumanickam, Niamh Thompson, Shaun Thompson, David Thomson, Emma C. Thomson, Matt Thorpe, Mathew Thorpe, Surain Raaj Thanga Thurai, Duong Bich Thuy, Ryan S. Thwaites, Andrea Ticinesi, Paul Tierney, Vadim Tieroshyn, Peter S Timashev, Jean-François Timsit, Bharath Kumar Tirupakuzhi Vijayaraghavan, Noémie Tissot, Fiona Toal, Jordan Zhien Yang Toh, Maria Toki, Kristian Tonby, Sia Loong Tonnii, Marta Torre, Margarida Torres, Antoni Torres, Rosario Maria Torres Santos-Olmo, Hernando Torres-Zevallos, Aboubacar Tounkara, Michael Towers, Fodé Amara Traoré, Tony Trapani, Huynh Trung Trieu, Cécile Tromeur, Ioannis Trontzas, Tiffany Trouillon, Jeanne Truong, Christelle Tual, Sarah Tubiana, Helen Tuite, Alexis F. Turgeon, Jean-Marie Turmel, Lance C.W. Turtle, Anders Tveita, Pawel Twardowski, François Téoulé, Makoto Uchiyama, PG Ishara Udayanga, Andrew Udy, Roman Ullrich, Alberto Uribe, Asad Usman, Effua Usuf, Timothy M. Uyeki, Michel Vaillant, Patemo Vainitoba, Cristinava Vajdovics, Luís Val-Flores, Piero Valentini, Ana Luiza Valle, Ilaria Valzano, Nicky Van Der Vekens, Sylvie Van Der Werf, Jarne Van Hattem, Gitte Van Twillert, Hugo Van Willigen, Stijn Van de Velde, Peter Van der Voort, Noémie Vanel, Henk Vanoverschelde, Michael Varrone, Shoban Raj Vasudayan, Charline Vauchy, Pavan Kumar Vecham, Shaminee Veeran, Aurélie Veislinger, Sebastian Vencken, Sara Ventura, Annelies Verbon, Hervé Viala, José Ernesto Vidal, César Vieira, Deepak Vijayan, Judit Villar, Pierre-Marc Villeneuve, Andrea Villoldo, Nguyen Van Vinh Chau, Gayatri Vishwanathan, Benoit Visseaux, Hannah Visser, Chiara Vitiello, Manivanh Vongsouvath, Harald Vonkeman, Fanny Vuotto, Noor Hidayu Wahab, Suhaila Abdul Wahab, Nadirah Abdul Wahid, Louise Wain, Marina Wainstein, Laura Walsh, Wan Fadzlina Wan Muhd Shukeri, Chih-Hsien Wang, Steve Webb, Jia Wei, Katharina Weil, Tan Pei Wen, Hassi Wesam, Sanne Wesselius, T. Eoin West, Murray Wham, Bryan Whelan, Nicole White, Paul Henri Wicky, Aurélie Wiedemann, Surya Otto Wijaya, Keith Wille, Sue Willems, Sue Willems, Bailey Williams, Patricia J Williams, Virginie Williams, Evert-Jan Wils, Evert-Jan Wils, Jessica Wittman, Yew Sing Wong, Calvin Wong, Xin Ci Wong, Teck Fung Wong, Natalie Wright, Lim Saio Xian, Ioannis Xynogalas, Sophie Yacoub, Siti Rohani Binti Mohd Yakop, Masaki Yamazaki, Elizabeth Yarad, Yazdan Yazdanpanah, Nicholas Yee Liang Hing, Abdelrahman Yehia Mahmoud Abdelaal, Cécile Yelnik, Chian Hui Yeoh, Stephanie Yerkovich, Touxiong Yiaye, Toshiki Yokoyama, Hodane Yonis, Obada Yousif, Saptadi Yuliarto, Akram Zaaqoq, Marion Zabbe, Gustavo E. Zabert, Kai Zacharowski, Masliza Zahid, Maram Zahran, Nor Zaila Binti Zaidan, Maria Zambon, Miguel Zambrano, Mostafa Zanaty, Alberto Zanella, Konrad Zawadka, Nurul Zaynah, Hiba Zayyad, Alexander Zoufaly, David Zucman, Ana da Silva Filipe, Mark de Boer, Menno de Jong, Gillian de Loughry, Diego de Mendoza, Rafael Freitas de Oliveira França, Ana Isabel de Pinho Oliveira, Thushan de Silva, Peter de Vries, Marlice van Dyk, Laura van Gulik, Carolien van Netten, Frank van Someren Gréve, Ilonka van Veen, Marcel van den Berge, Machteld van der Feltz, Job van der Palen, Paul van der Valk, Mazankowski Heart Institute, The Western Australian COVID-19 Research Response, PHOSP Collaborative Group, Long COVID India - Terna Specialty Hospital and Research Centre.

### Contributing Sites

#### Argentina

Clinica Pasteur National - University of Comahue, Neuquén; Hospital Alemán, Buenos Aires; Hospital De Niños “Sor María Ludovica”, La Plata; Hospital Municipal de Agudos Leonidas Lucero, Bah; Hospital Nacional Profesor A. Posadas, Buenos Aires; Hospital de Clínicas, Buenos Aires; Hospital de Pediatría Garrahan, Buenos Aires; Mar del Plata Medical Foundation Private Community Hospital, Mar Del Plata.

#### Australia

Albury Wodonga Health, Albury; Alice Springs Hospital, Alice Springs; Angliss Hospital, Melbourne; Austin Hospital, Melbourne; Ballarat Base Hospital, Ballarat; Bankstown-Lidcombe Hospital, Bankstown; Barwon Health, Geelong; Bendigo Hospital, Bendigo; Box Hill Hospital, Melbourne; Bunbury Hospital, Bunbury; Bundaberg Hospital, Bundaberg; Caboolture Hospital, Caboolture; Cabrini Hospital, Melbourne; Cairns Hospital, Cairns; Calvary Mater Hospital, Newcastle; Campbelltown Hospital, Campbelltown; Canberra Hospital, Canberra; Casey Hospital, Melbourne; Concord Hospital, Sydney; Dandenong Hospital, Melbourne; Epworth Hospital, Melbourne; Flinders Medical Centre, Adelaide; Footscray Hospital, Melbourne; Frankston Hospital, Melbourne; Gold Coast University Hospital, Gold Coast; Hervey Bay Hospital, Hervey Bay; Ipswich Hospital, Ipswich; John Hunter Hospital, New Lambton Heights; Joondalup Health Campus, Perth; Launceston General Hospital, Launceston; Lismore Base Hospital, Lismore; Liverpool Hospital, Liverpool; Logan Hospital, Logan; Lyell McEwan Hospital, Adelaide; Maroondah Hospital, Melbourne; Mater Misericordiae Brisbane, Brisbane; Mildura Base Hospital, Mildura; Monash Children’s Hospital, Melbourne; Monash Medical Centre, Melbourne; Monash University, Melbourne; Nepean Hospital, Sydney; Northeast Health Wangaratta, Wangaratta; Northern Health, Melbourne; Perth Children’s Hospital, Perth; Prince of Wales Hospital, Sydney; Princess Alexandra Hospital, Brisbane; Queensland Children’s Hospital, Brisbane; Redcliffe Hospital, Redcliffe; Rockingham Hospital, Rockingham; Royal Adelaide Hospital, Adelaide; Royal Brisbane and Women’s Hospital, Brisbane; Royal Children’s Hospital, Melbourne; Royal Darwin Hospital, Tiwi; Royal Hobart Hospital, Hobart; Royal Melbourne Hospital, Melbourne; Royal North Shore Hospital, Sydney; Royal Perth Hospital, Perth; Royal Prince Alfred Hospital, Sydney; Sir Charles Gairdner Hospital, Perth; St George Hospital, Sydney; St John of God Hospital, Perth; St John of God Hospital, Midland, Perth; St Vincent’s Hospital, Melbourne; St. Vincent, Sydney; Sunshine Coast University Hospital, Sunshine Coast; Sunshine Hospital, Melbourne; Sydney Children’s Hospital, Sydney; The Alfred Hospital, Melbourne; The Prince Charles Hospital, Brisbane; The Queen Elizabeth Hospital, Adelaide; Toowoomba Hospital, Toowoomba; University of Western Australia/Fiona Stanley Hospital, Murdoch; Warrnambool, Warrnambool; Werribee Mercy Hospital, Werribee; Westmead Hospital, Sydney; Wollongong Hospital, Wollongong; Women’s and Children’s Hospital, Adelaide.

#### Austria

Medical University of Vienna, Vienna; Sozialmedizinisches Zentrum Sud, Vienna. Bangladesh

NICVD Dhaka, Dhaka. Belgium AZ Maria Middelares, Gent; CUB-Hopital Erasme, Anderlecht; Civil Hospital Marie Curie, Charleroi; PREPARE and RECOVER EU Consortium; St-Pierre University Hospital, Brussels; Universitair Ziekenhuis, Gent.

#### Bolivia

Caja Nacional De Salud, Trinidad; Hospital del Niño Dr. Ovidio Aliaga Uria, La Paz; Hospital del Niño Manuel A. Villarroel, Cochambamba.

#### Botswana

Lenmed Bokamoso Private Hospital, Gaborone. Brazil Centro de Pesquisa Aggeu Magalhães, Fiocruz, Recife; Complexo Hospitalar Dr. Clementino Fraga, João Pessoa city; Hospital Escola da Universidade Federal de Pelotas, Pelotas; Hospital Naval Marcílio Dias, Rio De Janeiro; Hospital Sirio-Libanes, Sao Paulo; Hospital Universitário Clementino Fraga Filho, Rio de Janeiro; Hospital das Clinicas da Faculdade de Medicina da Universidade de Sao Paulo, Sao Paulo; Hospital de Amor, Sao Paulo; Instituto de Infectologia Emílio Ribas, Sao Paulo; Instituto do Coração da Universidade de São Paulo (INCOR), São Paulo; Mater Dei Hospital, Belo Horizonte; National Institute of Infectious Disease Evandro Chagas, Oswaldo Cruz Foundation (INI-FIOCRUZ), Ministry of Health, and D’Or Institute of Research and Education (IDOR), Rio de Janeiro; São Camilo Cura D’ars, Fortaleza.

#### Cameroon

Ad Lucem Bonamoussadi, Douala; Cass Nkolndongo, Yaoundé; Centre Hospitalier d’Essos, Yaoundé; Centre Médical Concha, Yaoundé; Clinical De Bonamoussadi, Douala; Clinique La Source, Douala; Croix-Rouge Camerounaise Hopital Médical Henry Dunant, Yaoundé; Etoug-Ebe Baptist Hospital, Yaoundé; Hospital Ad Lucem Bonaberi, Bonabéri; Laboratoire Du Lac, Nkolnsam; Obala District Hospital, Obala; St Elizabeth General Hospital, Mutengene; St. Therese Clinic, Yaoundé; University of Yaounde, Yaounde.

#### Canada

Alberta Children’s Hospital, Calgary; BC Children’s Hospital, Vancouver; Brantford General Hospital, Brantford; CISSS Chaudière-Appalaches, Quebec; Centre hospitalier Universitaire de Sherbrooke, Sherbrooke; Centre hospitalier de l’université de Montréal, Montreal; Children’s Hospital of Eastern Ontario, Ottawa; Foothills Medical Centre, Calgary; Grand River Hospital, Kitchener; Grande Prairie Queen Elizabeth II, Grande Prairie; Grey Nun’s Community Hospital, Edmonton; Hospital du Sacre Coeur, Montreal; Humber River Hospital, Toronto; Hôpital de l’Enfant-Jésus, Quebec; Institut Universitaire de Cardiologie et de Pneumologie de Québec, Quebec City; Joseph Brant Hospital, Burlington; Kingston Health Sciences Centre, Kingston; Lions Gate Hospital, Vancouver; London Health Sciences Centre, London; Mazankowski Heart Institute, Edmonton; McGill University Health Centre, Montreal; McMaster University, Hamilton; McMaster University REVIVe Group, Hamilton; Memorial University, St. John’s, Newfoundland, St John’s; Michael Garron Hospital, Toronto; Mills Memorial Hospital, Terrace; Misericordia Community Hospital, Edmonton; Mount Sinai Hospital, Toronto; Niagara Health, Niagara; North York General Hospital, Toronto; Red Deer Regional Hospital, Red Deer; Royal Alexandra Hospital, Edmonton; Royal Columbian Hospital, Vancouver; Sinai Health Systems, Toronto; St Joseph’s Health Center, Sherbrooke; St. Boniface Hospital, Manitoba; St. Joseph’s Healthcare Hamilton, Hamilton; Sturgeon Community Hospital, St Albert; Sunnybrook Health Sciences Centre, Toronto; The Centre hospitalier universitaire Sainte-Justine, Montreal; The Hospital for Sick Children (SickKids), Toronto; The Montreal Children’s Hospital, Montreal; The Ottawa Hospital, Ottawa; Unity Health Toronto, Toronto; University Health Network, Toronto; University Hospital Northern British Columbia, Prince George; University Institute of Cardiology and Respirology, Quebec; University of Alberta Adult ICU, Edmonton; University of Manitoba, Manitoba; Vancouver General Hospital, Vancouver; Vancouver Island Health, Vancouver; William Osler Health Sciences System - Etobicoke General Hospital, Toronto. Chile Clinica Alemana De Santiago, Santiago; Clinica Alemana DeSantiago, Santiago; Clinica Las Condes, Santiago; Hospital Carlos Van Buren, Valparaiso; Hospital Clinica Las Condes, Santiago; Hospital Dr. Exequiel Gonz, Santiago; Hospital Félix Bulnes Cerda, Santiago; Hospital Padre Hurtado, Santiago; Hospital Sotero del Rio, Santiago; Instituto Nacional Del Tórax, Santiago.

#### China

Pamela Youde Nethersole Eastern Hospital, Chai Wan; Princess Margaret Hospital, Kwai Hung; Queen Elizabeth Hospital, Yau Ma Tei; Queen Mary Hospital, Pok Fu Lam.

#### Colombia

Clinica Universidad de La Sabana, Chia; Clinica Valle de Lilli, Valle del Cauca; Clínica Infantil Colsubsidio, Bogota; Clínica Infantil Santa María del Lago, Bogota; Clínica de Fracturas y Ortopedia Ltda, Neiva; Fundacion Cardio Infantil Instituto de Cardiologia, Bogota; Fundación Cardiovascular de Colombia, Floridablanca; Fundación Valle del Lili Hospital, Cali; Hospital Infantil Los Ángeles, Pasto; Hospital Pablo Tobón Uribe, Medellin; Hospital Universitario San Ignacio, Bogota; Hospital Universitario San Jorge, Pereira; IMAT Oncomedica, Monter; Instituto Roosevelt, Bogota; UMHES Santa Clara de la Subred Centro Oriente, Bogota; Universidad de la Sabana, Bogota; Universidad del Cauca, Cauca.

#### Czechia

University Hospital Ostrava, Ostrava-Poruba. Democratic Republic of the Congo Heal Africa, Goma; Hôpital provincial, Bukavu; Institut National de Recherche Biomédicale, Kinshasa. Dominican Republic The Center for Diagnosis, Santo Domingo. Ecuador Catholic University, Quito; Hospital General San Francisco, Quito; Hospital de Especialidades Eugenio Espejo, Quito; Universidad de Las Américas, Quito.

#### Egypt

Al Menshawy General Hospital, Tanta; Alexandria University Hospital, Alexandria; Assiut University Hospital, Asyut; Aswan University Hospital, Aswan; Kafrelsheikh University Hospital, Kafr el-Sheikh; Mansoura University Hospital, Mansoura; Nasr City Hospital For Health Insurance, Cairo; Tanta University Hospital, Tanta.

#### Estonia

North Estonia Medical Centre, Tallin; Tartu University Hospital, Tartu. Fiji Fiji National University, Suva. France CHU Carémeau, Nimes; Centre Hospitalier Agen-Nérac, Agen; Centre Hospitalier Alpes-Leman, Contamine-sur-Arve; Centre Hospitalier Annecy Genevois, Annecy; Centre Hospitalier Antoine Béclère, Clamart; Centre Hospitalier Bretagne Atlantique, Vannes; Centre Hospitalier Départemental Vendée, La Roche-sur-Yon; Centre Hospitalier Emile Roux, Le Puy-en-Velay; Centre Hospitalier Henri Duffaut, Avignon; Centre Hospitalier Intercommunal Villeneuve-Saint-Georges, Villeneuve-Saint-Georges; Centre Hospitalier Le Mans, Le Mans; Centre Hospitalier Louis Raffalli, Manosque; Centre Hospitalier Mont-de-Marsan, Mont-de-Marsan; Centre Hospitalier Métropole Savoie, Chambéry; Centre Hospitalier Pierre Oudot, Bourgoin-Jallieu; Centre Hospitalier Régional Metz-Thionville, Metz; Centre Hospitalier Régional et Universitaire de Nancy - Hôpitaux de Brabois, Nancy; Centre Hospitalier Régional et Universitaire de Tours, Tours; Centre Hospitalier Techer, Calais; Centre Hospitalier Universitaire Ambroise-Paré, Boulogne-Billancourt; Centre Hospitalier Universitaire Amiens-Picardie, Amiens; Centre Hospitalier Universitaire Gabriel Montpied, Clermont-Ferrand; Centre Hospitalier Universitaire Grenoble-Alpes, Grenoble; Centre Hospitalier Universitaire Grenoble-Alpes_FU, Grenoble; Centre Hospitalier Universitaire Mitterrand Dijon-Bourgogne, Dijon; Centre Hospitalier Universitaire Rennes (Hôpital Pontchaillou), Rennes; Centre Hospitalier Universitaire Rennes (Hôpital Sud), Rennes; Centre Hospitalier Universitaire Rouen (Center Hospitalier Universitaire de Rouen), Rouen; Centre Hospitalier Universitaire Rouen (Hôpital Charles Nicolle), Rouen; Centre Hospitalier Universitaire Toulouse (IUCT), Toulouse; Centre Hospitalier Universitaire Toulouse (Larrey), Toulouse; Centre Hospitalier Universitaire Toulouse (Rangueil), Toulouse; Centre Hospitalier Universitaire d’Angers, Angers; Centre Hospitalier Universitaire de Besançon, Besançon; Centre Hospitalier Universitaire de Brest, Brest; Centre Hospitalier Universitaire de Lille, Lille; Centre Hospitalier Universitaire de Lyon - HCL, Lyon; Centre Hospitalier Universitaire de Montpellier, Montpellier; Centre Hospitalier Universitaire de Nantes (Hôpital Nord Laennec), Saint-Herblain; Centre Hospitalier Universitaire de Nantes (Hôpital femme-enfant-adolescent), Nantes; Centre Hospitalier Universitaire de Nantes (Hôtel-Dieu), Nantes; Centre Hospitalier Universitaire de Nice (Hôpital Archet), Nice; Centre Hospitalier Universitaire de Nice (Hôpital Pasteur), Nice; Centre Hospitalier Universitaire de Nîmes, Nîmes; Centre Hospitalier Universitaire de Poitiers, Poitiers; Centre Hospitalier Universitaire de Reims, Reims; Centre Hospitalier Universitaire de Saint-Étienne, Saint-Étienne; Centre Hospitalier Universitaire de Strasbourg, Strasbourg; Centre Hospitalier de Bourg-en-Bresse, Bourg-en-Bresse; Centre Hospitalier de Béziers, Béziers; Centre Hospitalier de Cahors, Cahors; Centre Hospitalier de Cholet, Cholet; Centre Hospitalier de Colmar, Colmar; Centre Hospitalier de Dax - Côte d’Argent, Dax; Centre Hospitalier de Digne-les-Bains, Digne-les-Bains; Centre Hospitalier de Melun, Melun; Centre Hospitalier de Pau, Pau; Centre Hospitalier de Perpignan, Perpignan; Centre Hospitalier de Périgueux, Périgueux; Centre Hospitalier de Saint-Denis, Saint-Denis; Centre Hospitalier de Saintonge, Saintes; Centre Hospitalier de Soissons, Soissons; Centre Hospitalier de Toulon, Toulon; Centre Hospitalier de Tourcoing, Tourcoing; Centre Hospitalier du Pays d’Aix, Aix-en-Provence; Centre Hospitalier intercommunal de Créteil, Créteil; Clinique de l’Infirmerie Protestante de Lyon, Lyon; Grand Hôpital de l’Est Francilien (Site de Marne-la-Vallée), Jossigny; Grand Hôpital de l’Est Francilien (Site de Meaux), Meaux; Groupe Hospitalier Diaconesses Croix Saint-Simon, Paris; Hôpital Albert Calmette, Lille; Hôpital Américain de Paris, Neuilly-sur-Seine; Hôpital Arnaud de Villeneuve, Montpellier; Hôpital Avicenne, Bobigny; Hôpital Bel-Air, Thionville; Hôpital Bichat Claude-Bernard AP-HP, Paris; Hôpital Cochin AP-HP, Paris; Hôpital Européen Georges-Pompidou AP-HP, Paris; Hôpital Européen Marseille, Marseille; Hôpital Femme Mère Enfant - HCL, Lyon; Hôpital Foch, Suresnes; Hôpital Henri-Mondor, Créteil; Hôpital Jacques Monod, Le Havre; Hôpital Kremlin-Bicêtre, Le Kremlin-Bicêtre; Hôpital Lariboisière AP-HP, Paris; Hôpital Laënnec - site de Quimper, Quimper; Hôpital Louis-Mourier, Colombes; Hôpital Lyon Sud - HCL, Lyon; Hôpital Necker-Enfants Malades AP-HP, Paris; Hôpital Nord, Marseille; Hôpital Pellegrin, Bordeaux; Hôpital Purpan, Toulouse; Hôpital Raymond-Poincaré, Garches; Hôpital Robert-Debré AP-HP, Paris; Hôpital Saint-Antoine AP-HP, Paris; Hôpital Saint-Joseph, Paris; Hôpital Saint-Louis AP-HP, Paris; Hôpital Tenon AP-HP, Paris; Hôpital d’Instruction des Armées Bégin, Saint-Mandé; Hôpital de la Conception, Marseille; Hôpital de la Croix-Rousse - HCL, Lyon; Hôpital de la Pitié Salpêtrière AP-HP, Paris; Hôpital de la Timone, Marseille; Hôpital privé d’Antony, Antony; INSERM, Paris; Thonon-les-Bains, Thonon-les-Bains.

#### French Guiana

Centre Hospitalier Andrée Rosemon, Cayenne. Gambia Clinical Services Department, MRC Unit, Fajara; Medical Research Council Gambia at LSHTM, Serrekunda. Germany Bernhard Nocht Institute for Tropical Medicine (BNITM), Hamburg; Jena University Hospital, Jena; Klinik und Poliklinik für Innere Medizin II, University Hospital Regensburg, Kiel; Klinikum Passau, Passau; Krankenhaus Barmherzige Brüder, Regensburg; LMU Hospital Munich, Medical Department II, Campus Großhadern, Munich; Uniklinik University Hospital, Frankfurt; University Children’s Hospital, University Medical Center Hamburg-Eppendorf, Hamburg; University Hospital Dusseldorf, Dusseldorf; University Hospital of Tubingen, Tubingen.

#### Ghana

A-AI-001-001; A-AI-001-002; A-AI-001-003; A-AI-002-001; Ga East Municipal Hospital, Accra; Ghana Health Service, Accra; Ghana Infectious Disease Center, Accra; Global Health and Infectious Diseases Research Group-Kumasi Center for Collaborative Research into Tropical Medicine-KNUST, Kumasi; Kintampo Health Research Centre, Kintampo; Pentecost Convention Center, Kasoa; St. Francis Xavier Hospital, Asso-Foso; St. Michael’s Hospital, Pramso; University of Ghana Medical School, Accra; WHO Ghana Country, Accra.

#### Gibraltar

St Bernard’s Hospital, Gibraltar. Greece Hippokration Hospital, Thessaloniki; Sotiria General Hospital, Athens. Guadeloupe Centre Hospitalier Universitaire de Guadeloupe, Pointe-à-Pitre; Saint-Martin, Saint-Martin. Guinea CTP COVID Gbéssia, Conakry; CTP COVID Kénien, Conakry; Centre d’Excellence Africain pour la Prévention et le Contrôle des Maladies Transmissibles (CEA-PCMT), Gamal Abdel Nasser university of Conakry, Conakry; Donka Hospital, Conakry; Health Center Amélioré De Maferenya, Maferenya.

#### Honduras

IHSS Hospital Regional del Norte Israel Salinas, San Pedro Sula. Hungary Heim Pál National Pediatric Institute, Budapest. India ABC Hospital, Visakhapatnam; All India Institute of Medical Sciences, Rishikesh; Apollo First Med Hospital, Chennai; Apollo Hospitals Chennai, Chennai; Apollo Main Hospital, Chennai; Apollo Speciality Hospital - OMR, Chennai; Apollo Speciality Vanagaram, Chennai; Ispat General Hospital, Rourkela; Kerala Institute of Medical Sciences, Trivandrum; Long COVID India - Terna Specialty Hospital and Research Centre, Mumbai; Manipal Hospital Whitefield, Bangalore; Mehta Hospital, Chennai; Pushpagiri Medical College Hospital, Kerala.

#### Indonesia

Dr Sardjito Government Hospital (Paediatric), Yogyakarta; Fatmawati Hospital, Jakarta; Hasan Sadikin Hospital (Adult), Bandung; Murni Teguh Memorial Hospital and Bunda Thamrin Hospital, North Sumatera; National Cardiovascular Center Harapan Kita Jakarta Indonesia, Jakarta; Persahabatan Hospital, Jakarta; Pratama Rada Bolo Hospital, Karitas Hospital and Waikabubak Hospital, Sumba; Prof Dr R. D. Kandou Central Hospital (Adult), Manado; Prof Dr R. D. Kandou Central Hospital (Paediatric), Manado; RSPI Prof Dr Sulianti Saroso, Jakarta; RSUD Dr. Soetomo, Surabaya; RSUD Pasar Minggu, South Jakarta; RSUP Fatmawati, South Jakarta; Saiful Anwar Hospital - Adult ICU, Malang; Saiful Anwar Hospital - PICU, Malang; Sanglah General Hospital (Paediatric), Bali; University Airlangga Hospital (Paediatric), Surabaya.

#### Iraq

Alshifa Center Medical City, Baghdad. Ireland Beacon Hospital, Dublin; Beaumont Hospital, Dublin; Bon Secours Hospital, Cork; Children’s Health Ireland, Dublin; Connolly Hospital Blanchardstown, Dublin; Cork University Hospital, Cork; Galway University Hospital, Galway; Mater Misericordiae University, Dublin; Mercy Hospital, Cork; Our lady of Lourdes Drogheda, Drogheda; Sligo University Hospital (Saolta), Sligo; St James’s Hospital, Dublin; St Vincents University Hospital, Dublin; Tallaght University Hospital, Dublin; University Hospital - Limerick, Limerick; University Hospital - Waterford, Waterford; University Hospital Kerry, Kerry; Wexford General Hospital, Wexford.

#### Israel

Bar-Ilan University, Ramat Gan; Rambam Hospital, Haifa; The Baruch Padeh Medical Center Poriya, Tiberias; Ziv Medical Centre, Safed.

#### Italy

Azienda Ospedaliero Universitario Pisana, Pisa; Azienda Provinciale per i Servizi Sanitari della Provincia Autonoma di Trento, Arco; Fondazione IRCCS Cà Granda Ospedale Maggiore Policlinico, Milan; Fondazione Policlinico Universitario Agostino Gemelli IRCCS, Rome; Istituto Mediterraneo per i Trapianti e Terapie ad Alta Specializzazione, Palermo; Monaldi Hospital, Napoli; Ospedale Molinette, Torino; Ospedale Niguarda, Milan; Ospedale Papa Giovanni XXIII - Bergamo, Bergamo; Ospedale Sacro Cuore Don Calabria, Negrar Di Valpolicella; Ospedale San Gerardo, Monza; Ospedale San Paolo, Milan; Policlinico of Padova, Padova; Policlinicodi Orsola Universitàdi Bologna, Bologna; San Martino Hospital, Genoa; University Hospital Policlinico Paolo Giaccone, Palermo; University Hospital of Parma, Parma; University of Brescia, Brescia; University of Padua, Padua; Università Cattolica del Sacro Cuore, Rome.

#### Japan

Chiba University Hospital, Chiba; Fujieda Municipal General Hospital, Fujieda; Fukuoka University, Fukuoka; Hiroshima University, Hiroshima; Hokkaido University Hospital, Hokkaido; Hyogo Prefectural Kakogawa Medical Center, Hyogo; Kimitsu Chuo Hospital, Chiba; Kouritu Tousei Hospital, Seto City; Kyoto Medical Centre, Kyoto; Kyoto Prefectural University of Medicine, Kyoto; Mie University Hospital, Tsu; Nagoya University Hospital, Nagoya; Obihiro-Kosei General Hospital, Obihiro; Rinku General Medical Center, Osaka; Saiseikai Senri Hospital, Tochigi; Saiseikai Utsunomiya Hospital, Tochigi; Shizuoka Children’s Hospital, Shizuoka; St. Marianna University School of Medicine, Kawasaki; Teine Keijinkai Hospital, Sapporo; Tohoku Medical and Pharmaceutical University, Sendai; Tohoku University, Sendai; Tokyo Metropolitan Tama Medical Center, Tokyo; Yokohama City University Medical Center, Yokohama.

#### Jordan

Prince Hamza Hospital, Amman. Kenya Mount Kenya University, Thika. Kuwait Al-Adan Hospital, Hadiya; Al-Amiri & Jaber Al-Ahmed Hospitals, Kuwait City. Laos Attapeu Provincial Hospital, Attapeu; Lao-Oxford-Mahosot Hospital-Wellcome Trust Research Unit, Vientiane; Luang Namtha Provincial Hospital, Luang Namtha; Salavan Provincial Hospital, Salavan; Xieng Khouang Provincial Hospital, Phonsavan.

#### Libya

Al-Hawari Center of Surgical Specialty, Al-Hawari; Al-Thawra Teaching Hospital, Albayda; Aljufrah Clinical Isolation Center, Al Jufrah; Gharyan Central Hospital, Gharyan; Misurata Medical Center, Misurata; Regdalin Hospital, Regdalin; Sabha Medical Center, Sabha; Soug Althulatha Isolation Center, Tripoli; Swani Health Isolation Center, Tripoli; Tripoli Central Hospital, Tripoli; Tripoli University Hospital, Tripoli.

#### Luxembourg

Luxembourg Institute of Health, Strassen. Malawi Malawi-Liverpool-Wellcome Trust Clinical Research Programme, Lilongwe. Malaysia Hospital Universiti Sains Malaysia (Mix medical & surgical ICU), Kota Bharu; Hospital Universiti Sains Malaysia (SICU), Kota Bharu; International Islamic University Malaysia Medical Centre (IIUMMC), Pahang; Kluang Hospital, Johor; Kuala Lumpur Hospital, WPKL; Lahad Datu Hospital, Sabah; Melaka Hospital, Melaka; National Institutes of Health (NIH), Ministry of Health Malaysia, Setia Alam; Permai Hospital, Johor; Pulau Pinang Hospital, Pulau Pinang; Queen Elizabeth Hospital, Sabah; Raja Perempuan Zainab II Hospital, Kelantan; Raja Permaisuri Bainun Hospital, Perak; Sarawak General Hospital, Sarawak; Sultanah Aminah Hospital, Johor; Sultanah Bahiyah Hospital, Kedah; Sultanah Nur Zahirah Hospital, Terengganu; Sungai Buloh Hospital, Selangor; Sunway Medical Centre, Selangor; Tawau Hospital, Sabah; Tengku Ampuan Afzan Hospital, Pahang; Tuanku Fauziah Hospital, Perlis; Tuanku Ja’afar, Negeri Sembilan; University Malaya Medical Centre, Kuala Lumpur.

#### Mayotte

Centre Hospitalier de Mayotte, Mamoudzou. Mexico Hospitales Puerta de Hierro, Jalisco; UMAE CMN IMSS Hospital de Pediatría, Mexico City; University of Guadalajara Health Sciences Center, Guadalajara.

#### Morocco

Centre Hospitalier Universitaire Ibn Sina, Rabat. Nepal B & B Hospital, Lalitpur; Chitwan Medical College (COVID ICU), Chitwan; Chitwan Medical College (MICU), Chitwan; Grande International Hospital, Kathmandu; Hospital for Advanced Medicine and Surgery (HAMS), Kathmandu; Karuna Hospital, Kathmandu; Nepal Mediciti Hospital (COVID ICU), Lalitpur; Nepal Mediciti Hospital (Nepal Mediciti ICU), Lalitpur; Nidan Hospital, Lalitpur; Om Hospital, Kathmandu; Tribhuvan University Teaching Hospital (COVID ICU), Kathmandu; Tribhuvan University Teaching Hospital (GICU), Kathmandu; Tribhuvan University Teaching Hospital (ICU C), Kathmandu.

#### Netherlands

Adrz, Goes; Alrijne Hospital, Leiden; Beatrix ziekenhuis, Gorinchem; Canisius Wilhelmina Ziekenhuis, Nijmenjen; Catharina Ziekenhuis, Eindhoven; Coöperatie Medisch Specialistisch Bedrijf, Amsterdam; Erasmus Medical Centre, Rotterdam; Flevoziekenhuis, Almere; Franciscus Gasthuis & Vlietland, Rotterdam; Gelre Hospitals, Zutphen; Leiden University Medical Center, Leiden; Maastricht University Medical Centre, Maastricht; Meander Medical Centre, Amersfoort; Medisch Spectrum Twente, Zutphen; Noordwest-Ziekenhuisgroep, Den Helder; Reinier de Graaf Gasthuis, Delft; Spaarne Gasthuis, Haarlem; Tergooi Hospital, Hilversum; University Medical Center Groningen, Groningen; Ziekenhuisgroep Twente, Hengelo.

#### New Zealand

Auckland City Hospital (CVICU), Auckland; Auckland City Hospital (DCCM 82), Auckland; Dunedin Public Hospital, Dunedin; Middlemore Hospital (Canties Manukan Health), Otahuhu; Nelson Hospital, Nelson; Waikato Hospital, Hamilton; Waitemata District Health Board, Auckland; Wellington Regional Hospital, Wellington.

#### Norway

Akershus University Hospital, Nordbyhagen; Drammen Hospital, Drammen; Oslo University Hospital, Oslo; Sykehusapoteket Bærum, Gjettum; The Norwegian Corona Cohort, Oslo; University Hospital of North Norway, Tromso.

#### Pakistan

Abbasi Shaheed Hospital (Covid Ward), Karachi; Abbasi Shaheed Hospital (MICU), Karachi; Abbasi Shaheed Hospital (SARI ICU), Karachi; Abbasi Shaheed Hospital (SICU), Karachi; Azeema Sheikh Hospital, Islamabad; Bahria International Hospital (MICU), Islamabad; Bahria International Hospital (SICU), Islamabad; Critical Care Asia Site A-AF-012-002, Karachi; Critical Care Asia Site A-AF-012-003, Karachi; Critical Care Asia Site A-AF-015-002, Karachi; Darul Sehat Hospital (GICU), Karachi; Darul Sehat Hospital (SARI ICU), Karachi; Doctors Hospital (SARI ICU), Lahore; Doctors Hospital (SICU), Lahore; GMMM Teaching Hospital, Sukkar; Hameed Latif Hospital (MICU), Lahore; Hameed Latif Hospital (SARI ICU), Lahore; Hameed Latif Hospital (SICU), Lahore; Jinnah Hospital, Lahore; Jinnah Post-Graduate Medical Center (SICU), Karachi; Lady Reading hospital (Floor 1-isolation ward), Peshawar; Lady Reading hospital (Floor 2-SARI ICU), Peshawar; Lady Reading hospital (Floor 3-SARI ICU-2), Peshawar; Lady Reading hospital (GICU(OLD), Peshawar; Lady Reading hospital (GICU), Peshawar; Lahore General Hospital, Lahore; Lahore General Hospital (Phase 3 SICU), Lahore; Liaquat University Hospital (SICU), Hyderabad; Mayo Hospital Lahore (BICU), Lahore; NICVD (CCU-2), Karachi; NICVD (MICU), Karachi; NICVD (SICU), Karachi; NICVD (Sari ICU), Karachi; National Hospital & Medical Center (SARI ICU), Lahore; National Hospital & Medical Center (SICU), Lahore; North West General Hospital (COVID Isolation Ward 4), Peshawar; North West General Hospital (COVID ward), Peshawar; North West General Hospital (HDU), Peshawar; North West General Hospital (MICU), Peshawar; North West General Hospital (SARI ICU), Peshawar; North West General Hospital (SICU), Peshawar; PIMS (MICU), Islamabad; PIMS (Sari ICU), Islamabad; Pakistan Kidney & Liver Institute (SARI ICU), Lahore; Pakistan Kidney & Liver Institute (SICU), Lahore; Patel Hospital (GICU), Karachi; Patel Hospital (SARI ICU), Karachi; SIUT Hospital, Karachi; Sheikh Zayed Medical College Rahim yar Khan (Mixed ICU), Rahim yar Khan; Sheikh Zayed Medical College Rahim yar Khan (SARI ICU), Rahim yar Khan; South City Hospital Karachi (MICU), Karachi; South City Hospital Karachi (SARI ICU), Karachi; South City Hospital Karachi (SARI Isolation Ward), Karachi; Ziauddin Medical University Clifton Campus (COVID Isolation Ward), Karachi; Ziauddin Medical University Clifton Campus (MICU), Karachi; Ziauddin Medical University Clifton Campus (SICU), Karachi; Ziauddin Medical University Clifton Campus (Sari ICU), Karachi; Ziauddin Medical University North Nazimabad Campus (3rd Floor COVID Isolation Ward), Karachi; Ziauddin Medical University North Nazimabad Campus (COVID Isolation Ward), Karachi; Ziauddin Medical University North Nazimabad Campus (General COVID Isolation Ward), Karachi; Ziauddin Medical University North Nazimabad Campus (MICU), Karachi; Ziauddin Medical University North Nazimabad Campus (Semi Medicine COVID Isolation Ward), Karachi.

#### Palestine

Al Ahli Hospital, Hebron; Alshifa Hospital, Gaza. Peru Clínica Internacional, Lima; Hospital Emergencia Ate Vitarte, Lima; Hospital Nacional Edgardo Rebagliati Martins, Lima; Hospital de Emergencias Villa El Salvador, Lima; Instituto Nacional del Niño San Borja, Lima.

#### The Philippines

Angeles University Foundation Medical Center, Angeles; Makati Medical Centre, Makati. Poland Consortium IMGEN, Piaseczno; Department of Children’s Infectious Diseases, Medical University of Warsaw, Warsaw; Institute of TB and Lung Diseases, Warsaw; University Hospital in Krakow, Krakow.

#### Portugal

Centro Hospital e Universitário de Coimbra, Coimbra; Centro Hospitalar Universitário do Algarve, Portimão; Centro Hospitalar Universitário do Porto (CHUP), Porto; Centro Hospitalar Vila Nova de Gaia/Espinho, Espinho; Centro Hospitalar de Leiria, Leiria; Centro Hospitalar de Tondela-Viseu, Viseu; Centro Hospitalar e Universitário de Coimbra - Hospital Pediátrico, Coimbra; Comissão de Ética - Unidade Local de Saúde de Matosinhos, Porto; Hospital Beatriz Ângelo, Loures; Hospital Curry Cabral - Intensive Care Unit - UCIP7, Lisbon; Hospital Da Luz, Lisbon; Hospital Egas Moniz, Lisboa; Hospital Espírito Santo de Évora, Évora; Hospital Garcia de Orta, Almada; Hospital Professor Doutor Fernando Fonseca, Amadora; Hospital Santa Maria, Centro Hospitalar Universitário Lisboa Norte, Amadora; Hospital São Francisco Xavier, Lisbon; Hospital Vila Franca de Xira, Lisbon; Hospital de Abrantes - ICU, Abrantes; Hospital de Curry Cabral - Infectious Diseases, Lisbon; Hospital de Curry Cabral - Internal Medicine, Lisbon; Hospital de São José - U.U.M., Lisbon; São João Hospital Centre, Porto; Unidade Local de Saúde de Alto Minho, Viana Do Castelo. Qatar Hamad General Hospital, Doha. Romania Grigore T Popa University of Medicine and Pharmacy, Bucharest; National Institute for Infectious Diseases Matei Bals, Bucharest.

#### Russia

Sechenov University, Moscow. Réunion Centre Hospitalier Félix-Guyon, Saint-Denis; Centre Hospitalier Universitaire de Saint-Pierre, Saint-Pierre. Saint Martin (French) Centre Hospitalier Universitaire de Martinique, Fort-de-France. Saudi Arabia Dr Mohammad Alfagih Hospital, Riyadh; King Abdulaziz Medical City, Riyadh; King Faisal Hospital Research Center, Riyadh; King Saud Medical City, Riyadh.

#### Senegal

CHU de Fann/Albert Royer, Dakar; Centre Hospitalier de l’Ordre de Malte (CHOM), Dakar; Hospital Center University De Fann, Dakar; Hospital Dalal Jamm, Dakar; Hôpital De Ndamatou, Deni Mali Gueye; Hôpital Principal de Dakar, Dakar; Hôpital de Diamnadio, Dakar; Institut Pasteur de Dakar, Dakar.

#### Singapore

National University Hospital, Singapore. South Africa 2 Military Hospital, Cape Town; 3 Military Hospital, Bloemfontein; A-AJ-001-001; A-AJ-001-002; A-AJ-001-003; AbaQulusi Private Hospital, Vryheid; Aberdeen Hospital, Aberdeen; Abraham Esau Hospital, Calvinia; Addington Hospital, Durban; Adelaide Hospital, Adelaide; Ahmed Al-Kadi Private Hospital, Durban; Alan Blyth Hospital, Ladismith; Albert Nzula District Hospital, Trompsburg; Alexandra Hospital, Cape Town; Aliwal North Hospital, Maletswai; All Saints Hospital, Ngcobo; Amajuba Memorial Hospital, Volksrust; Ampath Emfuleni Mediclinic, Vanderbijlpark; Andries Vosloo Hospital, Somerset East; Arwyp Medical Centre, Kempton Park; Aurora Hospital, Gqeberha; Aurora Rehabilitation Hospital, Gqeberha; Bambisana Hospital, Lusikisiki; Barberton Hospital, Barberton; Bayview Private Hopital (Life), Mossel Bay; Beaufort West Hospital, Beaufort West; Bedford Hospital, Bedford; Beethoven Recovery Centre, Hartbeespoort; Belfast Hospital (HA Grove), Belfast; Benedictine Hospital, Nongoma; Bertha Gxowa Hospital, Germiston; Bethal Hospital, Bethal; Bethesda Hospital, Bethesda; Bheki Mlangeni Hospital, Johannesburg; Bill Pickard Hospital, Prieska; Bisho Hospital, Bisho; Bloemcare Psychiatric Hospital, Bloemfontein; Boitumelo Hospital, Kroonstad; Bongani Nurses’ Dormitory Surge Facility, Edenburg; Bongani Regional Hospital, Welkom; Botlokwa Hospital, Mmatseke; Botshabelo Hospital, Botshabelo; Botshelong Empilweni Hospital, Vosloorus; Botshilu Private Hospital, Soshanguve; Brackengate Intermediate Care, Cape Town; Brewelskloof Hospital, Worcester; Brits Hospital, Brits; Bronkhorstspruit Hospital, Bronkhorstspruit; Brooklyn Chest Hospital, Cape Town; Bryanston Subacute, Sandton; Burgersdorp Hospital, Burgersdorp; Busamed - Paardevlei private hospital, Cape Town; Busamed Bram Fischer International Airport Hospital, Bloemfontein; Busamed Gateway Private Hospital, Umhlanga; Busamed Harrismith Private Hospital, Harrismith; Busamed Hillcrest Private Hospital, Outer West Durban; Busamed Modderfontein Private Hospital Orthopaedic and Oncology Centre, Lethabong; Butterworth Hospital, Butterworth; CTICC COVID Intermediate Care, Cape Town; Cairnhall Hospital, Bloemfontein; Cala Hospital, Cala; Caledon Hospital, Caledon; Canzibe Hospital, Ngqeleni; Cape Gate Mediclinic, Cape Town; Capital hospital, Durban; Care Cure Queenstown, Queenstown; Carewell Sub-Acute Hospital, Still Bay; Carletonville Hospital, Carletonville; Carolina Hospital, Carolina; Cathcart Hospital, Cathcart; Catherine Booth Hospital, Amatikulu; Cecilia Makiwane Hospital, Mdantsane; Ceres Hospital, Ceres; Ceza Hospital, Ceza; Charles Johnson Memorial District Hospital, Nqutu; Charlotte Maxeke Hospital, Johannesburg; Chris Hani Baragwanath Hosp, Johannesburg; Christ The King Hospital, Ixopo; Citrusdal Hospital, Citrusdal; Clairwood Hospital, Durban; Clanwilliam Hospital, Clanwilliam; Clinix Naledi Nkanyezi Hospital, Sebokeng; Clinix Solomon Stix Morewa Hospital, Johannesburg; Cloete Joubert Hospital, Barkly East; Cofimvaba Hospital, Cofimvaba; Corona Sub-Acute Hospital, Bethlehem; Cradock Hospital, Cradock; Crescent Clinic, Cape Town; Cullinan Rehabilitation Hospital, Cullinan; DP Marais Santa Centre, Cape Town; DR C N Phatudi Hospital, Mohlahlareng; Daxina Hospital, Lenasia; Daymed Private Hospital, Pietermaritzburg; De Aar Hospital, De Aar; Diamant Hospital, Jagersfontein; Dihlabeng Hospital, Bethlehem; Dilokong Hospital, Driekop; Donald Fraser Hospital, Thohoyandou; Donald Gordon Medical Centre, Johannesburg; Dora Nginza Hospital, Gqeberha; Dordrecht Hospital, Dordrecht; Doris Goodwin Hospital, Pietermaritzburg; Douglas CHC, Douglas; Dr George Mukhari Hospital, Ga-Rankuwa; Dr Harry Surtie Hospital, Upington; Dr JS Moroka Hospital, Thaba Nchu; Dr Malizo Mpehle Hospital, Tsolo; Dr S K Matseke Memorial Hospital, Diepmeadow; Dr Yusaf Dadoo Hospital, Krugersdorp; Duff Scott Memorial Hospital, City of Matlosana; Dundee Hospital, Dundee; Durdoc Hospital, Durban; East Griqualand & Usher Memorial Hospital, Kokstad; Eden Gardens Private Hospital, Pietermaritzburg; Edendale Hospital, Pietermaritzburg; Edenvale Hospital, Edenvale; Eerste River Hospital, Cape Town; Ekombe Hospital, Nkandla; Elim Hospital, Elim; Elizabeth Donkin Hospital, Gqeberha; Elizabeth Ross Hospital, Phuthaditjhaba; Elliot Hospital, Elliot; Ellisras Hospital, Lephalale; Elsie Ballot Hospital, Amersfoort; Emalahleni Private Hospital, Emalahleni; Embhuleni Hospital, Elukwatini; Emmaus Hospital, Winterton; EmoyaMed Private Hospital, Bloemfontein; Empilisweni Hospital, Tienbank; Empilweni Hospital, Gompo; Ermelo Provincial Hospital, Ermelo; Eshowe Hospital, Eshowe; Eskom Academy of Learning Quarantine Site, Midrand; Esselenpark School of Excellence Quarantine Site, Tembisa; Estcourt Hospital, Estcourt; Evander Hospital, Evander; Evuxakeni Hospital, Giyani; FH Odendaal Hospital, Modimolle; False Bay Hospital, Cape Town; Far East Rand Hospital, Springs; Fezi Ngubentombi Provincial Hospital, Sasolburg; Fisha wellness Hospital, Pretoria; Fort Beaufort Hospital, Fort Beaufort; Fort England Hospital, Makhanda; Fort Napier Hospital, Pietermaritzburg; Frere Hospital, East London; Fritz Visser Hospital, Noupoort; Frontier Hospital, Queenstown; GJ Crooke’s Hospital, Scottburgh; Ganyesa Community Hospital, Ganyesa; General Justice Gizenga Mpanza Hospital, Durban; George Hospital, George; George Masebe Hospital, Haakdoring; Glen Grey Hospital-Lady Frere, Emalahleni; Greenville Hospital, Etyeni; Grey Hospital, Qonce; Grey’s Hospital, Pietermaritzburg; Greytown Hospital, Greytown; Greytown Specialized TB Hospital, Greytown; Groblersdal Hospital, Groblersdal; Groote Schuur Hospital, Cape Town; Hanover Park Clinic Wc Par, Cape Town; Harry Comay Hospital Wc Hch, George; Hayani Psychiatric Hospital, Thohoyandou; Heidelberg Hospital, Heidelberg; Heideveld Cdc Wc Hvp, Cape Town; Heideveld Emergency Centre, Cape Town; Helderberg Hospital, Cape Town; Helderberg Village, Somerset West, Cape Town; Helen Joseph Hospital, Johannesburg; Helene Franz Hospital, Bochum; Hermanus Hospital, Hermanus; Hewu Hospital, Lukhanji; Hibiscus Cato Ridge Private Hospital, Cato Ridge; Hibiscus private hospital, Shepstone; Highway Subacute and Rehabilitation Hospital, Hillcrest; Hillandale Health Care Centre, Bloemfontein; Hillcrest Hospital, Hillcrest; Hlabisa Hospital, Nongoma; Holy Cross Hospital, Flagstaff; Hopetown Hospital, Hopetown; House Idahlia Critical Care Surge Facility, Bloemfontein; Humansdorp Hospital, Humansdorp; Indwe Hospital, Indwe; Inkosi Albert Luthuli Hospital, Durban; Isilimela Hospital, Gomolo; Isivivana Private Hospital, Humansdorp; Itemoheng Hospital, Senekal; Itshelejuba Hospital, Highlands; JMH Ascot Park Hospital, Durban; JMH City Hospital, Durban; JMH Isipingo Hospital, Durban; Jamestown Hospital, Jamestown; Jane Furse Hospital, Jane Furse; Job Shimankana Tabane Hospital, Rustenburg; Joe Morolong Memorial Hospital, Vryburg; John Daniel Newsberry Hospital, Clocolan; Jose Pearson TB Hospital, Bethelsdorp; Jubilee Hospital, Temba; Kakamas Hospital, Kakamas; Kalafong Hospital, Pretoria; Kareedouw Hospital, Kareedouw; Karl Bremer Hospital, Cape Town; Kathu private hospital, Kathu; Katleho Hospital, Virginia; Keimoes Hospital, Keimoes; Kgapane Hospital, Modjadjiskloof; Khayelitsha Dist Hospital, Cape Town; Khotsong TB Hospital, Matatiele; Kiaat Private hospital, Mbombela; KimMed Private Hospital, Kimberley; King Dinuzulu Hospital, Berea; King Edward VIII Hospital, Durban; Klerksdorp Hospital, Klerksdorp; Knysna Hospital, Knysna; Knysna Private Hospital Wc Kvh, Knysna; Kokstad Private Hospital, Kokstad; Komani Hospital, Queenstown; Komga Hospital, Komga; Kopanong Hospital, Vereeniging; Kuruman Hospital, Kuruman; Kwa-Magwaza Hospital, KwaMagwaza; KwaDukuza Private Hospital, KwaDukuza; Kwamhlanga Hospital, KwaMhlanga; La Verna Private Hospital, Ladysmith; Lady Grey Hospital, Lady Grey; Ladysmith Hospital, Ladysmith; Laingsburg Hospital, Laingsburg; Lapa Munnik Hospital, Porterville; Lebowakgomo Hospital, Lebowakgomo; Lenmed Ahmed Kathrada Hospital, Johannesburg; Lenmed Ethekwini Hospital and Heart Centre, Newlands East; Lenmed Randfontein Private Hospital, Randfontein; Lenmed Royal Hospital and Heart Centre, Kimberly; Lenmed Zamokuhle Private Hospital, Tembisa; Lentegeur Hospital, Cape Town; Leratong Hospital, Krugersdorp; Letaba Hospital, Nkowankowa; Life Anncron Hospital, Klerksdorp; Life Beacon Bay Private Hospital, East London; Life Bedford Gardens Hospital, Germiston; Life Brackenview, Alberton; Life Brenthurst Hospital, Johannesburg; Life Carstenhof Hospital, Midrand; Life Carstenview Hospital, Midrand; Life Chatsmed Garden Hospital, Chatsworth; Life Cosmos Hospital, Emalahleni; Life Dalview Hospital, Brakpan; Life East London Private Hospital, East London; Life Empangeni Private Hospital, Empangeni; Life Entabeni Hospital, Berea; Life Eugene Marais Hospital, Pretoria; Life Flora Hospital, Roodepoort; Life Fourways Hospital, Sandton; Life Genesis Clinic Saxonwold, Johannesburg; Life Glynnview Hospital, Benoni; Life Groenkloof Hospital, Pretoria; Life Health Care (Faerie Glen), Pretoria; Life Hilton Private Hospital, Hilton; Life Hunterscraig Hospital, Gqeberha; Life Kingsbury Hospital, Cape Town; Life Midmed Hospital, Middelburg; Life Mt Edgecombe Hospital, Phoenix; Life Pasteur Hospital, Bloemfontein; Life Peglerae Hospital, Rustenburg; Life Poortview Hospital, Roodepoort; Life Queenstown Private Hospital, Queenstown; Life Riverfield Rehab Centre, Randburg; Life Robinson Private Hospital, Randfontein; Life Roseacres Hospital, Germiston; Life Rosepark Hospital, Bloemfontein; Life Springs Parkland Hospital, Springs; Life St Dominic’s Hospital, East London; Life St George’s Hospital, Gqeberha; Life Suikerbosrand Hospital, Gauteng; Life The Glynnwood Hospital, Benoni; Life Vincent Pallotti Hospital, Cape Town; Life West Coast Private Hospital, Vredenburg; Life Westville Hospital, Durban; Life Wilgeheuwel Hospital, Roodepoort; Life Wilgers Hospital, Pretoria; Life the Crompton hospital, Pinetown; Livingstone Hospital, Gqeberha; Louis Pasteur Private Hospital, Pretoria; Louis Trichardt Hospital, Louis Trichardt; Lydenburg Hospital, Lydenburg; Lynnmed Clinic, Pretoria; MANCOVS Surge Facility, Virginia; Maclear Hospital, Maclear; Madadeni Hospital, Newcastle; Madwaleni Hospital, Madwaleni; Madzikane Ka Zulu Memorial Hospital, Mount Frere; Mafube Hospital, Frankfort; Mahatma Gandhi Memorial Hospital, Phoenix; Mahikeng Provincial Hospital, Mahikeng; Malamulele Hospital, Malamulele; Malmesbury Infectious Disease Hosp Wc Mid, Malmesbury; Mamelodi Hospital, Pretoria; Manapo Hospital, Phuthaditjhaba; Manguzi Hospital, Manguzi; Mani Dipico Hospital, Colesberg; Mankweng Hospital, Mankweng; Maphuta L Malatji Hospital, Phalaborwa; Mapulaneng Hospital, Bushbuckridge; Martjie Venter Hospital, Tarkastad; Maseve Filed - Royal Bafokeng (priv), Rustenburg; Matatiele Private Hospital, Matatiele; Matibidi Hospital, Matibidi; Matikwana Hospital, Mkhuhlu; Matlala Hospital, Matlala; Mbongolwane Hospital, Mbongolwane; Mccords Hospital, Berea; Mecklenburg Hospital, Limpopo; Medforum Private Hospital, Pretoria; Medicare private hospital, Rustenburg; Mediclinic Bloemfontein, Bloemfontein; Mediclinic Brits, Brits; Mediclinic Cape Town, Cape Town; Mediclinic Constantiaberg, Cape Town; Mediclinic Durbanville, Cape Town; Mediclinic Ermelo Hospital, Ermelo; Mediclinic Gariep Hospital, Kimberley; Mediclinic Geneva Hospital, George; Mediclinic George Hospital, George; Mediclinic Gynaecological Hospital, Pretoria; Mediclinic Heart Hospital, Pretoria; Mediclinic Hermanus, Hermanus; Mediclinic Highveld Hospital, Trichardt; Mediclinic Hoogland Hospital, Bethlehem; Mediclinic Howick, Howick; Mediclinic Kimberley, Kimberley; Mediclinic Klein Karoo Hospital, Oudtshoorn; Mediclinic Kloof Hospital, Pretoria; Mediclinic Legae Hospital, Mabopane; Mediclinic Lephalale Hospital, Lephalale; Mediclinic Limpopo, Polokwane; Mediclinic Louis Leipoldt, Cape Town; Mediclinic Midstream, Olifantsfontein; Mediclinic Milnerton Hospital, Cape Town; Mediclinic Morningside, Johannesburg; Mediclinic Muelmed Hospital, Pretoria; Mediclinic Paarl Hospital, Paarl; Mediclinic Panorama Hospital, Cape Town; Mediclinic Pietermaritzburg Hospital, Pietermaritzburg; Mediclinic Plettenberg Bay Hospital, Plettenberg Bay; Mediclinic Potchefstroom Hospital, Potchefstroom; Mediclinic Sandton Hospital, Sandton; Mediclinic Stellenbosch, Stellenbosch; Mediclinic Thabazimbi, Thabazimbi; Mediclinic Tzaneen, Tzaneen; Mediclinic Upington, Upington; Mediclinic Vereeniging Hospital, Vereeniging; Mediclinic Vergelegen, Cape Town; Mediclinic Victoria, oThongathi; Mediclinic Welkom Hospital, Welkom; Mediclinic Winelands Orthopaedic Hospital, Stellenbosch; Mediclinic Worcester, Worcester; Medicross Cape Town Foreshore Theatre, Cape Town; Medicross Procare, Kingsburgh; Medicross Richards Bay Day Theatre, Richards Bay; Medicross Upington Day Theatre, Upington; Medicross Wembley House, Pietermaritzburg; Medicross langeberg, Cape Town; Medleb, Lebowakgomo; Melomed Gatesville, Cape Town; Melomed Mitchells Plain Private Hospital, Cape Town; Melomed Richards Bay Private Hospital, Richards Bay; Melomed Tokai Private Hospital, Cape Town; Melomed bellville, Cape Town; Mercantile Private Hospital, Gqeberha; Messina Hospital, Musina; Middelburg Hospital, Middelburg; Midland Hospital, Graaff-Reinet; Midlands Medical Centre Private Hospital, Pietermaritzburg; Midvaal Private Hospital, Vereeniging; Mitchell’s Plain Covid Field Intermediate Care; Mitchell’s Plain District Hosp; Mjanyana Hospital; Mmametlhake Hospital, Napier; Modimolle MDR TB Hospital; Mohau Hospital; Mokopane Hospital; Molteno Hospital; Montagu Hospital; Montebello Hospital; Mooimed Private Hospital; Morehill Clinic Physical rehab; Mossel Bay Hospital; Mosvold Hospital; Mount Ayliff Hospital; Mowbray Maternity Hospital; Mseleni Hospital; Mthatha Private Hospital; Murchison Hospital; Murraysburg Hospital; N17 Hospital; Nala Hospital; Nasrec Quarantine Site; National District Hospital; National Institute for Communicable Diseases, Johannesburg; Nelson Mandela Academic Hospital; Nelspruit Medi Clinic; Nessie Knight Hospital; Netcare Akasia Hospital; Netcare Alberlito Hospital; Netcare Blaauwberg Hospital; Netcare Bougainville Hospital; Netcare Ceres Hospital; Netcare Christiaan Barnard Memorial Hospital; Netcare Clinton; Netcare Cuyler Hospital; Netcare Femina Hospital; Netcare Garden City Hospital; Netcare Greenacres Hospital; Netcare Jakaranda Hospital; Netcare Kingsway Hospital; Netcare Kroon Hospital; Netcare Krugersdorp Hospital; Netcare Kuils River Hospital; Netcare Lakeview Hospital; Netcare Linksfield Hospital; Netcare Linkwood Hospital; Netcare Linmed Hospital; Netcare Margate Hospital; Netcare Milpark Hospital; Netcare Montana Hospital; Netcare Moot Hospital; Netcare Mulbarton Hospital; Netcare N1 City Hospital; Netcare Olivedale Hospital; Netcare Optiklin Hospital; Netcare Parklands Hospital; Netcare Pelonomi Private Hospital; Netcare Pholoso Hospital; Netcare Pinehaven Hospital; Netcare Pretoria-East Hospital; Netcare Rehabilitation Hospital; Netcare Rosebank Hospital; Netcare St Anne’s Hospital; Netcare St Augustine Hospital; Netcare Sunninghill Hospital; Netcare Sunward Park Hospital; Netcare The Bay Hospital; Netcare Umhlanga Hospital; Netcare Union Hospital; Netcare Unitas ECMO Centre; Netcare Unitas Hospital; Netcare Vaalpark Clinic; Netcare Waterfall Hospital; Netcare ferncrest hospital; New Kensington Clinic (Life); New Somerset Hospital; Newcastle Hospital; Newcastle Private Hospital; Ngwelezana Hospital; Nkandla Hospital; Nketoana District Hospital; Nkhensani Hospital; Nkonjeni Hospital; Nkqubela Chest Hospital; Nompumelelo Hospital; Nurture Cape View; Nurture Ilembe; Nurture Newlands; Nurture Queenstown; Nurture Rynmed; Nurture Sunnyside; Nurture Vereeniging; Nurture Woodlands (Mental Health Institution); ODI Community Hospital; Orsmond TB Hospital; Osindisweni Hospital; Othobothini CHC; Otto Du Plessis Hospital; Oudtshoorn Hospital; Paarl Hospital; Park Lane Clinic; Parys Hospital; Pelonomie Hospital; Phekolong Hospital; Philadelphia Hospital; Pholosong Hospital; Phumelela Hospital; Phuthuloha Hospital; Piet Retief Hospital; Polokwane Hospital; Port Alfred Hospital; Port Shepstone Hospital; Postmasburg Hospital; Potchefstroom Hospital; Pretoria Eye Institute Arcadia, Pretoria; Pretoria Urology Hospital, Pretoria; Pretoria West Hospital; Prieska Hospital; Prince Albert Hospital; Prince Cyril Zulu Cdc; Prince Mshiyeni Hospital; Prof ZK Matthews Hospital; Queen Nandi Regional Hospital, Empangeni; RH Matjhabeng Hospital; RH Phodiclinic; RH Piet Retief Hospital; RH Rand Hospital; RK Khan Hospital; Radie Kotze Hospital; Rahima Moosa Mother And Child Hospital; Red Cross Childrens Hospital; Rev Dr Elizabeth Mamisa Chabula-Nxiweni Field Hospital; Richmond Hospital; Riemland Clinic; Rietvlei Hospital; Riversdale Hospital; Rob Ferreira Hospital; Robert Mangaliso Sobukwe Hospital; Robertson Hospital; Rondebosch medical centre; SS Gida Hospital; Sabie Hospital; Sawas Hospital; Schweizer Reneke Hospital; Sebokeng Hospital; Sekororo Hospital; Senorita Ntlabathi District Hospital; Seshego Hospital; Settlers Hospital PPP; Shelly Beach Private Hospital; Shifa private hospital; Shongwe Hospital; Siloam Hospital; Sipetu Hospital; Sizwe Tropical Diseases Hosp; Sonstraal Hospital; South Rand Hospital; Springbok Hospital; St Aidan’s Mission Hospital; St Andrews Hospital; St Apollinaris Hospital; St Barnabas Hospital (Ntlaza); St Elizabeth Hospital; St Francis Hospital (aliwal North); St Helena GM Hospital; St Lucy’s Hospital; St Margarets Hospital; St Mary’s Hospital; St Mary’s Hospital - Umtata; St Patrick’s Hospital; St Rita’s Hospital; Standerton Hospital; Standerton Tb Hospital; Stellenbosch Hospital; Sterkfontein Hospital; Sterkstroom Hospital; Steve Biko Academic Hospital; Steynsburg Hospital; Stikland Psychiatric Hospital; Stoffel Coetzee Hospital; Stutterheim Hospital; Sundays Valley Hospital; Sunningdale Hospital; Sunshine Hospital; Swartland Hospital; Swellendam Hospital; Tafalofefe Hospital; Tambo Memorial Hospital; Tara H Moross Hospital; Taung Hospital; Tayler Bequest Hospital (matatiele); Taylor Bequest Hospital (mount Fletcher); Tb Specialised Hospital (barberton); Telkom Learning Centre Quarantine Site; Tembisa Hospital; Thabamoopo Psychiatric Hosp; Thabazimbi Hospital; The Fountain Private Hospital; Thebe Hospital; Thelle Mogoerane Regional Hospital; Themba Hospital; Thulasizwe Hospital; Thusanong Hospital; Tintswalo Hospital; Tokollo Hospital; Tonga Hospital; Tower Psychiatric Hospital; Town Hill Hospital; Tshepo-Themba Private Hospital; Tshepong Hospital; Tshilidzini Hospital; Tshwane District Hospital, Pretoria; Tshwane Rehabilitation Hospital, Pretoria; Tygerberg Hospital; Uct Private Academic Hospital; Uitenhage Hospital; Umlamli Hospital; Umphumulo Hospital; Umtata General Hospital; Uniondale Hospital; Universitas Hospital; Universitas Private Hospital; Universitas Underground Parking Surge Facility; Untunjambili Hospital; Valkenberg Hospital; Van Velden Hospital; Victoria Hospital; Victoria Hospital (alice); Victoria Private Hospital; Vista Clinic; Voortrekker Hospital; Vredenburg Hospital; Vredendal Hospital; Vryburg private hospital; Vryheid Hospital; WF Knobel Hospital; Warmbaths Hospital; Waterval-boven Hospital; Wentworth Hospital; Wesfleur Hospital; Weskoppies Hospital; West Vaal Hospital; Western Cape Rehabilitation Centre; Wilhelm Stahl Hospital; Willowmore Hospital; Wilmed Park Private Hospital; Winburg Hospital; Winterberg Tb Hospital; Witbank Hospital; Witpoort Hospital; Worcester Hospital Wc Woc; Zebediela Hospital, Magatle; Zithulele Hospital, Mqanduli; Zoutpansberg Private Hospital, Louis Trichardt; Zuid Afrikaans Hospital, Pretoria.

#### South Korea

Chonnam National University Hospital, Dong-gu; Keimyung University Dong San Hospital, Daegu; Kyung Pook National University Chilgok Hospital, Daegu; Seoul National University Bundang Hospital, Seoul; Severance Hospital, Seoul.

#### Spain

Hospital 12 de Octubre, Madrid; Hospital Clinic de Barcelona, Barcelona; Hospital Nuestra Señora de Gracia, Zaragoza; Hospital Puerta de Hierro Majadahonda, Madrid; Hospital Ramon y Cajal, Madrid; Hospital Universitari Sagrat Cor, Barcelona; Hospital Universitari Sant Joan D’Alacant, Alicante; Hospital Universitario Dr Negrín, Las Palmas; Hospital Universitario Virgen de Valme, Seville; Hospital Universitario de Alava, Araba; Hospital Vall d’Hebron, Barcelona; Hospital Verge de la Cinta, Tortosa; Hospital del Mar, Barcelona; La Paz Hospital, Madrid; Reina Sofia University Hospital, Cordoba; Rio Hortega University Hospital, Valladolid; San Pedro de Alcantara Hospital, Cáceres; University Hospital Virgen del Rocío / Institute of Biomedicine of Seville, Seville.

#### Sudan

Atbara Teaching Hospital, Atbara; Jabra Hospital Khartoum, Khartoum; Soba University Hospital, Khartoum. Syrian Arab Republic Al-Mouwasat University Hospital, Damascus; Tishreen University Hospital, Latakia. Taiwan National Taiwan University Hospital, Taipei City. Thailand Siriraj Piyamaharajkarun Hospital (SiPH), Bangkok. Turkey Marmara University Hospital, Istanbul. Uganda Entebbe regional referral hospital, Entebbe; Lubaga hospital, Kampala; Mengo hospital, Kampala; St. Francis hospital Nsambya, Kampala; Uganda Virus Research Institute, Entebbe; Uganda virus research institute clinic, Entebbe.

#### Ukraine

Kharkiv Regional Clinical Infectious Diseases Hospital, Kharkiv; Lugansk State Medical University - Department of Internal Medicine No2, Lugansk.

#### United Arab Emirates

Cleveland Clinic Abu Dhabi, Abu Dhabi; NMC Al Nahda Hospital Dubai, Dubai; NMC Royal Hospital, Abu Dhabi. United Kingdom Aberdeen Royal Infirmary, Aberdeen; Abraham Cowley Unit, Chertsey; Adamson Hospital, Cupar; Addenbrooke’s Hospital, Cambridge; Airedale General Hospital, Steeton; Alder Hey Children’s NHS Foundation Trust, Liverpool; Alderney Hospital, Poole; Amherst Court, Chatham; Arrowe Park Hospital, Wirral; Ashworth Hospital, Liverpool; Barnet Hospital, Barnet; Barnsley Hospital, Barnsley; Basildon University Hospital, Basildon; Basingstoke and North Hampshire Hospital, Basingstoke; Bassetlaw Hospital, Worksop; Bedford Hospital, Bedford; Bedford Hospital South Wing, Bedford; Beech House, Maidstone; Birmingham Children’s Hospital, Birmingham; Birmingham City Hospital, Birmingham; Birmingham and Solihull Mental Health NHS Foundation Trust, Birmingham; Blackberry Hill Hospital, Bristol; Blackpool Victoria Hospital, Blackpool; Blandford Community Hospital, Blandford Forum; Borders General Hospital, Melrose; Bowmere Hospital, Chester; Bradford Royal Infirmary, Bradford; Bridport Community Hospital, Bridport; Brighton General Hospital, Brighton; Bristol Royal Hospital for Children, Bristol; Bristol Royal Infirmary, Bristol; Bronglais General Hospital, Aberystwyth; Broomfield Hospital, Chelmsford; Bryn Beryl Hospital, Pwllheli; Burton Hospital, Burton-on-Trent; CUH at Basildon University Hospital, Basildon; Caithness General Hospital, Wick; Caludon Centre, Coventry; Cameron Hospital, Leven; Campbeltown Hospital, Campbeltown; Castle Hill Hospital, Cottingham; Cefni Hospital, Llangefni; Central Middlesex Hospital, London; Charing Cross Hospital, London; Charlton Lane Hospital, Cheltenham; Chelsea & Westminster Hospital, London; Chesterfield Royal Hospital, Chesterfield; Chirk Community Hospital, Wrexham; Cirencester Hospital, Cirencester; Clatterbridge Cancer Centre -Wirral, Wirral; Cockermouth Hospital, Cockermouth; Colchester General Hospital, Colchester; Colwyn Bay Community Hospital, Colwyn Bay; Community Rehabilitation Team (East), Leicester; Conquest Hospital, Saint Leonards; Copeland Unit, Whitehaven; Countess of Chester Hospital, Chester; County Hospital, Stafford; Cowal Community Hospital, Dunoon; Croydon University Hospital, Thornton Heath; Cumberland Infirmary, Carlisle; Darent Valley Hospital, Dartford; Darlington Memorial Hospital, Darlington; Deeside Community Hospital, Deeside; Denbigh Community Hospital, Denbigh; Derbyshire Healthcare NHS Foundation Trust, Derby; Derriford Hospital, Plymouth; Diana, Princess of Wales Hospital, Grimsby; Dilke Memorial Hospital, Cinderford; Dolgellau & Barmouth District Hospital Site, Dolgellau; Doncaster - Tickhill Road Site, Doncaster; Doncaster Royal Infirmary, Doncaster; Dorset County Hospital, Dorchester; Dumfries & Galloway Royal Infirmary, Dumfries; Ealing Hospital, Southall; East Surrey Hospital, Surrey; Eastbourne District General Hospital, Eastbourne; Eryri Hospital, Caernarfon; Fairfield General Hospital, Bury; Fieldhead Hospital, Wakefield; Forest Close, Sheffield; Forest Lodge, Sheffield; Forth Valley Royal Hospital, Larbert; Frimley Park Hospital, Camberley; Fulbourn Hospital, Cambridge; Furness General Hospital, Barrow-in-Furness; George Eliot Hospital - Acute Services, Nuneaton; Glangwili Hospital Child Health Section, Carmarthen; Glenrothes Hospital, Glenrothes; Gloucestershire Royal Hospital, Gloucester; Golden Jubilee National Hospital, Clydebank; Grantham & District Hospital, Grantham; Great Ormond Street Hospital Central London Site, London; Grenoside Grange, Sheffield; Hairmyres Hospital, Glasgow; Hammersmith Hospital, London; Harefield Hospital, Uxbridge; Harplands Hospital, Stoke-on-Trent; Harrogate District Hospital, Harrogate; Hellesdon Hospital, Norwich; Hereford County Hospital, Hereford; Hillingdon Hospital, Uxbridge; Hinchingbrooke Hospital, Huntingdon; Hollins Park, Warrington; Holywell Community Hospital, Holywell; Homerton University Hospital, London; Huddersfield Royal Infirmary, Huddersfield; Hull Royal Infirmary, Hull; Ipswich Hospital, Ipswich; James Paget University Hospital, Great Yarmouth; John Radcliffe Hospital, Oxford; Kent & Canterbury Hospital, Kent; Keswick Hospital, Keswick; Kettering General Hospital, Kettering; King’s College Hospital (Denmark Hill), London; King’s Mill Hospital, Sutton-in-Ashfield; Kings Court 2, Worcester; Kingston Hospital, Kingston upon Thames; Lantern Centre, Preston; Leeds General Infirmary, Leeds; Leicester Royal Infirmary, Leicester; Leicestershire Partnership NHS Trust Mental Health Services, Leicester; Leighton Hospital, Crewe; Lincoln County Hospital, Lincoln; Lincolnshire Community Health Services Unit 11, Sleaford; Lincolnshire Partnership NHS Foundation Trust, Sleaford; Lister Hospital, Stevenage; Liverpool Heart and Chest Hospital NHS Trust HQ, Liverpool; Liverpool Women’s Hospital, Liverpool; Livewell Southwest, Plymouth; Llandudno General Hospital Site, Llandudno; Lorn and Islands Hospital, Oban; Luton & Dunstable Hospital, Luton; Lynfield Mount Hospital, Bradford; Macclesfield District General Hospital, Macclesfield; Manchester Royal Infirmary, Manchester; Medway Maritime Hospital, Gillingham; Michael Carlisle Centre, Sheffield; Midhurst Community Hospital, Midhurst; Milton Keynes Hospital, Milton Keynes; Mold Community Hospital; Monklands District General Hospital; Moorgreen Hospital; Morriston Hospital; Moseley Hall Hospital; Musgrove Park Hospital; New Cross Hospital; Ninewells Hospital; Noahs Ark Childrens Hospital for Wales; Norfolk & Norwich University Hospital; North Devon District Hospital; North Lincs - Great Oaks Inpatient Unit; North Manchester General Hospital; North Middlesex Hospital; North Tyneside General Hospital; Northampton General Hospital (Acute); Northern General Hospital; Northumbria Specialist Emergency Care Hospital; Northwick Park Hospital; Norwich Community Hospital; Nottingham University Hospitals NHS Trust - City Campus, Nottingham; Nottingham University Hospitals NHS Trust - Queen’s Medical Centre Campus, Nottingham; PHOSP, University of Leicester, Leicester; Penn Hospital; Penrith Hospital; Peterborough City Hospital; Pilgrim Hospital; Pinderfields General Hospital; Poole General Hospital, Poole; Prince Charles Hospital Site; Prince Philip Hospital; Princess Alexandra Hospital, Harlow; Princess Royal Hospital, Haywards Heath; Queen Alexandra Hospital; Queen Elizabeth Hospital, Gateshead; Queen Elizabeth Hospital, London; Queen Elizabeth Hospital, Birmingham; Queen Elizabeth The Queen Mother Hospital, Margate; Queen Elizabeth University Hospital, Glasgow; Queen’s Hospital, Romford; RNI Community Hospital; Raigmore Hospital; Robert Jones & Agnes Hunt Orthopaedic Hospital; Rotherham District General Hospital; Rotherham Older People’s Mental Health Services; Royal Albert Edward Infirmary; Royal Berkshire Hospital; Royal Blackburn Hospital; Royal Bolton Hospital; Royal Bournemouth General Hospital, Bournemouth; Royal Bournemouth Hospital Bcsc, Bournemouth; Royal Brompton Hospital; Royal Cornwall Hospital (Treliske); Royal Derby Hospital; Royal Devon & Exeter Hospital (Wonford); Royal Free Hospital; Royal Gwent Hospital, Newport; Royal Hallamshire Hospital; Royal Hampshire County Hospital; Royal Hospital for Sick Children (Edinburgh); Royal Infirmary of Edinburgh at Little France; Royal Lancaster Infirmary; Royal Liverpool University Hospital; Royal Manchester Children’s Hospital; Royal Oldham Hospital; Royal Orthopaedic Hospital; Royal Papworth Hospital (Papworth Everard); Royal Preston Hospital; Royal Shrewsbury Hospital; Royal Stoke University Hospital; Royal Surrey County Hospital; Royal Sussex County Hospital; Royal United Hospital; Russells Hall Hospital; Ruthin Community Hospital; Salford Royal; Salisbury District Hospital; Sandwell General Hospital; Scarborough General Hospital; Scunthorpe General Hospital; Sheffield Children’s Hospital; Shropshire Community Health NHS Trust HQ; South Tyneside District Hospital; Southampton General Hospital; Southend Hospital; Southern Health & Social Care Trust; Southmead Hospital; Southport & Formby District General Hospital; Springfield University Hospital; St George’s Hospital; St George’s Hospital (Tooting); St Helier Hospital; St James’s University Hospital; St John’s Hospital; St Marks Hospital; St Mary’s Health Campus; St Mary’s Hospital; St Mary’s Hospital (HQ); St Michael’s Hospital; St Nicholas Hospital (Newcastle Upon Tyne); St Peter’s Hospital, Surrey; St Richard’s Hospital; St Thomas’ Hospital; Stepping Hill Hospital; Stockdale House; Stoke Mandeville Hospital; Stroud General Hospital; Sunderland Royal Hospital; Sussex Partnership NHS Foundation Trust, Worthing; Tameside General Hospital; Tees, Esk, Wear Valley NHS Trust (Durham); Tewkesbury General Hospital; The Christie NHS Foundation Trust, Manchester; The Great Western Hospital; The James Cook University Hospital; The Longley Centre; The Maidstone Hospital; The Mount, Leeds; The Queen Elizabeth Hospital; The Royal Glamorgan Hospital; The Royal London Hospital, London; The Royal Marsden Hospital (London); The Royal Marsden Hospital (Surrey); The Royal Victoria Infirmary; The Tunbridge Wells Hospital; The Walton Centre NHS Foundation Trust; The Whittington Hospital; Torbay Hospital; Trafford General Hospital; Trengweath Mental Health Unit, Redruth; Trust Head Office; Trust Headquarters; University College Hospital; University College London Hospitals NHS Foundation Trust HQ; University Hospital Aintree; University Hospital Ayr; University Hospital Coventry; University Hospital Crosshouse; University Hospital Lewisham; University Hospital of North Durham; University Hospital of North Tees; University Hospital of Wales; University of Edinburgh, Edinburgh; Urgent Care Centre; Victoria Hospital; Victoria Hospital W’Borne; Walsall Manor Hospital, Walsall; Walton Hospital; Wansbeck Hospital; Warrington Hospital; Warwick Hospital; Watford General Hospital; West Cumberland Hospital; West Middlesex University Hospital; West Suffolk Hospital; Western Community Hospital; Western General Hospital; Westminster Memorial Hospital; Westmorland General Hospital; Weston General Hospital; Wexham Park Hospital; Weymouth Community Hospital; Whiston Hospital; William Harvey Hospital (Ashford); Wishaw General Hospital; Withybush General Hospital; Wonford House; Worcestershire Royal Hospital, Worcester; Worthing Hospital, Worthing; Wotton Lawn Hospital, Gloucester; Wrexham Maelor Hospital, Wrexham; Wycombe Hospital, High Wycombe; Wythenshawe Hospital, Manchester; Yeovil District Hospital, Yeovil; York Hospital, York; Ysbyty Alltwen, Gwynedd; Ysbyty Cwm Cynon, Mountain Ash; Ysbyty Cwm Rhondda, Tonypandy; Ysbyty Glan Clwyd, Bodelwyddan; Ysbyty Gwynedd, Bangor; Ysbyty Penrhos Stanley, Holyhead.

#### United States of America

Allegheny General Hospital, Pittsburgh; Avera McKennan Hospital & University Health Center, South Dakota; Baylor All Saints Medical Centre, Fort Worth; Baylor Scott & White Health, Temple; Baylor University Medical Centre, Dallas; Baystate MC, Springfield; Beth Israel Deaconess, Boston; Beth Israel Deaconess Medical Center, Boston; Brigham and Women’s Hospital, Boston; Brooke Army Medical Centre, San Antonio; Carilion Clinic, Roanoke; Cedars-Sinai MC, Los Angeles; Cleveland Clinic, Weston; Cleveland Clinic Foundation, Cleveland; Cleveland Clinic, Ohio, Ohio, OH; Columbia University, New York; Denver Health MC, Colorado; Duke University MC, North Carolina; Emory University Healthcare System, Atlanta; Harborview MC, Washington; Hartford HealthCare, Hartford; Hennepin County MC, Minneapolis; Henry Ford Health System, Detroit; Houston Methodist Hospital, Texas; INOVA Alexandria Hospital, Alexandria, Virginia; INOVA Fair Oak Hospital, Chantilly, Virginia; INOVA Fairfax Heart Hospital, Annandale, Virginia; INOVA Fairfax Medical Center, Fairfax, Virginia; INOVA Fairfax Women & Children Hospital, Annandale, Virginia; INOVA Loudoun Hospital, Virginia; INOVA Mount Vernon Hospital, Hybla Valley, Virginia; Intermountain Medical Center, Utah; Johns Hopkins, Baltimore; Lancaster General Health, Pennsylvania; Lankenau Institute of Medical Research, Wynnewood; Legacy Emanuel Medical Center, Portland; Louisiana State University MC, Louisiana; Maine Health, Portland; Massachusetts General Hospital, Massachusetts; Mayo Clinic School of Medicine, Arizona; McLeod Healthcare System, Florence; MedStar Washington Hospital Centre, Washington; Medical College of Wisconsin, Wisconsin; Medical University of South Carolina, South Carolina; Montefiore MC, New York; Montefiore MC North, New York; Montefiore MC Weiler, New York; Mount Sinai Medical Center, Miami; National Jewish Health / St Joseph, Colorado; Nationwide Children’s Hospital, Columbus; Northwell Health, New York; Ochsner Clinic Foundation, New Orleans; Ohio State East Hospital, Columbus; Ohio State University, Columbus; Oklahoma Heart Institute, Oklahoma; Oregon Health and Science University, Portland; Perelman School of Medicine at the University of Pennsylvania, Philadelphia; Piedmont Atlanta Hospital, Atlanta, Georgia; Presbyterian Hospital Services, Alberquerque; Providence Saint John’s Health Centre, Santa Monica; Rochester General Hospital, New York; Rush University Medical Center, Chicago; Sentara Norfolk General Hospital, Norfolk; St Christopher’s Hospital for Children, Philadelphia; St Vincent, Minnesota; Stanford University, Palo Alto; Stanford University Hospital, California; Swedish Hospital Cherry Hill, Washington; Swedish Hospital First Hill, Washington; Temple University Hospital, Pennsylvania; The Christ Hospital, Ohio; The Heart Hospital Baylor Plano, Plano; The Mount Sinai Hospital, New York; Tufts Medical Centre, Boston; UH Cleveland Hospital, Cleveland; UPMC, Pittsburgh; UPMC Mercy, Pittsburgh; UPMC Shadyside, Pennsylvania; US NHLBI PETAL Network, Boston; UT Southwestern, Dallas; University of Alabama at Birmingham Hospital, Birmingham; University of Arizona, Arizona; University of California - San Francisco (UCSF), San Francisco; University of California Davis, California; University of California Ronald Reagan MC, California; University of California SF, California; University of California SF Fresno, California; University of California San Francisco - Fresno, Fresno; University of Chicago, Chicago; University of Cincinnati, Cincinnati; University of Cincinnati MC, Cincinnati; University of Colorado Hospital, Colorado; University of Florida, Gainesville; University of Florida HSC Shands, Gainesville; University of Iowa, Iowa City; University of Kansas Medical Center, Kansas; University of Kentucky, Kentucky; University of Maryland, Baltimore; University of Michigan MC, Michigan; University of Michigan Schools of Medicine & Public Health, AnnArbor; University of Mississippi MC, Mississippi; University of Nebraska Medical Center, Omaha; University of Oklahoma Health Sciences Center, Oklahoma; University of Texas HSC Houston, Texas; University of Utah, Salt Lake City; University of Utah HSC, Utah; University of Virginia MC; University of Washington MC, Washington; University of Washington Medical Center - Northwest, Seattle; Vanderbilt University MC, Tennessee; Virginia Commonwealth University, Richmond; Wake Forest Baptist Health, North Carolina; Washington University in St. Louis, St Louis, Missouri.

#### Viet Nam

Hospital for Tropical Diseases, Ho Chi Minh City.

